# Deciphering epistatic genetic regulation of cardiac hypertrophy

**DOI:** 10.1101/2023.11.06.23297858

**Authors:** Qianru Wang, Tiffany M. Tang, Michelle Youlton, Chad S. Weldy, Ana M. Kenney, Omer Ronen, J. Weston Hughes, Elizabeth T. Chin, Shirley C. Sutton, Abhineet Agarwal, Xiao Li, Merle Behr, Karl Kumbier, Christine S. Moravec, W. H. Wilson Tang, Kenneth B. Margulies, Thomas P. Cappola, Atul J. Butte, Rima Arnaout, James B. Brown, James R. Priest, Victoria N. Parikh, Bin Yu, Euan A. Ashley

## Abstract

Although genetic variant effects often interact non-additively, strategies to uncover epistasis remain in their infancy. Here, we develop low-signal signed iterative random forests to elucidate the complex genetic architecture of cardiac hypertrophy, using deep learning-derived left ventricular mass estimates from 29,661 UK Biobank cardiac MRIs. We report epistatic variants near *CCDC141*, *IGF1R*, *TTN*, and *TNKS*, identifying loci deemed insignificant in genome-wide association studies. Functional genomic and integrative enrichment analyses reveal that genes mapped from these loci share biological process gene ontologies and myogenic regulatory factors. Transcriptomic network analyses using 313 human hearts demonstrate strong co-expression correlations among these genes in healthy hearts, with significantly reduced connectivity in failing hearts. To assess causality, RNA silencing in human induced pluripotent stem cell-derived cardiomyocytes, combined with novel microfluidic single-cell morphology analysis, confirms that cardiomyocyte hypertrophy is non-additively modifiable by interactions between *CCDC141*, *TTN*, and *IGF1R*. Our results expand the scope of cardiac genetic regulation to epistasis.

## Main

Heart disease is closely tied to the structure of the heart^1^. Heart failure, a syndrome characterized by increased pressure within, or decreased output from, the heart is influenced by structural features including atrial and ventricular chamber size and wall thickness^2,3^. Left ventricular hypertrophy – increased thickness of the left ventricle (LV) – can be the result of mendelian genetic diseases like hypertrophic cardiomyopathy^4^ but is also a complex phenotypic trait influenced by multiple factors, genetic and environmental. Progressive LV hypertrophy carries significant independent risk for incident heart failure, atrial arrhythmia, and sudden death^5,6^, highlighting the need to understand genetic determinants of cardiac phenotype.

Recent discoveries leveraging cardiac magnetic resonance imaging in the UK Biobank (UKBB) have revealed that cardiac structure is in part determined by complex genetics^7–9^. Common genetic variants, many located near genetic loci associated with dilated cardiomyopathy and heart failure, have been found to influence LV size and systolic function^7^. Further, specific genetic variants that influence LV trabeculation have been shown to impact systolic function and overall risk of cardiomyopathy^8^.

However, these variants remain inadequate to explain the total heritable disease risk^10^. Indeed, common genetic variants rarely act independently and additively as modeled by most genome-wide association studies (GWAS)^11^. There is growing biological and clinical evidence to support a disease risk model in which multiple genes interact non-additively with each other through epistasis^12,13^. While some computational studies estimated a minor average epistatic component compared to the additive component within the total genetic variance, these epistatic variance estimates exhibit a large trait-to-trait variation^14^. In addition, it’s important to distinguish between the concepts of statistical epistasis, estimated through variance components and influenced by allele frequencies, and biological epistasis (e.g., gene actions), which is independent from allele frequencies^15^. Recent work has shown that common genetic variation influences susceptibility and expressivity of hypertrophic cardiomyopathy^9^. This raises the possibility that common epistatic interactions drive cardiac phenotype, holding significant potential for uncovering disease mechanisms and developing potential therapeutic strategies.

Several computational and experimental challenges need to be resolved to allow robust identification of epistasis. First, the combinatorial nature of possibly high-order interactions makes an exhaustive search computationally intractable. To reduce the computational burden and ensure stable discoveries, we developed an approach based on signed iterative random forests^16,17^ to uncover higher-order (not limited to pairwise) nonlinear interactions in a computationally-tractable manner. Second, many previously reported epistatic relationships were not replicated^18^. To achieve more trustworthy results, we adhered to a new framework for veridical data science^19^, centered around the principles of predictability, computability, and stability (PCS) and the need for transparent documentation of decisions made in data analysis pipelines. A third challenge is the generally small effect size of common genetic variants^10^ which impedes both the data-driven discovery and functional validation of epistatic interactions. In human biobanks, recent advances in deep-learning-enabled phenotyping^20^ using cardiac magnetic resonance images have led to more refined phenotypes at larger scales. At the cellular level, high-throughput microfluidic technologies^21,22^ have been integrated with artificial intelligence-based image analysis of single-cell morphology^23^ and human induced pluripotent stem cell-derived cardiomyocytes^24^, opening up new possibilities for rapid, label-free detection of the phenotypic consequences of genetic perturbation.

## Results

In contrast to many studies^12,14,18^ that have investigated the statistical significance or causality of epistasis solely from observational data, we tackle the aforementioned challenges and conceptual gap between statistical epistasis and biological epistasis^15^ via a multi-stage approach. This approach begins with a data-driven prioritization of promising statistical epistasis followed by extensive functional interpretations and experimental validations to reliably assess the biological epistasis consistency.

More specifically, our methodology includes four major stages: derivation of estimates of LV mass (green boxes, Fig. 1); computational prioritization of epistatic drivers (orange boxes, Fig.1); functional interpretation of the hypothesized epistatic genetic loci (purple boxes, Fig.1); and experimental confirmation of epistasis through perturbation (blue boxes, Fig. 1).

**Fig. 1:**
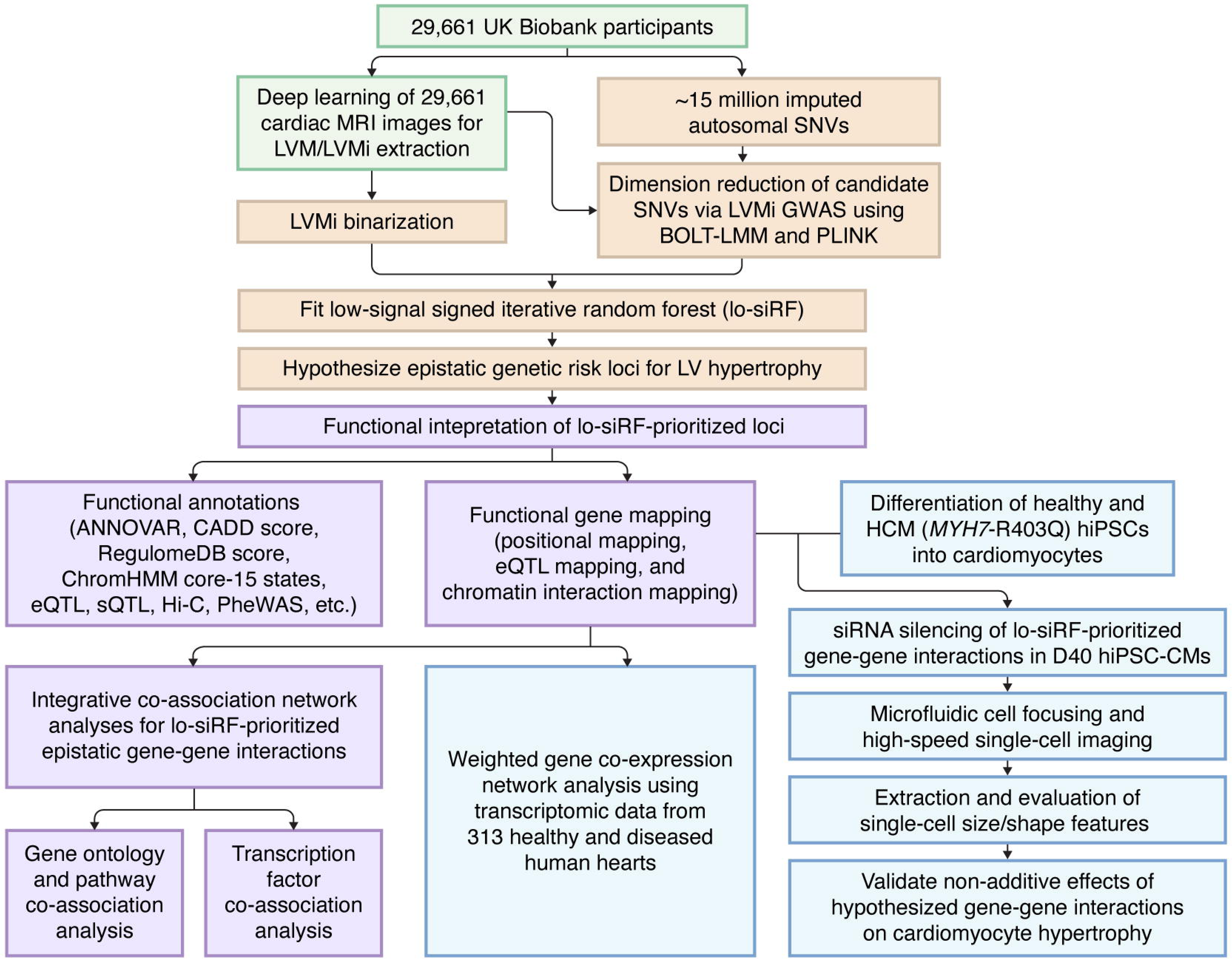
Schematic of the study workflow. The study workflow includes four major stages: (a) derivation of left ventricular mass from cardiac magnetic resonance imaging (green boxes); (b) computational prioritization of epistatic drivers (orange); (c) functional interpretation of the hypothesized epistatic genetic loci (purple); and (d) experimental confirmation of epistasis in cardiac tissues and cells (blue). Abbreviations: MRI, magnetic resonance imaging; LV, left ventricle; LVM, left ventricular mass; LVMi, left ventricular mass indexed by body surface area; SNV, single-nucleotide variant; GWAS, genome-wide association study; PLINK^26^ and BOLT-LMM^27^, two different GWAS software packages; lo-siRF, low-signal signed iterative random forest; ANNOVAR^74^, a software for functional annotation of genetic variants; CADD^36^, combined annotation dependent depletion, which scores the deleteriousness of variants; RegulomeDB^35^, a database that scores functional regulatory variants; ChromHMM^34^, a multivariate Hidden Markov Model for chromatin state annotation; eQTL, expression quantitative trait locus; sQTL, splicing quantitative trait locus; Hi-C, high-throughput chromosome conformation capture; PheWAS, phenome-wide association study; siRNA, small interfering RNA; hiPSC-CM, human induced pluripotent stem cell-derived cardiomyocyte; HCM, hypertrophic cardiomyopathy.

### Deep learning of cardiac imaging quantifies LV hypertrophy

We accessed all cardiac magnetic resonance images from the UKBB substudy (44,503 people at the time of this analysis)^25^. We focused on the largest ancestry subset of 29,661 unrelated individuals (summary characteristics in Supplementary Table 1) and analyzed the most recent image per individual. We leveraged a recent deep learning model^20^ to quantify LV hypertrophy from these 29,661 multislice cine magnetic resonance images (Fig. 2a). A fully convolutional network had been previously trained for image segmentation and was evaluated on manual pixelwise-annotations of images from 4,875 UKBB participants^20^. This fully convolutional network learns features across five different resolutions through sequential convolutional layers interspersed with non-linearities, and has displayed accurate performance compared to cardiac segmentation by human experts^20^. Using this segmentation model, we extracted areas of the LV chamber wall in each slice of the short axis image at the end of diastole. Areas extracted from each image slice in the same image stack were then integrated to calculate the heart muscle volume, which was converted to LV mass and normalized by body surface area to obtain the LV mass index (LVMi, Extended Data Fig. 1). Details regarding this analysis can be found in Methods.

**Fig. 2:**
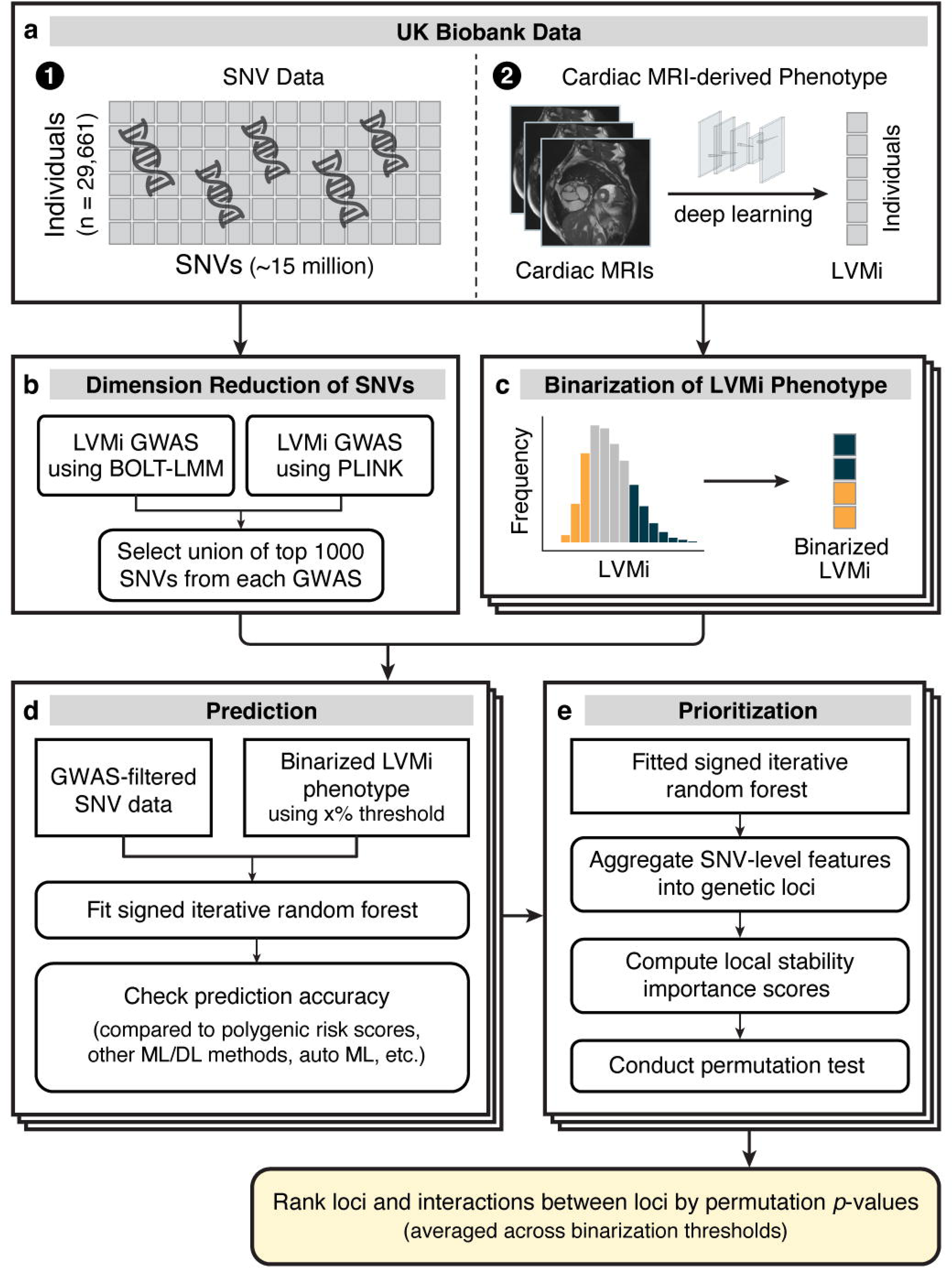
Low-signal signed iterative random forest (lo-siRF) workflow. **a**, Lo-siRF took as input single-nucleotide variant (SNV) data and cardiac MRI-derived left ventricular mass indexed by body surface area (LVMi) from 29,661 UK Biobank participants. **b**, Two genome-wide association studies (GWAS) were applied using different software to reduce the dimensionality of SNVs. **c**, LVMi was binarized into high and low categories using three thresholds (shown as stacked boxes). **d**, For each threshold, a signed iterative random forest (siRF) was fitted using the GWAS-filtered SNVs to predict the binarized LVMi, achieving prediction accuracy better than or comparable to other methods, before interpreting the model fit. **e**, siRF aggregates SNVs into genetic loci using ANNOVAR^74^ and finally ranks loci and their interactions based on a stability-driven importance score averaged across the three binarization thresholds.

### Low-signal signed iRF prioritizes epistatic genetic loci

We developed low-signal signed iterative random forests (lo-siRF, Fig. 2) to prioritize statistical epistatic interactions from the extracted LV mass and single-nucleotide variants (SNVs) from UKBB. Given the inherent low signal-to-noise ratio and aforementioned challenges, lo-siRF aims to recommend reliable candidate interactions for experimental validation rather than directly assessing claims of statistical significance from data. This prioritization pipeline is guided by the PCS framework^19^ and builds upon signed iterative random forests^16,17^, a computationally-tractable algorithm to extract predictive and stable nonlinear higher-order interactions that frequently co-occur along decision paths in a random forest. More specifically, lo-siRF proceeds through four steps:

1. *Dimension reduction* (Fig. 2b): we combined the results of two initial genome-wide association studies, implemented via PLINK^26^ and BOLT-LMM^27^ (Extended Data Fig. 2, Supplementary Data 1) to reduce the interaction search space from 15 million imputed variants down to 1405 variants (Supplementary Data 2). Details can be found in the Methods section *Lo-siRF step 1: Dimension reduction of variants via genome-wide association studies*.
2. *Binarization* (Fig. 2c): we partitioned the LV mass measurements into high, middle, and low categories using three different partitioning schemes (Supplementary Table 2). The partitioning enabled us to transform the original low-signal regression problem for a continuous trait into a relatively easier binary classification task for predicting individuals with high versus low LV mass measurements (omitting the middle category). This transformation is necessary to obtain a sufficient prediction signal, ensuring that the model indeed captures pertinent information about reality (Supplementary Table 3). Further justification and details on the partitioning can be found in the Methods section *Lo-siRF step 2: Binarization of the left ventricular mass phenotype*.
3. *Prediction* (Fig. 2d): we trained a signed iterative random forest using the 1405 GWAS-filtered SNVs to predict the binarized LV mass measurements. The learnt model yields on average the highest (balanced) classification accuracy (55%), area under the receiver operator characteristic (0.58), and area under the precision-recall curve (0.57) compared to other common machine learning prediction algorithms (Supplementary Table 4). Details about the model and prediction check can be found in the Methods section *Lo-siRF step 3: Prediction*.
4. *Prioritization* (Fig. 2e): we developed a stability-driven feature importance score (Extended Data Fig. 3), which leveraged the fitted signed iterative random forest and a permutation test, to aggregate SNVs into genetic loci and prioritize interactions between genetic loci. This importance score provides the necessary new interpretable machine learning ingredient to complete the lo-siRF discovery pipeline. Details can be found in the Methods section *Lo-siRF step 4: Prioritization*.

Additional discussion of the philosophy and modeling decisions driving lo-siRF can be found in Supplementary Note 1, an interactive HTML webpage hosted at https://yu-group.github.io/epistasis-cardiac-hypertrophy/. The webpage also provides a comparison of lo-siRF to alternative epistasis detection methods (implementation details in Supplementary Note 2), including an exhaustive regression-based pairwise interaction search^28^, MAPIT^29^, and a locus-level variant of MAPIT using gene set enrichment analysis^30^, demonstrating the challenges and limitations of existing methods for analyzing low-signal, complex phenotypes.

Lo-siRF identified six genetic risk loci that exhibited stable and reliable associations with LV mass (Table 1). Because these loci are either located within a gene body or in between two genes (Fig. 3a), for convenience we denote these loci by their nearest genes. Notably, out of the six loci, three (*TTN*, *CCDC141*, and *IGF1R*) were prioritized by lo-siRF as epistatic loci. These loci not only interact with other loci, but also marginally affect LV mass. The other three lo-siRF-prioritized loci are *LOC157273;TNKS*, *MIR588;RSPO3*, and *LSP1*. The *LOC157273;TNKS* locus is located within the intergenic region between genes *LOC157273* and *TNKS* (semicolon indicates intergenic region). This locus was prioritized by lo-siRF to be hypostatic (i.e., effects are deemed stable by lo-siRF only when interacting with the *CCDC141* locus). Interestingly, all three identified epistatic interactions involved the *CCDC141* locus (Fig. 3a, green links in circle 1). Furthermore, while the *MIR588;RSPO3* and *LSP1* loci lacked evidence for epistasis by lo-siRF, they were each identified to be marginally associated with LV mass. The specific prioritization order of these loci can be found in Supplementary Table 5, and details regarding the direction or sign of the interactions can be found in Supplementary Note 1. In total, lo-siRF identified 283 SNVs located within the six loci (Supplementary Data 3, Extended Data Fig. 4).

**Table 1.** Lo-siRF-prioritized risk loci and epistatic interactions for left ventricular hypertrophy.

| Lo-siRF-prioritized LV hypertrophy risk loci |  |  |  |  |  |  |  |  |  |  |  |  |  |  |
| --- | --- | --- | --- | --- | --- | --- | --- | --- | --- | --- | --- | --- | --- | --- |
| Lo-siRF locus<sup>1</sup> | # IndSig SNVs<sup>2</sup> | # SNVs<sup>3</sup> | Nominal lo-siRF p-value<sup>4</sup> | Nominal lo-siRF p-value<sup>5</sup><br>(excluding hypertension) | Max CADD<sup>6</sup> | Min RDB<sup>7</sup> | Top SNV<sup>8</sup> | Chr:Pos<sup>9</sup> | MAF<sup>9</sup> | NEA/EA<sup>9</sup> | Function | GWAS p-value<sup>9</sup> | GWAS Beta<sup>9</sup> | GWAS SE<sup>9</sup> |
| ***TTN*** | 52 | 148 | 0.009 | 0.002 | 25.5 | 1b | rs66733621 | 2:178799323 | 0.290 | A/G | intronic | 8.78E-6 | 0.0437 | 0.00983 |
| ***CCDC141*** | 65 | 85 | 0.018 | <1E-3 | 27.3 | 1a | rs7591091 | 2:178889467 | 0.298 | C/T | Intronic | 3.67E-11 | -0.0646 | 0.00975 |
| *MIR588;RSPO3* | 75 | 218 | 0.002 | 0.071 | 20.2 | 1b | rs9401921 | 6:126925592 | 0.379 | A/G | intergenic | 3.14E-7 | 0.0474 | 0.00926 |
| ***LOC157273;TNKS*** | 76 | 50 | 0.142 | 0.172 | 16.88 | 1b | rs6999852 | 8:9478458 | 0.301 | A/G | intergenic | 1.15E-5 | -0.0426 | 0.00970 |
| *LSP1* | 9 | 11 | 0.017 | <1E-3 | 10.91 | 1b | rs569550 | 11:1865838 | 0.386 | T/G | ncRNA intronic | 3.85E-6 | 0.0423 | 0.00915 |
| ***IGF1R*** | 6 | 60 | <1E-3 | 0.002 | 17.46 | 1a | rs62024491 | 15:98733068 | 0.312 | G/A | intronic | 9.43E-6 | -0.0426 | 0.00962 |
| Lo-siRF-prioritized epistatic interactions between LV hypertrophy risk loci |  |  |  |  |  |  |  |  |  |  |  |  |  |  |
| Epistatic interaction<sup>1</sup> | Nominal lo-siRF p-value<sup>4</sup> |  | Nominal lo-siRF p-value<sup>5</sup><br>(excluding hypertension) |  | Top SNV-SNV interaction<sup>8</sup> |  | Chr:Pos<sup>9</sup> |  | # Partner SNVs<sup>10</sup> |  |  |  |  |  |
| ***CCDC141-IGF1R*** | <1E-3 |  | <1E-4 |  | rs7591091<br>rs62024491 |  | 2:178889467<br>15:98733068 |  | 133<br>62 |  |  |  |  |  |
| ***CCDC141-TTN*** | 0.011 |  | <1E-3 |  | rs7591091<br>rs66733621 |  | 2:178889467<br>2:178799323 |  | 133<br>61 |  |  |  |  |  |
| ***CCDC141-LOC157273;TNKS*** | 0.056 |  | 0.007 |  | rs7591091<br>rs6999852 |  | 2:178889467<br>8:9478458 |  | 133<br>59 |  |  |  |  |  |

**Fig. 3:**
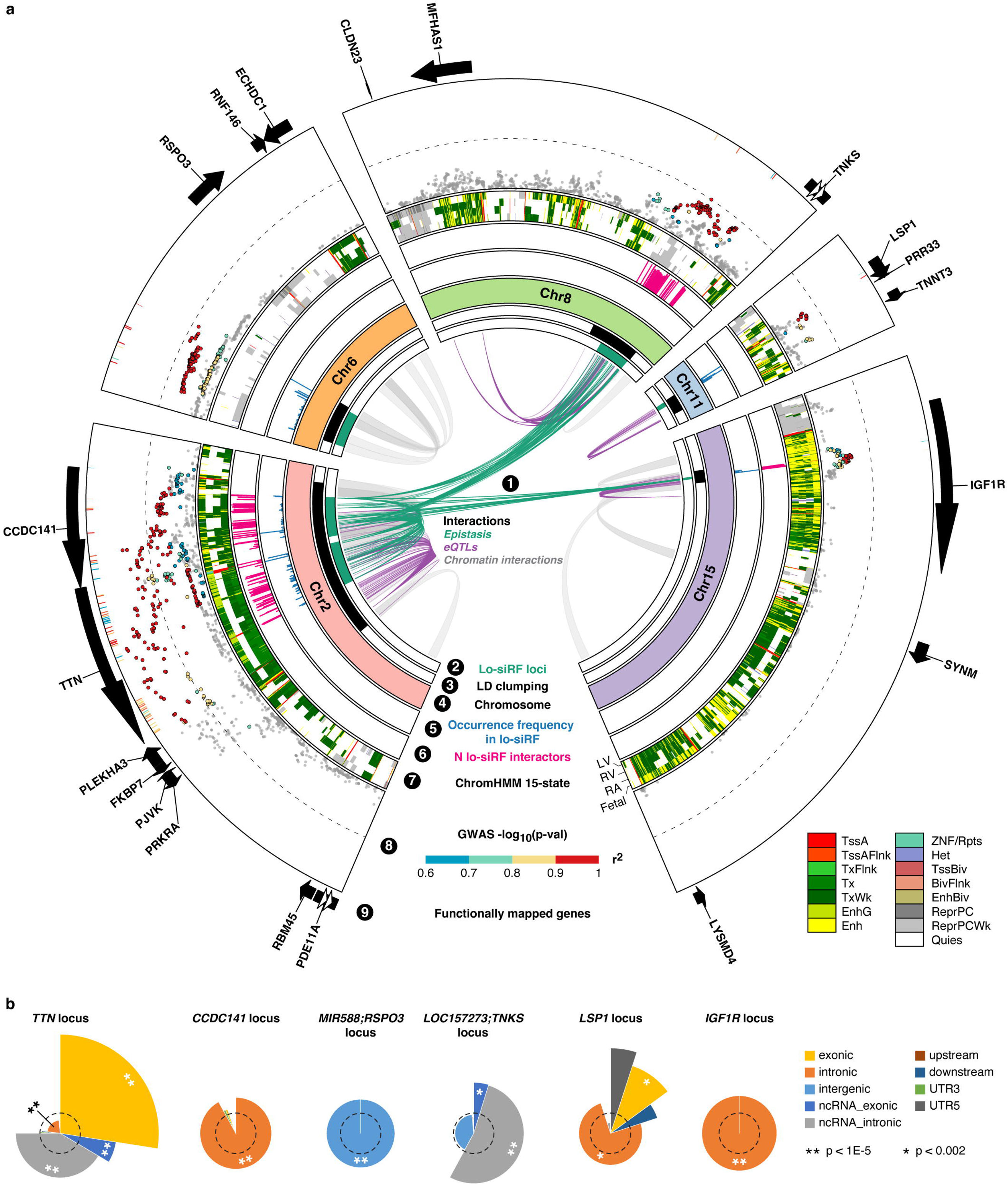
Lo-siRF finds epistasis between genetic risk loci for left ventricular hypertrophy. **a,** Circos plot illustrating genetic risk loci identified by lo-siRF (green, circle 2) and regions after clumping FUMA-extracted SNVs in LD (*r*^2^ > 0.6) with lo-siRF-prioritized SNVs (black, circle 3). Circle 1 shows the top 300 epistatic SNV-SNV pairs ranked by occurrence frequency in lo-siRF (green), SNV-gene linkages (FDR < 0.5) from GTEx^37^ V8 *cis*-eQTL data of heart and skeletal muscle tissues (purple), and 3D chromatin interactions^33^ from Hi-C data of left ventricular tissue (GSE87112). Circle 5 and 6 show occurrence frequencies and the number of partner SNVs in epistasis (normalized by the maximum value per locus) identified by lo-siRF, respectively. Circle 7 displays the ChromHMM^34^ core-15 chromatin state for left ventricle (LV), right ventricle (RV), right atrium (RA), and fetal heart (Fetal). Circle 8 presents a GWAS Manhattan plot (points, two-sided *p* < 0.05 from PLINK^26^), where SNVs are color-coded by their maximum *r*^2^ value relative to the 283 lo-siRF-prioritized SNVs. The outer heatmap layer represents LD-linked SNVs (*r*^2^ > 0.6) extracted from the FUMA reference panel, which lack GWAS *p*-values. SNVs not in LD (*r*^2^ ≤ 0.6) with any of the 283 lo-siRF-prioritized SNVs appear in gray. The dashed line indicates GWAS *p* = 5E-8. Circle 9 shows genes functionally mapped by FUMA. **b**, Pie charts depicting ANNOVAR enrichment results for the six lo-siRF loci (circle 2 in Fig. 3a and Table 1). Slice arc length represents the proportion of SNVs with a specific functional annotation. Slice radius indicates log_2_(E + 1), where E is the enrichment score computed as (proportion of SNVs with an annotation for a given locus)/(proportion of SNVs with an annotation relative to all available SNVs in the FUMA reference panel). The dashed circle indicates E = 1 (no enrichment). Asterisks denote two-sided Fisher’s exact test *p*-values for the enrichment of each annotation. Details in Supplementary Data 4 and 5.

Ninety percent of the 283 SNVs have previously been shown to harbor multiple distinct cardiac function associations^31^ in phenome-wide analyses (e.g., pulse rate, Supplementary Data 3), suggesting a strong likelihood that these lo-siRF-prioritized loci contribute to determining cardiac structure and function.

Considering the correlations between LV hypertrophy and hypertension^32^, we evaluated whether these identified variants affect LV mass through regulating blood pressure. Specifically, we repeated the lo-siRF analysis using only the subset of UKBB individuals without hypertension (details in Methods). All previously highlighted loci and interactions maintained priority in this non-hypertensive subset, except for the *MIR588;RSPO3* locus (Table 1) which was not stably prioritized across all three binarization thresholding schemes. Additionally, none of the lo-siRF-prioritized variants showed a strong marginal association with hypertension, failing to meet the genome-wide (*p* < 5E-8) and even the suggestive (*p* < 1E-5) significance level. However, the *MIR588;RSPO3* locus with lead SNV rs2022479 gave the smallest *p*-value of 5E-5, which may suggest a possible pleiotropic effect of *MIR588;RSPO3* on both LV hypertrophy and blood pressure. In brief, while we cannot completely rule out pleiotropy, the highly stable prioritization of all three epistatic interactions in both analyses with and without hypertensive individuals suggest that the identified epistases on LV mass is not solely driven by blood pressure (additional discussion in Supplementary Note 1).

### Loci associated with LV mass exhibit regulatory enrichment

We performed functional mapping and annotation (FUMA)^33^ for the 283 lo-siRF-prioritized SNVs (Fig. 1, purple and Fig. 3). For linkage disequilibrium (LD), we used a default threshold of *r*^2^ = 0.6 and chose the UKBB release 2b reference panel created for British and European subjects to match the population group used for lo-siRF prioritization. FUMA identified 572 additional candidate SNVs (Supplementary Data 4) in strong LD (*r*^2^ > 0.6) with any of the 283 lo-siRF-prioritized SNVs, including 492 SNVs from the input GWAS associations (points in Fig. 3a, circle 8) and 80 non-GWAS-tagged SNVs extracted from the selected reference panel (heatmap tracks in Fig. 3a, circle 8). We then assigned these 572 FUMA-extracted candidate SNVs to a lo-siRF-prioritized locus (Table 1) based on the corresponding lo-siRF-prioritized SNV (out of the 283 SNVs), which has the maximum *r^2^* value with the candidate SNV.

The two loci contributing to the top-ranked epistatic interaction by lo-siRF, the *CCDC141* and *IGF1R* loci (Table 1), both showed a significant enrichment of intronic variants relative to the background reference panel (Fig. 3b, Supplementary Data 5). Over 88% of the SNVs in or in LD with these two loci were mapped to actively transcribed chromatin states (TxWk) or enhancer states (Enh) in left ventricles based on the ChromHMM Core 15-state model^34^ (Fig. 3a, circle 7). More than 47% and 76% of the identified SNVs in or in LD with the *CCDC141* and *IGF1R* loci, respectively, showed the highest RegulomeDB^33,35^ categorical score (ranked within category 1 from the 7 main categories). The Combined Annotation-Dependent Depletion (CADD) score^36^ was used to judge the deleteriousness of prioritized variants (Supplementary Data 4). As expected, GTEx^37^ data revealed that 82% of SNVs in or in LD with the *IGF1R* locus are expression quantitative trait loci (eQTLs) for the gene *IGF1R*. In contrast, of the SNVs in or in LD with the *CCDC141* locus, only 14% are eQTLs for gene *CCDC141* and 22% are splicing quantitative trait loci (sQTLs) for gene *FKBP7*. Furthermore, Hi-C data indicated that all SNVs identified in or in LD with the *IGF1R* locus are in 3D chromatin interaction with gene *SYNM* while more than 54% SNVs identified in or in LD with the *CCDC141* locus are in 3D chromatin interaction with gene *TTN*. These known 3D chromatin interactions could suggest a possibility of higher-order interactions between more than two genes.

The *CCDC141* and *TTN* loci exhibit genomic proximity (Fig. 3a). Their interaction, however, does not appear to stem from this proximity. Indeed, the *CCDC141* and *TTN* genes have been individually associated with LV mass^38,39^. Due to this proximity, previous studies^40,41^ have assumed *CCDC141* as a secondary gene that affects LV mass through the *TTN* gene expression. However, we found low LD (*r^2^* < 0.6) between any two of the 283 lo-siRF-prioritized SNVs, suggesting that the identified *CCDC141-TTN* interaction is unlikely driven by non-random LD associations between SNVs in these two loci. In addition, we compared all the epistasis-contributing SNVs that were aggregated to the *TTN* locus, including both lo-siRF-prioritized SNVs and their LD-linked variants, with the complementary set of *TTN*-annotated SNVs in lo-siRF. We found that the *TTN* locus showed a significant depletion of SNVs located close to (<10 kb) the gene *CCDC141* (*p* = 2.38E-9, two-sided Fisher’s exact test). Similarly, the *CCDC141* locus showed a substantially decreased enrichment of SNVs that are close to gene *TTN* (*p* = 0.02, two-sided Fisher’s exact test). These results suggest that although the *CCDC141* and *TTN* loci are located close to each other in the genome, the prioritized epistatic SNVs are located farther apart relative to randomly selected SNVs from the two loci.

In contrast to the *CCDC141* and *IGF1R* loci, the *TTN* locus showed a significant enrichment of exonic variants and intronic variants that are transcribed into non-coding RNA (ncRNA_intronic, Fig. 3b). Of those exonic variants, 62% are nonsynonymous. This differential enrichment of exonic variants for the *TTN* locus may suggest a potential epistatic contribution to the structural alterations in the titin protein. Over 90% of SNVs in or in LD with the *TTN* locus were mapped to actively transcribed states (Tx, TxWk) in left ventricles (Fig. 3a, circle 7). Interestingly, these SNVs were associated with a quiescent chromatin state (Quies) in the right atrium, indicating that the epistatic effects of the *TTN* locus may be specific to ventricular tissues. Nearly half of SNVs in or in LD with the *TTN* locus are eQTLs for the gene *FKBP7*. In addition, 83% of these SNVs are sQTLs for gene *FKBP7* or *TTN*, suggesting a regulatory effect of the *TTN* locus on the expression and splicing of gene *FKBP7*. Moreover, the *TTN* locus was suggested to impact genes *PDE11A*, *RBM45*, *PRKRA*, and *PJVK* through 3D chromatin interactions.

The hypostatic locus *LOC157273;TNKS* showed a significant enrichment of variants within non-coding RNA regions of exons and introns (Fig. 3b). Over 95% of identified SNVs in or in LD with this locus were mapped to inactive chromatin states (ReprPCWk, Quies) in left ventricles (Fig. 3a, circle 7). This suggests that in the absence of an epistatic partner, the *LOC157273;TNKS* locus is epigenetically quiescent or repressed by polycomb group proteins. In addition, of all the SNVs in or in LD with this locus, 66% are eQTLs for *MFHAS1* or *CLDN23* and 22% are in 3D chromatin interaction with gene *TNKS*.

Functional annotations for the other two lo-siRF-prioritized loci that were marginally associated with LV mass can be found in Supplementary Data 4 and 5.

### Epistatic loci functionally map to 20 protein-coding genes

Three strategies, positional, eQTL, and chromatin interaction, mapped the six LV hypertrophy risk loci to 20 protein-coding genes (Fig. 4a). Genes prioritized by eQTL and chromatin interaction mapping are not necessarily located in the corresponding risk locus, but they are linked to SNVs within or in LD with the locus (Fig. 3a). Among the 20 genes, *CCDC141* and *IGF1R* were prioritized by all the three mapping strategies (Fig. 4a), suggesting that these two genes are very likely involved in determining LV mass. Interestingly, none of the SNVs mapped to *IGF1R* were statistically significant in our GWAS studies using BOLT-LMM and PLINK (Extended Data Fig. 2 and Supplementary Data 1). Set-based association tests^42^ also failed to identify the *IGF1R* locus (details in Methods and Supplementary Note 1). This reveals the potential of lo-siRF to identify risk loci that may be overlooked by GWAS. Based on the expression data from GTEx V8, *TTN*, *TNNT3*, and *SYNM* are up-regulated while *CLDN23* and *MFHAS1* are down-regulated in both heart and muscle tissues (Fig. 4b). In contrast, *CCDC141* is up-regulated specifically in heart tissues whereas *RSPO3* is down-regulated in heart but up-regulated in muscle tissues (Fig. 4b).

**Fig. 4:**
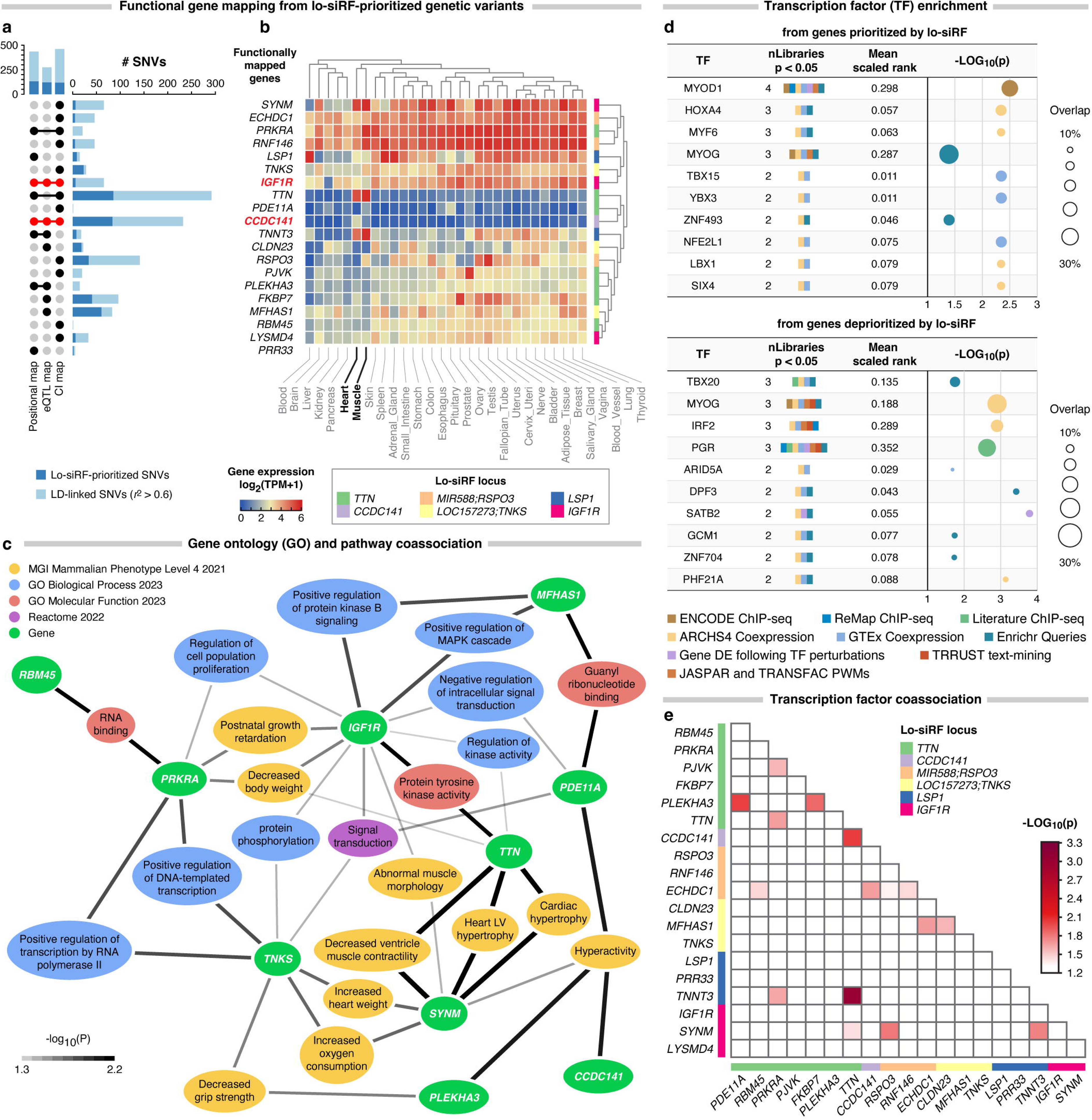
Genes mapped from epistatic loci show strong functional co-associations. **a**, UpSet plot showing the number of lo-siRF-prioritized SNVs (dark blue) and their LD-linked (*r*^2^ > 0.6) SNVs (light blue, circle 8 in Fig. 3a) functionally mapped to 20 protein-coding genes via positional, eQTL, and/or chromatin interaction (CI) mapping in FUMA^33^. *CCDC141* and *IGF1R* (highlighted red) are prioritized by all three mapping strategies (details in Supplementary Data 4). **b**, Heatmap of averaged expression (GTEx, 50% winsorization, log_2_(TPM + 1)) across different tissues for these functionally mapped genes. **c**, Co-association network built from an enrichment analysis integrating multiple annotated gene set libraries for gene ontology (GO) and pathway terms from Enrichr^43^. Nodes represent genes (green) functionally linked from lo-siRF-prioritized epistatic and hypostatic loci (Table 1) and top enriched GO/pathway terms. Edge width indicates the co-association strength between enriched terms and genes ranked by two-sided *p*-values from an exhaustive permutation of the co-association score for all possible gene-gene combinations in the network (details in Methods and Supplementary Data 6). **d,** Comparison between the top 10 transcription factors (TFs) enriched from genes prioritized (top) and deprioritized (bottom) by lo-siRF. Prioritized genes are functionally linked to lo-siRF-prioritized SNVs (panel **a**), while deprioritized genes are genes linked to SNVs failed to pass the lo-siRF prioritization thresholds. Enrichment was performed against nine TF-annotated gene set libraries from ChEA3^44^ and Enrichr^43^, ranked by the number of significantly (*p* < 0.05, two-sided Fisher’s exact test) overlapped libraries (numbers in *nLibraries* column) and the mean scaled rank across all libraries containing that TF (colored boxes in *nLibraries* column). The balloon plot shows the lowest two-sided Fisher’s exact test *p*-values (horizontal axis) and the proportion of overlapped genes (balloon size) between the input gene set and the corresponding TF-annotated gene set. **e**, Heatmap showing the TF co-association strength of gene-gene combinations among lo-siRF-prioritized genes relative to randomly selected gene pairs in the co-association network. Further details in Extended Data Fig. 5 and Supplementary Data 7.

### Epistatic genes show strong network co-associations

We performed gene ontology (GO) and pathway enrichment analysis on the 20 genes mapped from lo-siRF loci. We adopted previously established approaches^43,44^ and integrated enrichment results across libraries from multiple sources to establish a GO and pathway co-association network (Fig. 4c). To evaluate the correlation strength between any two genes in the network, we calculated a co-association score for every possible gene-gene combination (n = 72,771) from both genes prioritized and deprioritized by lo-siRF. Lo-siRF-prioritized genes are the 20 genes functionally mapped from the 283 lo-siRF-prioritized SNVs and their LD-linked SNVs (Fig. 4a). Lo-siRF-deprioritized genes are those functionally mapped from the SNVs that failed to pass the lo-siRF prioritization threshold. Compared to random gene pairs in the network, 10 genes that were functionally mapped from the lo-siRF-prioritized epistatic and hypostatic loci showed significant co-associations with multiple GO/pathways (Fig. 4c, Supplementary Data 6). Consistent with our hypothesized epistasis (Table 1), gene *CCDC141* showed a significant co-association to *SYNM* (functionally linked to the *IGF1R* locus) and *PDE11A* and *PLEKHA3* (both functionally linked to the *TTN* locus) through the GO term of hyperactivity (excessive movement), which has been linked to increased risk of cardiac disease^45^. Beyond that, *TTN*, *IGF1R*, and *SYNM* are co-associated with kinase activity and cardiac structure related GO terms, indicating that these genes may jointly affect cardiac structure by regulating the process of kinase activity.

### Epistatic genes share myogenic regulatory factors

We next performed an integrative enrichment analysis to assess transcriptional regulation of genes prioritized and deprioritized by lo-siRF. Due to assay-specific limitations and biases, we integrated the enrichment results across nine distinct gene set libraries^43,44^ (Fig. 4d, Supplementary Data 7). We found that the lo-siRF-prioritized epistatic genes shared important myogenic regulatory factors, such as MYOD1, MYF6, and MYOG (Fig. 4d, top). These myogenic regulatory factors coordinate to regulate muscle development and differentiation. In contrast, transcription factors enriched from lo-siRF-deprioritized genes display a less coordinated regulatory pattern (Fig. 4d, bottom). These analyses enriched transcription factors based on their associations to given sets of individual genes rather than co-association to gene pairs^43,44^. To further evaluate the correlation strength between any two genes that share transcription factors, we calculated a transcription factor co-association score for all the 72,771 possible gene-gene combinations (see Methods). Compared with random gene pairs,16 gene-gene combinations from the lo-siRF-prioritized genes displayed a significant co-association (empirical *p* < 0.05, Fig. 4e). These co-associations were found in gene-gene combinations from both intra- and inter-lo-siRF-prioritized loci (Fig. 4e). In particular, pairwise combinations among *TTN*, *TNNT3*, *CCDC141*, and *SYNM* share a common splicing regulator, RBM20 (Extended Data Fig. 5). RBM20 has been reported to regulate the alternative splicing of genes important for cardiac sarcomere organization^46^. This suggests that the splicing patterns of these four genes are likely to be co-regulated by RBM20, which is consistent with the exhibited enrichment of sQTLs by the *CCDC141*, *TTN* and *LSP1* lo-siRF loci (Supplementary Data 4).

### Human heart transcriptomic correlations reflect epistasis

We proceeded to the fourth stage for experimental confirmation (Fig. 1, blue) and evaluated how the identified epistases contribute to the progression of heart failure (Fig. 5). We employed a series of weighted gene co-expression networks derived from human cardiac transcriptomic data from 177 failing hearts isolated at the time of heart transplant and 136 non-failing hearts harvested from cardiac transplant donors whose organs were not able to be placed^47^ (Fig. 5a). We compared the molecular connectivity of genes identified by lo-siRF as statistical epistatic interactors. We defined connectivity as the edge weights between two genes normalized to the distribution of all network edge weights, and compared this to the connectivity of all other available gene-gene combinations in the network. This revealed strong co-expression correlations between *CCDC141* and genes functionally linked to the *IGF1R* locus (*SYNM* and *LYSMD4*) and *TTN* locus (*TTN* and *FKBP7*) in the healthy control network (Fig. 5b). In contrast, most of these gene pairs (except for *CCDC141*-*TTN*) no longer exhibit a strong connectivity in the heart failure network (Fig. 5c). All of these connectivities showed a significant decrease (indicated by the negative connectivity difference score and *p* < 0.05 in Fig. 5d) in the differential network, suggesting a declined co-expression correlation between these gene pairs relative to random gene pairs during the progression of failing hearts. This difference is potentially related to the rewired gene modular assignments between the control and heart failure networks^47^ (Fig. 5e and Extended Data Fig. 6). For instance, *CCDC141*, *SYNM*, *TTN*, and *TNNT3* are co-associated with the electron transport chain/metabolism module in the control network. In the failing hearts, *SYNM* and *TTN* rewire to the muscle contraction/cardiac remodeling module, whereas *CCDC141* and *TNNT3* remain associated with the metabolism module (Fig. 5e). In addition, other genes functionally linked to *IGF1R* and *TTN* lo-siRF loci are co-associated with the membrane transport or unfolded protein response module in healthy hearts and rewire to the muscle contraction/cardiac remodeling or cell surface/immune/metabolism module in failing hearts.

**Fig. 5:**
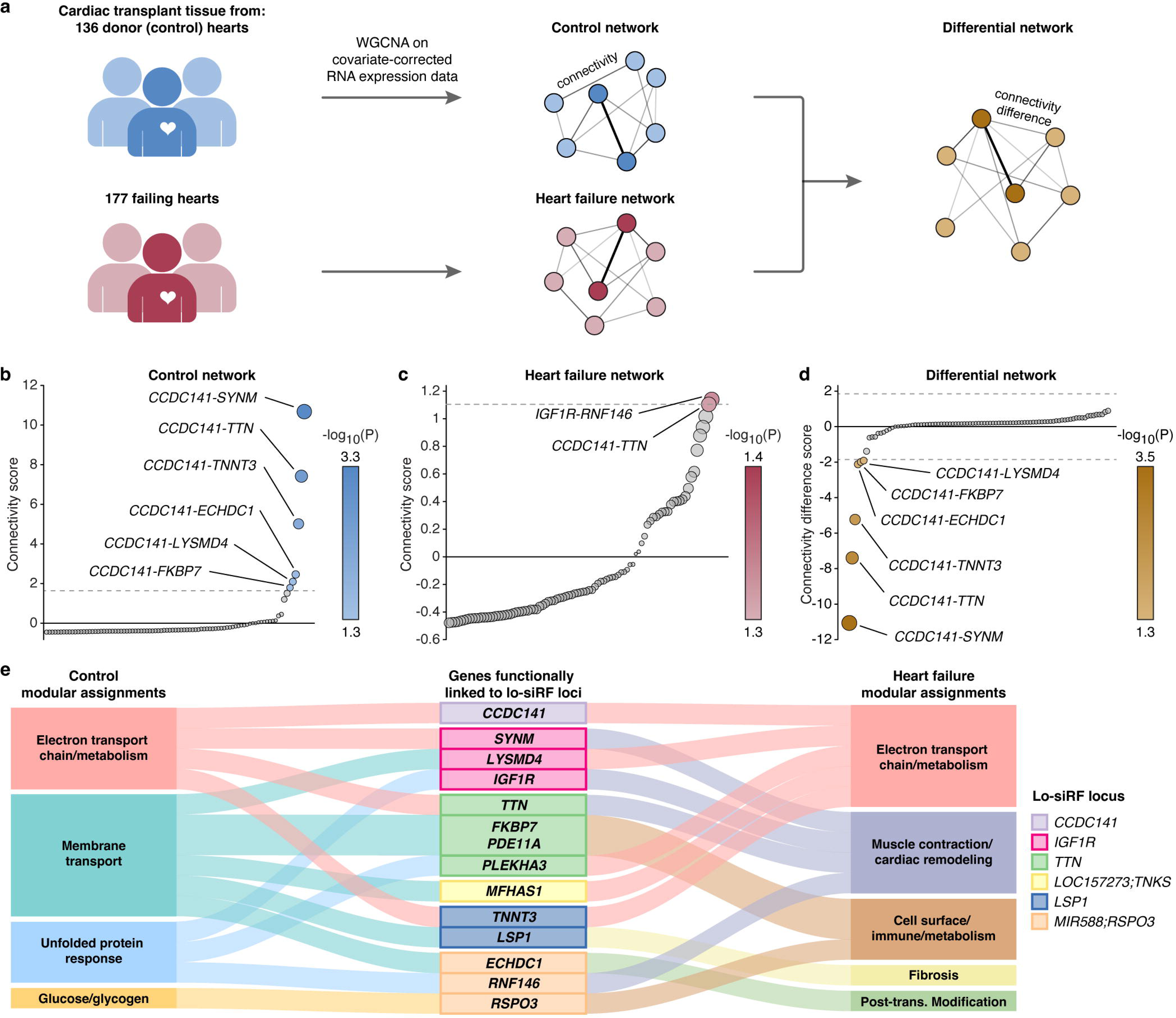
Network analysis using transcriptomic data from 313 human hearts reveals strong correlations between suggestive statistical epistasis contributors. **a**, Gene co-expression networks for control (blue) and heart failure (red) conditions were established using weighted gene co-expression network analysis (WGCNA) on transcriptomic data from 313 non-failing and failing human heart tissues^47^. **b-c**, The connectivity between lo-siRF-prioritized genes was assessed against the full connectivity distributions of all possible gene-gene combinations in the control (**b**) and heart failure (**c**) networks. *CCDC141* showed a significant connectivity to *SYNM* and *LYSMD4* (*IGF1R* lo-siRF locus) and *TTN* and *FKBP7* (*TTN* lo-siRF locus) in the control network. Color scales indicate two-sided empirical *p*-values. **d**, Comparing between the control and heart failure networks indicated a significant connectivity decrease of these gene pairs during heart failure progression. **e**, A Sanky plot demonstrating the rewired gene modular assignments for the lo-siRF loci-associated genes (middle column) in the control vs. heart failure networks. Module names in control (left column) and heart failure (right column) networks were derived from KEGG and Reactome associations of genes within each module.

### Perturbation confirms epistasis in cardiomyocyte hypertrophy

We interrogated epistatic associations in a genetic model of cardiac hypertrophy (Fig. 1, blue): induced pluripotent stem cell cardiomyocytes derived from patients with and without hypertrophic cardiomyopathy caused by the cardiac myosin heavy chain (*MYH7)* p.R403Q variant^24^ (Fig. 6a).

**Fig. 6:**
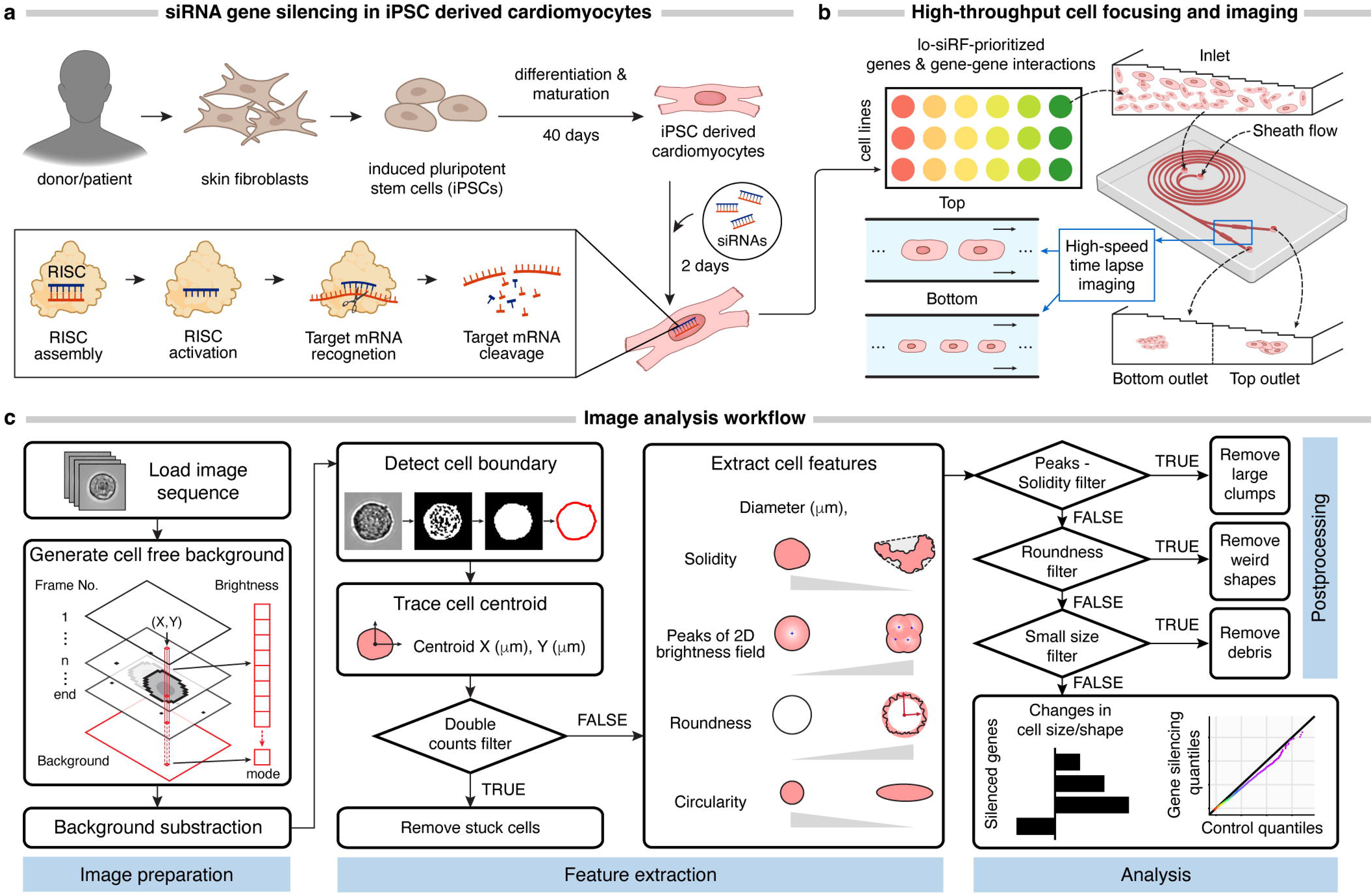
Single-cell image analysis of epistatic gene-silencing effects on cardiomyocyte hypertrophy. **a**, Human induced pluripotent stem cell (iPSC)-derived cardiomyocytes, with and without hypertrophic cardiomyopathy (*MYH7*-R403Q mutation), were transfected with scramble siRNA or siRNAs specifically targeting single (*CCDC141*, *IGF1R*, *TTN*) or combined (*CCDC141*-*IGF1R*, *CCDC141*-*TTN*) loci prioritized by lo-siRF. **b**, Gene-silenced cardiomyocytes were bifurcated into large and small cells using a spiral microfluidic device (Extended Data Fig. 7) for high-resolution single-cell imaging. **c**, Image analysis workflow. Single-cell images were analyzed using a customized MATLAB program to extract cell size/shape features through a multi-step process.

Cardiac myosin heavy chain 7 is a key component of the cardiac sarcomere, and the most common cause of hypertrophic cardiomyopathy^24^. The patient presented with typical symptoms, and echocardiography revealed severe LV hypertrophy and a small LV cavity^24^. At the cellular level, cardiomyocytes exhibit an elevated mean cell size and non-Gaussian size distribution with a long tail relative to the unaffected control (Fig. 7a).

**Fig. 7:**
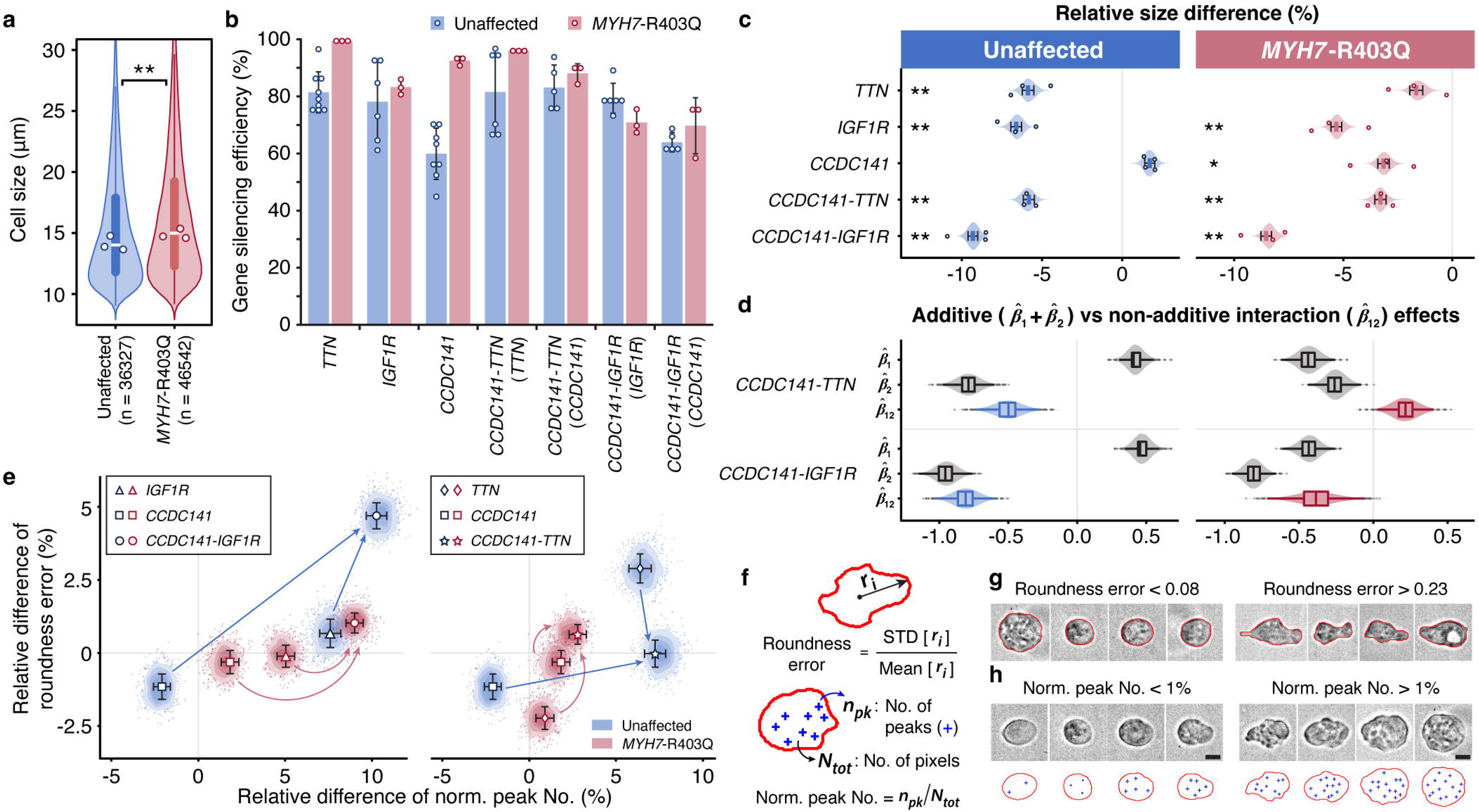
*CCDC141* non-additively interacts with *TTN* and *IGF1R* to modify cardiomyocyte morphology. **a**, Violin plots showing total cell diameter distributions for unaffected (blue, *n* = 36327) and *MYH7*-R403Q variant (red, *n* = 46542) cardiomyocytes from three independent cell batches (*p* = 2.1E-36, two-sided Mann-Whitney U test). In all panels, boxplots display the interquartile range (IQR) with the median line, and whiskers extend to 1.5x IQR. Overlaid points represent median diameters from individual batches. **b**, Gene-silencing efficiency measured by RT-qPCR in unaffected (blue) and *MYH7*-R403Q variant (red) cells from at least *n* = 3 cell batches (exact *n* numbers provided in Source Data). Bars, overlaid points, and error bars show mean values, individual biological replicates, and standard deviations. **c-h**, Gene-silencing effects on single-cell morphology analyzed for *n* = 3 independent cell batches (*n* = 4 for unaffected cells silencing *CCDC141*), with over 33,000 cells measured per condition per batch. **c**, Percent change in median cell diameter for each gene-silencing condition relative to scramble controls (i.e., relative size difference). Squares represent mean relative size differences across batches, with overlaid points indicating individual batches. Violins display mean relative size differences from 1,000 bootstrap samples, with error bars indicating standard deviations. Asterisks: \**p* < 0.05, \*\**p* < 6E-5 (exact *p*-values in Source Data), two-sided Mann-Whitney U test. **d**, Violin plots showing non-additive interaction effects (*β̂*_12_) for unaffected (blue) and *MYH7*-R403Q variant (red) cells compared to marginal effects (*β̂* _1_ and r*β̂*_2_, gray), estimated via a quantile regression model across 10,000 bootstrap samples (details in Methods). **e**, Scatter plots showing gene-silencing effects on cellular texture (i.e. norm. peak No., *x*-axis) and boundary (i.e. roundness error, *y*-axis) features. Markers represent mean values across cell batches, with error bars indicating standard deviations from 1,000 bootstrap samples. Bootstrapped values are visualized as density contours (scattered dots represent a density level lower than 0.2). **f**, Cell boundary waviness and texture irregularity were measured by roundness error (top) and normalized peak number (bottom), respectively. **g**-**h**, Representative single-cell images illustrating boundary irregularity (**g**) and textural variation (**h**). Scale bars: 10 μm. Statistical details in Supplementary Data 8 and Source Data.

To determine if *CCDC141* can act both independently and in epistatic interactions with other genes to attenuate the pathologic cellular hypertrophy caused by *MYH7*-R403Q, we silenced genes *CCDC141*, *IGF1R*, *TTN*, and gene pairs *CCDC141*-*IGF1R* and *CCDC141*-*TTN* using siRNAs in both diseased and healthy cardiomyocytes and compared them with cells transfected with scramble siRNAs (control) (Fig. 6a and 7b). Phenotypic consequences of these perturbations on cellular morphology were then evaluated in high-throughput using a spiral inertial microfluidic device (Fig. 6b) in combination with automated single-cell image analysis (Fig. 6c). The microfluidic device adopted the Dean flow focusing principle^22^ (details in Extended Data Fig. 7 and Methods) to mitigate the non-uniform cell focusing^48^, thereby enhancing the imaging resolution^49^ affected by the large variations in cardiomyocyte diameter (Fig. 7a).

We first assessed the knockdown effects of the *CCDC141*-*IGF1R* interaction on cardiomyocyte size (Fig. 7c). Bootstrapped hypothesis tests were performed, for which the *p*-values are capped below by *p* < 1E-4 (Supplementary Data 8). Silencing *IGF1R* alone reduces the median cell size by 5.3% ± 0.3% (*p* < 1E-4) in diseased cells compared to scrambled control and 6.6% ± 0.3% (*p* < 1E-4) in healthy cells. Silencing *CCDC141* alone also decreases median cell size by 3.2% ± 0.3% (*p* < 1E-4) in diseased cells, but had no impact on healthy cells. Digenic silencing of *CCDC141* and *IGF1R* reveals a synergistic effect on attenuating pathologic cell hypertrophy in diseased cells, resulting in an 8.5% ± 0.2% (*p* < 1E-4) decrease in the median cell size. This is consistent in healthy cells, where silencing *CCDC141* alone fails to affect cell size, but digenic silencing of *CCDC141* and *IGF1R* decreases the median cell size by 9.3% ± 0.3% (*p* < 1E-4). Moreover, according to our estimated quantile regression analysis (details in Methods), this interaction effect appears to be non-additive for both healthy and diseased cells (r _12_< 0, Fig. 7d; *p <* 1E-4 for non-additivity, Supplementary Data 8), consistent with an epistatic mechanism. These findings serve to confirm the strongest epistatic association identified by lo-siRF (Table 1).

We found a comparable non-additive effect for the *CCDC141*-*TTN* interaction. Digenic silencing of *CCDC141*-*TTN* leads to a pronounced reduction in median cell size (by 5.8% ± 0.3% for healthy cells and 3.3% ± 0.3% for diseased cells, *p* < 1E-4) relative to monogenic silencing (Fig. 7c). This interaction appears to be non-additive for both healthy and diseased cells (*p*-values in Supplementary Data 8) yet demonstrating opposite epistatic directions in these two cell states (Fig. 7d). Additionally, *CCDC141* and *TTN* show distinctive independent roles in repressing cardiomyocyte hypertrophy. In healthy cells, monogenic silencing of *TTN* leads to a larger cell size reduction compared to the case of silencing *CCDC141*. In contrast, diseased cells display a larger size reduction in response to monogenic silencing of *CCDC141*.

Furthermore, both *CCDC141*-*IGF1R* and *CCDC141*-*TTN* interactions show a stronger effect on rescuing larger cardiomyocytes over smaller ones in both cell lines (Extended Data Fig. 8 and 9). In contrast, monogenic silencing does not exhibit such a non-uniform effect on reshaping the cell size distribution, which reinforces the hypothesized non-additivity of these two epistatic interactions (details in Supplementary Data 8 and Supplementary Note 4).

Recent studies have shown that cellular morphological features, such as cell boundary and textural irregularities, are informative readouts of cytoskeletal structure, which is highly associated with disease state in hypertrophic cardiomyopathy^23,50^. We analyzed relative changes in cell shape and texture (Fig. 7e) by measuring the counts of peak intensities normalized to the total number of pixels enclosed by the cell boundary (Fig. 7f). Cells with a high normalized peak number display a ruffled texture, which manifests in unevenly distributed 2D intensities (Fig. 7h). Our analysis shows that silencing both *CCDC141* and *IGF1R* (circles in Fig. 7e, left) yields a larger increase in intensity peak number than silencing *IGF1R* alone (triangles in Fig. 7e, left) for both cell lines, exhibiting a synergistic epistasis between *CCDC141* and *IGF1R* (*p* < 1E-4 for non-additivity). We also analyzed cell roundness error, a measure of how far radii measured on the cell outline deviate from a perfect circle (Fig. 7f). This parameter increases with an increasing cell boundary waviness or elongation (Fig. 7g). We show that the silencing of *CCDC141* and *IGF1R* synergistically interact to increase roundness error of diseased cardiomyocytes (*p* < 1E-4 for non-additivity, Fig. 7e, left). In addition, *CCDC141* and *TTN* display antagonistic epistasis and synergistic epistasis in their impact on roundness error for healthy and diseased cells (*p* < 1E-4 for non-additivity, Fig. 7e, right), respectively.

## Discussion

While computational models^12,13^ have supported epistatic contributions to human complex traits and disease risk, examples in the literature are rare, with even fewer experimentally confirmed. Here, we developed a veridical machine learning^19^ approach to identify epistatic associations with cardiac hypertrophy derived from a deep learning model that estimates LV mass from cardiac imaging of almost thirty thousand individuals in the UK Biobank. We report novel epistatic effects on LV mass of common genetic variants associated with *CCDC141*, *TTN*, and *IGF1R.* We used established tools to functionally link risk loci to genes, and then confirmed gene level co-associations through network analyses, including via shared transcription factors and pathways enriched against multiple annotated gene set libraries and co-expression networks we built using transcriptomic data from over three hundred healthy and diseased human hearts. Finally, using a cellular disease model incorporating monogenic and digenic silencing of individual genes, we assessed phenotypic changes in cardiomyocyte size and morphology using a novel microfluidic system, confirming the non-additive nature of the interactions.

Our approach advances epistasis discovery in several key ways. First, unlike studies relying on linear-based models^51,52^, we leverage a more realistic, nonlinear tree-based model that mirrors the thresholding (or switch-like) behavior commonly observed in biomolecular interactions^53^. Second, in contrast to other tree-based approaches that evaluate interactions on a variant-by-variant basis^54,55^, our novel stability-driven importance score consolidates individual variants into loci for the assessment of feature importance, allowing for more reliable extraction of epistatic interactions from weak association signals. This is particularly valuable for evaluating non-coding variants and resembles ideas from marginal association mapping with sets of SNVs^42,56^. Moreover, instead of exhaustively searching all possible interactions, signed iterative random forests internally employ a computationally-efficient algorithm, which automatically narrows the search space of interactions to only those that stably appear in the forest and thus achieves a scalability much higher than existing tree-based approaches^54^. This allows lo-siRF to handle larger datasets without the need for LD pruning before the interaction search, which may inadvertently eliminate important epistatic variants, given that epistasis between loci in strong LD has been evidenced by a recent study^57^. Furthermore, our computational prioritization is rigorously validated through multiple functional network analyses and robust experimental confirmation.

Our results add to a small literature on epistasis in cardiovascular disease. Two recent studies have found epistasis influencing the risk of coronary artery disease^12,13^. Li et al.^13^ identified epistasis between *ANRIL* and *TMEM106B* in coronary artery tissues. Although their method predicted functionally interpretable interactions between risk loci of interest, they relied heavily on prior knowledge and careful selection of the causal gene pairs,^13^ making the approach challenging to scale. Zeng et al.^12^ used population-scale data and performed epistasis scans from regions around 56 known risk loci. This study identified epistasis between variants in *cis* at the *LPA* locus without experimental confirmation. In contrast, our approach allows discovery of not only *cis*-epistasis, but also long-range interactions between interchromosomal loci (e.g., *CCDC141* and *IGF1R*) and is supported by gene perturbation experiments. More importantly, both studies searched for interactions around known risk loci identified by genome-wide association, which can be far away from the possible epistatic or hypostatic loci that are statistically insignificant in linear univariate association studies. In addition, both studies relied on a logistic regression model, which imposes restrictive assumptions that can be avoided using a nonlinear machine learning approach as in lo-siRF.

Our study has limitations. Given our primary interest in biological epistasis rather than statistical epistasis^15^, we tailored lo-siRF to conservatively prioritize reliable targets for experimental validation as opposed to finding all possible epistatic drivers. Lo-siRF should ideally be used as a first-stage hypothesis generation tool within a broader scientific discovery pipeline. To assess significance of the lo-siRF-prioritized targets, we rely on and encourage follow-up investigations such as the high-throughput gene-silencing experiments conducted here. We focused this analysis on a single ancestry in order to enhance the likelihood of finding reliable interactions from weak association signals. These findings cannot be automatically applied to others. It was not feasible to conduct a formal genetic replication study because the UK Biobank is the only large-scale population cohort with integrated cardiac magnetic resonance images and genetic data. However, to help reduce the possibility of overfitting and increase generalizability, lo-siRF employed numerous stability analyses (see Supplementary Note 1) in addition to a proper training-validation-test data split. Beyond these computational checks, we also present functional supporting evidence and experimental validation. Our computational prioritization via lo-siRF currently groups SNVs based on genomic proximity, without accounting for their functional interdependencies, but this could be addressed by integrating functional annotation into the lo-siRF pipeline. Lo-siRF also relies on a GWAS to reduce the number of SNVs to a computationally manageable size, but this could be improved with more sophisticated epistasis detection algorithms such as MAPIT^29^. Lastly, lo-siRF is not as scalable as linear-based methods, though it is more scalable than alternative tree-based methods for epistasis detection. It also should be noted that although this study did not identify stable higher-order (> order-2) interactions due to the weak association signal between SNVs and LV mass, the method exhibits the capability to detect such interactions for broader phenotypes and complex traits without incurring additional computational cost.

In summary, our work adds to the discovery toolkit for the genomic architecture of complex traits and expands the scope of genetic regulation of cardiac structure to epistasis.

## Methods

### Study participants

The use of human subjects (IRB - 4237) and human-derived induced pluripotent stem cells (SCRO - 568) in this study has been approved by the Stanford Research Compliance Office. The UK Biobank received ethical approval from the North West - Haydock Research Ethics Committee (21/NW/0157).

The UK Biobank (UKBB) is a biomedical database with detailed phenotypic and genetic data from over 500,000 UK individuals aged 40 - 69 at recruitment^58^. In this study, we focused on the largest ancestry subset (i.e., the White British population) of 29,661 unrelated individuals who have both genetic and cardiac magnetic resonance imaging (MRI) data (Supplementary Table 1). More specifically, we considered UKBB individuals who self-reported as White British and have similar genotypic backgrounds based on principal components analysis^58^. To ensure unrelatedness, we identified and excluded third-degree relatives or closer via kinship estimation, retaining one individual per related group^58,59^. This yielded a cohort of 337,535 unrelated White British individuals, of which 29,661 have both genetic and cardiac MRI data. We randomly split this data (29,661) into training (15,000), validation (5,000), and test (9,661) sets.

### Genotyping and quality control

For the 29,661 individuals in the study cohort, we leveraged genotype data from approximately 15 million imputed autosomal SNVs. These variants were imputed from 805,426 directly assayed SNVs (obtained by the UKBB from one of two similar Affymetrix arrays) using the Haplotype Reference Consortium and UK10K reference panels^58^. Imputed variants were subject to several quality-control filters, including outlier-based filtration on effects due to batch, plate, sex, array, and discordance across control replicates. We excluded variants due to extreme heterozygosity, missingness, minor allele frequency (< 10^-4^), Hardy-Weinberg equilibrium (< 10^-10^), and poor imputation quality (< 0.9)^58,59^.

### Quantification of left ventricular hypertrophy

We retrieved cardiac MRI images from 44,503 UKBB participants and followed the method described by Bai et al.^20^. Briefly, a fully convolutional network^20^ was trained on 4,875 subjects with 93,500 pixelwise segmentations of UKBB short-axis cardiac MRI multi-slice images generated manually with quality control checks for inter-operator consistency^60^. The cardiac MRI image resolution was 1.8 x 1.8 mm^2^, with a slice thickness of 8.0 mm and a 2.0 mm gap, typically consisting of 10 slices. Each slice was converted to an image and cropped to a 192 x 192 square, and measurements were 0-1 normalized. The network architecture employed multiple convolutional layers to learn image features across five resolution scales. Each scale involved two or three convolutions with kernel size 3 x 3 and stride 1 or 2 (2 appearing every 2 or 3 layers), followed by batch normalization and ReLU transformation. Feature maps from the five scales were upsampled back to the original resolution, combined into a multi-scale feature map, and processed through three additional convolutional layers with kernel size 1 x 1, followed by a softmax function to predict the segmentation label for each pixel. Notably, the pixelwise annotations used for training and evaluation were hand-segmented and validated by a human expert. The model demonstrated strong concordance with human-generated gold standard in UKBB datasets^20^. To our knowledge, this is the only published model trained in the UKBB on gold standard labels. We thus applied this trained deep learning model to our dataset of 44,503 cardiac MRIs, obtaining segmentation of the LV cavity and myocardium from each short axis frame. After quality control checks^20^, 44,219 segmentations were retained. LV myocardium volume was calculated by integrating segmented areas over slices, converted to left ventricular mass (LVM) using a standard density estimate of 1.05 g/mL^61^, and then normalized by an estimate of body surface area using the Du Bois formula^62^ to obtain LVMi. From the 44,219 segmentations, we focused our analysis on LVMi measurements for 29,661 unrelated White British individuals, using data from their most recent imaging visit if multiple imaging visits were recorded.

### Lo-siRF step 1: Dimension reduction of variants via genome-wide association studies

As the first step in lo-siRF, we performed a genome-wide association study (GWAS) on rank-based inverse normal-transformed LVMi from the training dataset using PLINK^26^ and BOLT-LMM^27^. Similar to typical screening in fine-mapping^63^ and other tree-based epistasis detection methods^64,65^, this step reduces over 15 million SNVs to a more computationally-feasible size (Fig. 2b). Since BOLT-LMM and PLINK use different statistical models, we employed both to mitigate model dependence due to this arbitrary choice. Specifically, we performed two GWAS runs: one using a linear regression model implemented via ‘glm’ in PLINK^66^ and another with BOLT-LMM^27^, a fast Bayesian-based linear mixed model. Each GWAS was adjusted for the first five principal components of ancestry, sex, age, height, and body weight. SNVs were ranked by *p*-value for each GWAS run separately, and the top 1000 from each run were combined, yielding 1405 GWAS-filtered SNVs for downstream lo-siRF analysis. PLINK and BOLT-LMM results are provided in Supplementary Data 1, and the 1405 GWAS-filtered SNVs are listed in Supplementary Data 2. These 1405 SNVs strictly contain SNVs that passed the genome-wide significance threshold (*p* = 5E-8).

### Lo-siRF step 2: Binarization of the left ventricular mass phenotype

Next, we partitioned the raw (continuous) LVMi phenotype into low, middle, and high groups before fitting signed iterative random forest (Fig. 2c). Specifically, for a given threshold *x*, we binned individuals within the top and bottom *x*% of LVMi values into high and low LVMi classes, respectively, while omitting individuals in the middle quantile range. Given the sex-specific variation of LVMi (Supplementary Note 1), this partitioning was performed separately for males and females, with cutoffs per sex and binarization threshold detailed in Supplementary Table 2. This binarization step simplifies the original low-signal regression problem, predicting continuous LVMi, into a relatively easier binary classification problem, distinguishing individuals with very high versus very low LVMi. The decision to use this approach was driven by the observation that regression models yielded validation *R^2^* values below 0 (Supplementary Table 3), suggesting poor relevance to reality. The PCS framework for veridical data science^19^ advocates that a model should fit the data well, as measured by prediction accuracy, before trusting any of its interpretations. Since the threshold choice is arbitrary, we ran the remainder of the lo-siRF pipeline using three binarization thresholds (15%, 20%, 25%) to balance prediction signal improvement and data lost. Results stable across all binarization thresholds were aggregated (see Methods, *Lo-siRF step 4.4: Ranking genetic loci and interactions between loci*).

### Lo-siRF step 3: Prediction

#### Lo-siRF step 3.1: Fitting signed iterative random forest on the binarized LV mass index phenotype

For each binarization threshold, we trained a signed iterative random forest (siRF) model^17^ using the 1405 GWAS-filtered SNVs to predict the binarized LVMi phenotype and identify candidate interactions (Fig. 2d). siRF iteratively grows a sequence of feature-weighted random forests, re-weighting features in each iteration proportional to their feature importance from the previous iteration to stabilize the decision paths. If the stabilized forest provides reasonable prediction performance (see Methods, *Lo-siRF step 3.2: Prediction check*), siRF leverages random intersection trees (RIT)^67^ to identify sets of features that frequently co-occur along decision paths. These co-occurring features are more likely to interact and are outputted as candidate interactions by siRF. siRF is well-suited for prioritizing epistatic interactions because (1) it efficiently searches for nonlinear higher-order interactions at a computational cost similar to a traditional random forest and (2) its decision tree thresholding mimics the switch-like behavior observed in biomolecular interactions^53^. Improved upon its predecessor, iterative random forests^16^, siRF also tracks the sign of features^17^. In brief, a signed feature *X^−^* (or *X^+^*) indicates a decision rule of *X* < *t* (or *X > t*) for some threshold *t*. In SNV data, *SNV^+^*typically represents a heterozygous or homozygous mutation, while *SNV^−^* represents no mutation at the locus. We trained siRF using the iRF2.0 R package with the following hyperparameters: number of iterations = 3, number of trees = 500, number of bootstrap replicates = 50, depth of RIT = 3, number of RIT = 500, number of children in RIT = 5, and minimum node size in RIT = 1. Hyperparameter tuning was not performed, since siRF is robust to different choices of hyperparameters^17^. We trained siRF using 10,000 randomly sampled training samples (from 15,000 total), with the remaining 5,000 training samples reserved for selecting genetic loci for the permutation test (see Methods, *Lo-siRF step 4.3: Permutation test for difference in local stability importance scores*).

#### Lo-siRF step 3.2: Prediction check

Per the PCS framework for veridical data science^19^, we assessed the validation prediction accuracy of siRF (Fig. 2d) to ensure it captures biologically-relevant phenotypic signals rather than simply noise before interpreting the model. We compared siRF to several machine learning prediction methods: *L_1_*- regularized (LASSO)^68^ and *L_2_*-regularized (ridge)^69^ logistic regression, random forests^70^, support vector machines^71^, a multilayer perceptron^72^ (fully-connected feedforward neural network with one hidden layer and ReLU activations), and AutoGluon TabularPredictor^73^ (an auto machine learning framework that ensembles multiple models, including neural networks, LightGBM, boosted trees, random forests, and k-nearest neighbors, by stacking them in multiple layers). Implementation and hyperparameters (tuned via 5-fold cross-validation) are detailed in Supplementary Table 6. We also compared siRF to a basic polygenic risk score, constructed using PLINK with lead SNVs from the LVMi PLINK GWAS identified by FUMA at the suggestive significance threshold of 1E-5 (Supplementary Data 1). Logistic regression was performed using this polygenic risk score as a predictor for binarized LVMi. Prediction performance for each of these methods was assessed using classification accuracy, area under the receiver operator curve (AUROC), and area under the precision-recall curve (AUPRC). Though siRF’s prediction power was modest (∼55% balanced classification accuracy, ∼0.58 AUROC, ∼0.57 AUPRC), it consistently outperformed random guessing (i.e., >50% balanced classification accuracy and >0.5 AUROC/AUPRC, which is not guaranteed given the high phenotypic diversity of LVMi) and exceeded all other prediction methods across almost all binarization thresholds and evaluation metrics (Supplementary Table 4). We thus deemed that the siRF fit for LVMi passed the prediction check.

### Lo-siRF step 4: Prioritization

To interpret the siRF fit, we developed a novel stability-driven importance score to prioritize genetic loci and their interactions for follow-up experimental validation (Fig. 2e). Our proposed importance score aggregates weak, unstable variant-level importances into stronger, more stable locus-level importances by: (1) assigning each variant to a genetic locus, (2) evaluating the local (or per-individual) importance of each locus or locus-locus interaction in the siRF fit via a stability-driven measure, and (3) conducting a permutation test to summarize the importance of the locus or interaction across all individuals. We detail each step next.

#### Lo-siRF step 4.1: Aggregation of SNVs into loci

We aggregated SNVs into genetic loci based on genomic proximity, using ANNOVAR^74^ and hg19 refSeq Gene annotations. Each SNV was assigned to a single locus, with a default 1 kb maximum distance from gene boundaries. In lo-siRF, a genetic locus is a non-overlapping group of SNVs, and a signed genetic locus consists of signed SNVs (i.e., *Locus^+^* consists of *SNV ^+^, …, SNV ^+^*, while *Locus*⎯ consists of *SNV_1_*⎯*, …, SNV_p_*⎯).

#### Lo-siRF step 4.2: Local stability importance score

We next measured the importance of a genetic locus or locus-locus interaction based on their stability, defined as frequency of occurrence within the siRF fit (i.e., how often SNVs from a locus or interaction were split upon in the fitted forest). Since raw occurrence frequencies inherently bias towards larger loci containing more SNVs, we developed a *local* (or per-individual) *stability importance (LSI) score*, which quantifies the importance of a signed locus or interaction for predicting each individual’s response. Let *G* = {*g*_1_, …, *g_k_*} denote a signed order-*K* interaction involving the signed loci *g*_1_, …, *g_k_*, and let v_1_^(*j*)^, …, *v_pj_*^(*j*)^ denote the signed SNVs within locus *G*. Given a forest T, a signed interaction c, and an individual i, the *LSI score* is defined as:

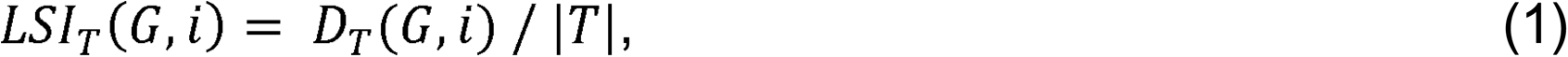

where |*T*| is the number of trees in the forest *T*, and *D_T_*(*G*, *i*) is the number of decision paths in the forest *T* satisfying two criteria: (1) individual *i* appears in its terminal node and (2) for each *j* = 1, …, *K*, there exists an *l* ∈ {1, …, *p_j_*} such that *v_i_*^(*j*)^ was used in a decision split along the path (Extended Data Fig. 3a). Thus, *LSI_T_*(*G*, *i*) is the proportion of trees in the forest *T* where at least one signed variant from each signed locus in c was used to predict individual *i*. A high *LSI score* indicates that the interaction c was frequently used to predict individual i’s response and is an important interaction for individual *i*. Since a single locus can be viewed as an order-1 interaction, the *LSI score* also applies to individual loci for assessing their marginal importance.

#### Lo-siRF step 4.3: Permutation test for difference in local stability importance scores

After computing LSI scores for all individuals, we performed a two-sample permutation test (Extended Data Fig. 3a) to assess whether the LSI scores for a given signed locus or interaction, *G*, differ between individuals with high and low LVMi (conditional on the rest of the fitted forest). This permutation test tests the null hypothesis *L = H* versus the alternative hypothesis *L* ≠ *H*, where *L* and *H* are the LSI distributions for low and high LVMi individuals, respectively. A small nominal *p*-value indicates that *G* can differentiate between high versus low LVMi individuals, suggesting an important risk locus or interaction for LVMi in the fitted siRF. We performed this permutation test using 10,000 permutations, the difference in means as the test statistic, and the 5,000 validation samples. To enhance reliability of our findings, we followed the PCS framework and conservatively tested a subset of genetic loci and interactions that passed predictive and stability checks. Namely, we tested:

a. The 25 genetic loci with the highest average LSI scores across the 5,000 validation samples, which were set-aside from the 15,000 training samples and not used in fitting siRF (see Methods, *Lo-siRF step 3.1: Fitting signed iterative random forest on the binarized LV mass index phenotype*);
b. The signed interactions between loci that were stably identified across 50 siRF bootstrap replicates.

RIT searches within siRF were performed at the locus-level (using the ‘varnames.grp” argument when running siRF in R). We defined an interaction as “stable” if it passed the following siRF stability criteria (Supplementary Table 7):

- Stability score > 0.5 (interaction appears frequently in siRF);
- Stability score for mean increase in precision > 0 (interaction is predictive),
- Stability score for feature selection dependence > 0 (filters out additive interactions)^16,17^.

Given the challenges of analyzing low-signal data, these nominal permutation *p*-values were used primarily to rank candidate loci and interactions rather than as formal tests of statistical significance, which relies heavily on untestable model assumptions that often do not hold in practice.*Lo-siRF step*

### 4.4: Ranking genetic loci and interactions between loci

As a final stability check, we recommended only those signed loci and interactions that underwent the permutation test and consistently yielded a nominal *p*-value < 0.1 across all three binarization runs.

These stably-identified signed loci and interactions were ranked by their mean nominal *p*-value, averaged across the three binarization thresholds (Supplementary Table 5). To prioritize candidates for experimental validation, if both the + and - version of a signed locus (or interaction) appeared, the final ranking was based on the smaller of the two mean nominal *p*-values (Table 1). Nominal *p*-values for all permutation tests, including those for loci and interactions that were unstable across binarization thresholds, are provided in Supplementary Note 1.

### Lo-siRF: PCS documentation, additional stability analyses, and simulations

We acknowledge that many human judgment calls were inevitably made throughout our veridical machine learning pipeline and that alternative choices could have been made (e.g., different dimension reduction techniques, binarization procedures, and prediction models). We provide extensive documentation, discussion, and justification for these decisions in Supplementary Note 1. We also performed stability analyses^19^ to ensure that our findings are stable and robust to these human judgment calls. We lastly conducted a simulation study using the simChef R package^75^, showing that lo-siRF produces calibrated *p*-values under a null response model and investigated the performance of lo-siRF under both a marginal effect and interaction effect simulation model. Supplementary Note 1 is an HTML document, which can be downloaded and displayed in a browser or found at https://yu-group.github.io/epistasis-cardiac-hypertrophy/.

### Non-hypertensive cohort analysis

We defined hypertensive individuals as anyone with self-reported hypertension, a doctor-diagnosed high blood pressure, or any ICD10 billing code diagnosis in I10-I16. Out of the 29,661 UKBB participants in the original lo-siRF analysis, 7,371 had hypertension, leaving 22,290 for the non-hypertensive analysis. Using the same 1405 GWAS-filtered SNVs as in the original lo-siRF analysis, we repeated steps 2-4 of lo-siRF exclusively in the non-hypertension cohort. Additionally, We assessed the marginal effect of each of the 1405 GWAS-filtered SNV on hypertension by fitting logistic regression models that regressed hypertension status (binary) on each SNV while adjusting for the first five principal components of ancestry, sex, age, height, and body weight. Detailed discussion of the non-hypertension analysis results can be found in Supplementary Note 1.

### Functional interpretation of lo-siRF-prioritized variants

#### Functional interpretation step 1: Extraction of candidate SNVs and LD structures

We used the SNP2GENE function in FUMA (v1.5.4)^33^ to incorporate LD structure and prioritize candidate genes. Taking GWAS summary statistics from PLINK^26^ and BOLT-LMM^27^ as an input, we submitted the 283 lo-siRF-prioritized SNVs into SNP2GENE as predefined SNVs. This allows SNP2GENE to define LD blocks for each SNV and include both the input 283 SNVs and those in LD with them for further annotations. Using the default *r*^2^ threshold (0.6), FUMA defined all 283 SNVs as independent significant SNVs, since any two of them are in LD with each other at *r*^2^ < 0.6. To match the population used in lo-siRF prioritization, we selected the UKBB release 2b reference panel for British and European subjects to compute *r*^2^ and minor allele frequencies. FUMA identified 572 candidate SNVs in strong LD (*r*^2^ < 0.6) with the 283 independent significant SNVs, extracted from both the input GWAS (*p* < 0.05) and the reference panel. Each candidate SNV was then assigned to one of the six lo-siRF-identified loci (Table 1) based on the independent significant SNV (from the 283 SNVs), with which it had the highest *r^2^*value. A combination of the 283 independent significant SNVs and the 572 candidate SNVs (details in Supplementary Data 4) was defined as the *lo-siRF-prioritized SNV set* and used to generate the lo-siRF-prioritized gene list (Fig. 4a) for enrichment analysis (Fig. 4c-4e). For comparison, we also uploaded all 1405 GWAS-filtered SNVs (Supplementary Data 2) as the predefined SNVs in a separate SNP2GENE job. Using the same approach and parameter settings, FUMA identified 929 independent significant SNVs within the input set and 5771 candidate SNVs in LD with the 929 SNVs. The combined set of 929 independent significant SNVs and 5771 candidate SNVs was defined as the *reference SNV set*. Unlike the *lo-siRF-prioritized SNV set*, this *reference SNV set* was derived purely from GWAS prioritization, without considering epistatic effects.

#### Functional interpretation step 2: ANNOVAR enrichment test

We performed ANNOVAR enrichment test on the *lo-siRF-prioritized SNV set* against the selected reference panel in FUMA (see previous step). SNP2GENE generated unique ANNOVAR^74^ annotations for all identified SNVs. The enrichment score for a given annotation in each lo-siRF-prioritized locus (Fig. 3b) was computed as the proportion of SNVs with that annotation in the locus, divided by the proportion of SNVs with the same annotation in the reference panel. To compute the enrichment *p*-value for the *i^th^*ANNOVAR annotation in the *j^th^* lo-siRF-prioritized locus, we performed a two-sided Fisher’s exact test on the 2-by-2 contingency table containing (Supplementary Data 5):

· *n_j_*(*i*) = the number of SNVs with the *i^th^* annotation in the *j^th^* lo-siRF-prioritized locus,
· ∑*_t_n_j_*(t) -*n_j_*(*i*), with ∑*_t_n_j_* (t) being the summation of *n_j_* (*i*) for all available annotations in the *j^th^* lo-siRF-prioritized locus,
· N(*i*) -*n_j_*(*i*), with N(*i*) being the number of SNVs with the *i^th^* annotation in the reference panel,
· ∑*_t_ N*(*t*) - ∑*_t_ n_j_*(*t*) - *n_j_*(ii), where ∑*_t_ N*(*t*) is the summation of *N*(*i*) for all available annotations in the reference panel.

#### Functional interpretation step 3: Functional annotations

In addition to ANNOVAR annotations, FUMA annotated all identified SNVs for potential regulatory functions (core-15 chromatin state prediction and RegulomeDB score) and deleterious effects (CADD score). The core-15 chromatin state was annotated to all SNVs of interest by ChromHMM^45^ derived from 5 chromatin markers (H3K4me3, H3K4me1, H3K36me3, H3K27me3, and H3K9me3) for four relevant tissue/cell types, including left ventricle (E095), right ventricle (E105), right atrium (E104), and fetal heart (E083) (Fig. 3a, circle 7). Data and a description of the core-15 chromatin state model can be found at https://egg2.wustl.edu/roadmap/web_portal/chr_state_learning.html. RegulomeDB^33,35^ annotations guide interpretation of regulatory variants through a seven-level categorical score, with category 1 (including 6 subcategories ranging from 1a to 1f) indicating the strongest functional evidence.

Because the RegulomeDB database (v1.1) used in FUMA has not been updated, we queried all SNVs identified by lo-siRF and FUMA in the RegulomeDB database v2.2 (https://regulomedb.org/regulome-search). Deleteriousness annotations were obtained from CADD database (v1.4)^36^ by matching chromosome, position, reference, and alternative alleles of all SNVs. High CADD scores indicate highly deleterious effects of a given variant, with a threshold score of 12.37^36^. We also retrieved eQTL and sQTL data for all 283 independent significant SNVs and 572 candidate SNVs from GTEx v8^37^. All functional annotation results are summarized in Supplementary Data 4.

#### Functional interpretation step 4: Functional gene mapping

We performed three functional gene mapping strategies in SNP2GENE – positional, eQTL, and 3D chromatin interaction mapping – using the *lo-siRF-prioritized* and *reference SNV set* (see Methods, *Functional interpretation step 1: Extraction of candidate SNVs and LD structures*). For positional mapping^33,35^, a default 10 kb maximum distance was used between SNVs and genes. For eQTL mapping, we used *cis*-eQTL data of heart left ventricle, atrial appendage, and muscle skeletal tissues from GTEx v8^37^, with significant SNV-gene pairs (FDR < 0.05, *p* < 1E-3). For 3D chromatin interaction mapping, Hi-C data of left ventricle tissue (GSE87112) with a default FDR < 1E-6 threshold was used.

A default promoter region window was defined as 250 bp upstream and 500 bp downstream of TSS^33,35^. Using these strategies, we mapped the lo-siRF-prioritized SNV set to 20 HGNC-recognizable protein-coding genes (Fig. 4a). Each gene was functionally linked to a specific lo-siRF-prioritized locus (Table 1), to which the highest proportion of SNVs mapped to that gene were assigned. A Circos plot (Fig. 3a) was created by TBtools^76^ to visualize the lo-siRF-prioritized epistatic interactions, eQTL SNV-to-gene connections, 3D chromatin interactions, as well as LD structures and the 20 prioritized genes. These 20 genes were submitted to GENE2FUNC in FUMA, yielding GTEx gene expression data for 19 genes across multiple tissue types (Fig. 4b). In a separate SNP2GENE job, we mapped the reference SNV set to 382 HGNC-approved genes using the same approach, and used both gene sets for enrichment analysis.

### Gene ontology and pathway enrichment analysis

To assess differential GO and pathway co-association among lo-siRF-prioritized genes versus their counterparts deprioritized by lo-siRF, we performed an integrative GO and pathway enrichment analysis followed by an exhaustive permutation of co-association scores for all possible gene-gene combinations within the aforementioned 382 HGNC-approved genes (see Methods, *Functional interpretation step 4: Functional gene mapping*).

To improve GO and pathway prioritization, we adopted the concept from Enrichr-KG^77^ and ChEA3^44^ to assess enrichment results across libraries and domains of knowledge as an integrated network of genes and their annotations. We queried the 382 HGNC-approved genes against various prior-knowledge gene set libraries in Enrichr^43^ (https://maayanlab.cloud/Enrichr/), including GO biological process^78,79^, GO molecular function^78,79^, MGI Mammalian Phenotypes^80^, Reactome pathways^81^, and KEGG pathways^82^. This allowed us to search for a union of enriched GO and pathway terms and their correspondingly annotated gene sets, from which we built a co-association network, where nodes are either the enriched terms or genes. To measure the degree of co-association to specific GO or pathway terms for two given genes, we computed a co-association score for each of the 72,771 possible gene pairs (from the 382 queried genes). The co-association score was calculated as:

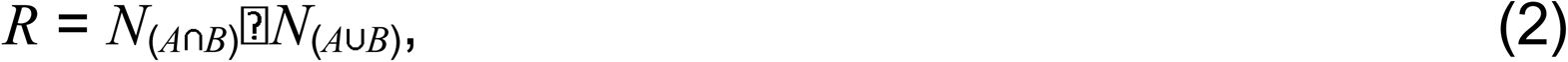

where *N*_(*A*_⋂*_B_*_)_ denotes the number of GO or pathway terms significantly enriched for both *gene A* and *gene B* in a gene pair, and *N*_(*A*_⋃*_B_*_)_ is the number of GO or pathway terms enriched for either *gene A* or *gene B*. For cases where *N*_(*A*_⋃*_B_*_)_ = 0, *R* = 0. Note that the 20 genes mapped from lo-siRF-prioritized SNVs form a subset of the 382 HGNC-approved genes. We compared the co-association scores *R* for the 20 lo-siRF-prioritized genes relative to the full distribution of *R* from an exhaustive permutation of all possible gene pairs within the 382 HGNC-approved genes, ranking gene pairs by two-sided empirical *p*-values (Fig. 4c). Further details are in Supplementary Data 6.

### Transcription factor enrichment analysis

Owing to the limitations and biases of various specific assays, we performed an integrative transcription factor (TF) enrichment analysis against the following 9 annotated gene set libraries in ChEA3^44^ and Enrichr^43^ from distinct sources:

1. Putative TF target gene sets determined by ChIP-seq experiments from ENCODE^83^;
2. Putative TF target gene sets determined by ChIP-seq experiments from ReMap^84^;
3. Putative TF target gene sets determined by ChIP-seq experiments from individual publications^44^;
4. TF co-expression with other genes based on RNA-seq data from GTEx^37^;
5. TF co-expression with other genes based on RNA-seq data from ARCHS4^85^;
6. Single TF perturbations followed by gene differential expression^43^;
7. Putative target gene sets determined by scanning PWMs from JASPAR^86^ and TRANSFAC^87^ at promoter regions of all human genes;
8. Gene sets predicted by transcriptional regulatory relationships unraveled by sentence-based text-mining (TRRUST)^88^;
9. Top co-occurring genes with TFs in a large number of Enrichr queries^44^.

Of the mentioned 9 gene set libraries, libraries 1, 3, and 5 were assembled by combining gene set libraries downloaded from both ChEA3^44^ and Enrichr^43^. Libraries 2, 4, and 9 were downloaded from ChEA3^44^. Libraries 6, 7, and 8 were downloaded from Enrichr^43^. Following the ChEA3^44^ integration method, when multiple gene sets were annotated to the same TF, the gene set with the lowest two-sided Fisher’s exact test (FET) *p*-value was used for each library. As mentioned previously, we have mapped lo-siRF-prioritized SNVs and all GWAS-filtered SNVs to two gene sets: a lo-siRF-prioritized set (20 HGNC-recognizable genes) and a reference set (382 HGNC-recognizable genes). The 362 genes in the reference set complementary to the 20 lo-siRF-prioritized genes served as the lo-siRF-deprioritized gene set. We then performed separate enrichment analysis on the lo-siRF-prioritized (20 genes) and lo-siRF-deprioritized (362 genes) sets across the 9 gene set libraries. TF enrichment significance was ranked for each library by FET *p*-values, with ties assigned the same integer rank.

Each TF’s integer rank was scaled by dividing by the maximum rank within its respective library. We then integrated the 9 sets of TF rankings and re-ordered the TFs by two sequential criteria: (1) the number of libraries showing significant enrichment (FET *p* < 0.05) and (2) the mean scaled rank across all libraries containing that TF. This approach prioritized distinct sets of TFs for the lo-siRF-prioritized genes (Fig. 4d, top) and lo-siRF-deprioritized genes (Fig. 4d, bottom).

To assess TF co-association^43,44^ between lo-siRF-prioritized genes, we computed TF co-association scores using Equation (2), where *N*_(*A*_⋂*_B_*_)_ and *N*_(*A*_⋃*_B_*_)_ denote the number of enriched TF terms instead of GO/pathway terms (see Methods, *Gene ontology and pathway enrichment analysis*). TF co-associations between lo-siRF-prioritized genes were ranked by empirical *p*-values (Fig. 4e) from an exhaustive permutation of all 72,771 possible gene pairs from the 382 genes. Further details are in Supplementary Data 7.

### Disease-state-specific gene co-expression network analysis

To evaluate gene connectivity and its role in myocardial transition from healthy to failing states, we analyzed the topological structure of gene co-expression networks in human heart tissues (Fig. 5). To construct gene co-expression networks, cardiac tissue samples from 177 failing hearts and 136 donor, non-failing (control) hearts were collected from operating rooms and remote locations for RNA expression measurements. We performed weighted gene co-expression network analysis (WGCNA) on the covariate-corrected RNA microarray data for the control and heart failure networks separately^47^ (Fig. 5a). Data for these networks is available at https://doi.org/10.5281/zenodo.2600420. To evaluate the degree of connectivity between correlating genes in each of the networks, we compared edge weights between lo-siRF-prioritized genes relative to the distribution of all possible gene pairs (Fig. 5b and 5c). We also evaluated the difference of edge weights (Z-score normalized) between the control and heart failure networks to understand how these gene-gene connectivities change between non-failing and failing hearts (Fig. 5d). Two-tailed empirical *p*-values represent the proportion of absolute difference in edge weights of all gene pairs that exceed the absolute difference score for gene pairs of interest. Additionally, we compared the structure of modules derived from dendrograms on the WGCNA control and heart failure networks (Extended Data Fig. 6). Modules were labeled according to Reactome enrichment analysis of genes within each module. Full gene module descriptions and Benjamini-Hochberg-adjusted enrichment *p*-values are available in Supplementary Data 5 and 6 of Cordero et al.^47^.

### Induced pluripotent stem cell cardiomyocytes differentiation

The studied patient-specific human induced pluripotent stem cells (hiPSCs) were derived from a 45-year-old female proband with a heterozygous *MYH7*-R403Q mutation. Derivation and maintenance of hiPSC lines were performed following Dainis et al.^24^. Briefly, hiPSCs were maintained in MTeSR (StemCell Technologies) and split at a low density (1:12) onto fresh 1:200 matrigel-coated 12 well plates. Following the split, cells were left in MTeSR media supplemented with 1 μM Thiazovivin. The hiPSCs were maintained in MTeSR until cells reached 90% confluency, which began Day 0 of the cardiomyocyte differentiation protocol. Cardiomyocytes were differentiated from hiPSCs using small molecule inhibitors. For Days 0-5, cells were given RPMI 1640 medium + L-glutamine and B27 - insulin. On Days 0 and 1, the media was supplemented with 6 μM of the GSK3β inhibitor, CHIR99021. On Days 2 and 3, the media was supplemented with 5 μM of the Wnt inhibitor, IWR-1. Media was switched to RPMI 1640 medium + L-glutamine and B27 + insulin on Days 6-8. On Days 9-12, cells were maintained in RPMI 1640 medium + L-glutamine - glucose, B27 + insulin, and sodium lactate. On Day 13, cells were detached using Accutase for 7-10 minutes at 37 ℃ and resuspended in neutralizing RPMI 1640 medium + L-glutamine and B27 + insulin. This mixture was centrifuged for 5 minutes at 1000 rpm (103 rcf). The cell pellet was resuspended in 1 μM thiazovivin supplemented RPMI 1640 medium + L-glutamine and B27 + insulin. For the rest of the protocol (Days 14-40), cells were exposed to RPMI 1640 medium + L-glutamine - glucose, B27 + insulin, and sodium lactate. Media changes occurred every other day on Days 14-19 and every three days for Days 20-40. On Day 40, cardiomyocytes reached maturity.

### RNA silencing in induced pluripotent stem cell-derived cardiomyocytes

Mature hiPSC-derived cardiomyocytes were transfected with Silencer Select siRNAs (Thermofisher) using TransIT-TKO Transfection reagent (Mirus Bio). Cells were incubated for 48 hours with 75 nM siRNA treatments. Four wells of cells were transfected with each of the six siRNAs: scramble, *CCDC141* (ID s49797), *IGF1R* (ID s223918), *TTN* (ID s14484), *CCDC141* and *IGF1R*, and *CCDC141* and *TTN*. After 2 days, hiPSC-CMs were collected for RNA extraction.

### RT-qPCR analysis for siRNA gene silencing efficiency

Following cell morphology measurement, all cells for each condition were centrifuged for 5 minutes at 1000 rpm (103 rcf). Cell pellets were frozen at -80 L prior to RNA extraction. RNA was extracted using Trizol reagent for RT-qPCR to confirm gene knockdown occurred. Reverse Transcription of RNA was done using High-Capacity cDNA Reverse Transcription Kit (Thermofisher). qPCR of the single stranded cDNA was performed using TaqMan Fast Advanced MM (Thermofisher) with the following annealing temperatures: 95°C 20” and 40 cycles of 95°C 1” and 60°C 20”. qPCR of the silenced genes was performed using TaqMan® Gene Expression Assays, including *CCDC141* (Hs00892642_m1), *IGF1R* (Hs00609566_m1), and *TTN* (Hs00399225_m1). For gene-silencing efficiency analysis, gene RPLP0 (Hs00420895_gH) was used as a reference gene. Data were analyzed using the delta-delta Ct method.

### Cell sample preparation for cell morphology measurement

Following siRNA treatments, cells were detached for microfluidic single-cell imaging using a mixture of 5 parts Accutase and 1 part TrypLE, treated for 6 minutes at 37 L. Cells were then added to the neutralizing RPMI 1640 medium + L-glutamine and B27 + insulin. These mixtures were centrifuged for 5 minutes at 1000 rpm (103 rcf). For each gene-silencing condition, four wells of cells were resuspended in 4 mL of the MEM medium, which is composed of MEM (HBSS balanced) medium, 10% FBS, and 1% Pen Strep (Gibco). Cells were filtered with 100 μm strainers (Corning) before adding into the microfluidic devices.

### Microfluidic inertial focusing device

We adopted the “Dean flow focusing” concept by Guan et al.^22^ and developed a spiral inertial microfluidics system to focus randomly suspended cells into single streams based on cell size for high- resolution, high-throughput single-cell imaging. The microfluidic device (Extended Data Fig. 7) contains a five-loop spiral microchannel with an expanding radius (3.3 mm to 7.05 mm) and a cross-section with a slanted ceiling (dimensions and fabrication details in Supplementary Note 3). This geometry induces strong Dean vortices in the outer half of the channel cross-section, optimizing size-based separation and cell focusing. Two inlets at the spiral center are used to introduce cell suspensions and sheath flow of fresh medium, while the top and bottom outlets enable high-throughput imaging of hypertrophic and non-hypertrophic cardiomyocytes (Extended Data Fig. 9).

### High-throughput single-cell imaging

Before each experiment, microchannels were flushed with 3 mL of the MEM medium. Prepared cell samples and fresh MEM medium were loaded into 3 mL syringes, which were connected to the corresponding microchannel inlets using Tygon PVC tubing (McMaster). Both cells and the fresh MEM medium were infused into the microchannel using a Pico Plus Elite syringe pump (Harvard Apparatus) at 1.2 mL/min. Microscope image sequences of cells focused to the top and bottom observation channels were captured using a VEO 710S high-speed camera (Phantom) with a sampling rate of 700 fps and a 5 μsec light exposure.

### Image analysis for cell feature extraction

For each gene-silencing condition in each biological replicate, 21,000 images were processed to extract cell morphology features. To analyze cell size and shape changes induced by gene silencing, we developed a MATLAB-based image analysis pipeline with three major steps: image preparation, feature extraction, and post-processing (Fig. 6c). In step one, image sequences were background-corrected by subtracting an automatically generated background image, where each pixel’s value was computed as the mode intensity value of that pixel location across the entire image sequence. After illumination correction, step two detects cell edges using local maxima of the bright-field intensity gradient, following which the program closes edge gaps, removes border-adjacent cells, filters small features (noise), and fills holes to generate binary images with centroid positions for each cell. A double-counting filter was used to exclude repeated measurements of stuck cells collected in the same location and cell size range using a Gaussian kernel density method (bandwidth = 0.09). Binary images passing the double-counting filter were used to create cell outline coordinates (X, Y), which leads to various morphological features, including cell area (2D integration of the cell outline), diameter 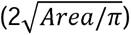, solidity (Cell area/Convex hull area), roundness error (standard deviation/mean of radii from centroid), circularity (4*π* x *Area*/*Perimeter*^2^), and intensity spatial relationship enclosed within the cell boundary.

The 2D intensity distributions within cell outlines were used to derive peak locations and count peak numbers, which is a measure of intensity spatial relationship and a gating parameter to remove clumped cells. In the post-processing step, data were filtered using three gating thresholds. To remove large clumps, the peak-solidity filter removes data outside of the polygonal region defined by {(0.9, 0), (0.9, 3.2), (0.934, 8.26), (1, 28), (1, 0)} in the (solidity, peak No.) space. Then, the roundness filter removes cells with weird shapes by excluding data with a roundness error higher than 0.3 or a circularity lower than 0.6. Finally, the small size filter removes cell debris whose major diameter is lower than 15 μm (12 μm) or minor diameter is lower than 12 μm (10 μm) for images photographed at the top (bottom) outlet microchannels. All these filters were manually validated through visual inspection.

### Statistical assessment of gene-silencing effects on single-cell morphology

We conducted various statistical analyses to investigate how and where cell size distributions differ between the gene/gene-pair silenced groups and their respective scrambled control groups. First, we compared differences in median cell size (i.e., diameter in μm) using both a two-sided Mann-Whitney U test (Fig. 7c) and a bootstrap quantile test at the 0.5 quantile level. In accordance with the PCS framework, we used two different tests to ensure that our findings are robust to this arbitrary modeling choice. Median cell size difference was of greater interest than mean size difference due to the heavy right-skewness of cell size distributions. Additionally, we compared differences in upper quantiles (0.6, 0.7, 0.8, and 0.9 quantile levels) of size distributions between gene/gene-pair silenced cells and their scrambled controls using a bootstrap quantile test (Extended Data Fig. 8). Identifying cell size differences at these upper quantiles, which highlight larger, hypertrophic cells, is particularly relevant for understanding the pathologic phenotype of cardiac hypertrophy and its clinical implications. All tests were performed separately for each experimental batch, and the most conservative *p*-value (maximum across batches) was reported. Similar analyses were conducted for assessing differences in morphological features (i.e., cell roundness error and normalized peak number).

### Statistical assessment of non-additivity in gene-silencing effects

We next assessed whether gene pairs (*CCDC141*-*TTN* and *CCDC141*-*IGF1R*) affect cell size additively or non-additively (i.e., through epistasis, Fig. 7d). To assess this (non-)additivity for a given gene pair, say *gene 1* and *gene 2*, we fit the following quantile regression:

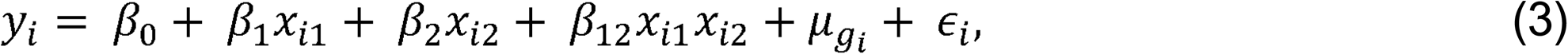

where *y_i_* is the diameter (μm) of cell *i, x_i_*_1_ and *x_i_*_2_ indicate whether *gene 1* and *gene 2* were silenced in cell *i*, respectively, *µ_gi_* accounts for batch effects (with *g_i_* denoting the batch identifier for cell *i*), and *∈_i_* is the random error or noise term for cell *i*. This regression was fitted using scrambled control cells (*x_i_*_1_ = *x_i_*_2_ = 0), *gene 1*-silenced cells (*x_i_*_1_ = 1; *x_i_*_2_ = 0), *gene 2*-silenced cells (*x_i_*_1_ = 0; *x_i_*_2_ = 1), and double-gene-silenced cells (*x_i_*_1_ = *x_i_*_2_ = 1). Under this regression model, we tested the null hypothesis *β*_12_ =0 versus the alternative hypothesis *β*_12_ *0 via a percentile bootstrap t-test and a traditional t-test (Supplementary Data 8) for varying quantile levels (0.5, 0.6, 0.7, 0.8, 0.9). A small *p*-value suggests non-additive epistatic interaction. Again, two different tests were performed to ensure robustness against modeling assumptions. To further bolster the robustness of our conclusions, we repeated this assessment of epistasis using a rank-based inverse normal-transformed cell diameter as the response *y* in an ordinary linear regression model, yielding similar *p*-values and directions of non-additive interaction effects (Supplementary Data 8). Since gene-silencing efficiency varied across conditions (e.g., silencing *CCDC141* and *TTN* vs only silencing *CCDC141* vs only silencing *TTN*), we restricted the analysis to experimental batches with high gene-silencing efficiencies (>60%) to minimize spurious epistatic signals. See Supplementary Note 5 (available on the website: https://yu-group.github.io/epistasis-cardiac-hypertrophy/simulations_efficiency) for further discussion and simulations. Similar analyses were conducted for assessing differences in morphological features (i.e., cell roundness error and normalized peak number).

## Supporting information

Supplementary webpage 1

Supplementary webpage 2

## Data availability

All genotype and cardiac MRI data used as input to the lo-siRF pipeline are available from the UK Biobank (https://www.ukbiobank.ac.uk/). This work was conducted under the UK Biobank application 22282. Data for the gene co-expression networks from 313 explanted human hearts is available at https://doi.org/10.5281/zenodo.2600420. The RNA-seq data used to establish the healthy and heart failure gene co-expression networks is available in the Gene Expression Omnibus (GEO) under accession number GSE57338. Other databases used in this study include RegulomeDB (v2.2), CADD (v1.4), GTEx (v8), Enrichr (last updated version: June 8, 2023), and ChIP-X Enrichment Analysis Version 3 (ChEA3). All other data have been provided as Tables and Supplementary Data files.

## Code availability

All code for running the custom lo-siRF analysis and analyzing the experimental results can be found on GitHub (https://github.com/Yu-Group/epistasis-cardiac-hypertrophy). This custom lo-siRF analysis was conducted using R v3.6.1, Python v3.6.1, and iRF2.0 (https://github.com/karlkumbier/iRF2.0). The LVMi derivation from cardiac MRI images was computed using the ukbb_cardiac package (https://github.com/baiwenjia/ukbb_cardiac) and Python v3.9. PLINK v1.90b5.3 (https://www.cog-genomics.org/plink/) and BOLT-LMM v2.3.4 (https://alkesgroup.broadinstitute.org/BOLT-LMM/BOLT-LMM_manual.html) were used to perform the GWAS dimension reduction. ANNOVAR (downloaded version: June 8, 2020) (https://annovar.openbioinformatics.org/en/latest/) was used to map each SNV to a genetic locus within lo-siRF. Comparisons to existing methods were performed using the SKAT R package (v2.2.5), mvMAPIT R package (v2.0.3), GSEA v4.3.3 (https://www.gsea-msigdb.org/gsea/index.jsp), and MAGMA (v1.6). Functional annotation and gene mapping were performed using FUMA v1.5.4 (https://fuma.ctglab.nl/), ChromHMM v1.10 Core 15-state (https://egg2.wustl.edu/roadmap/web_portal/chr_state_learning.html), RegulomeDB v2.2 (https://regulomedb.org/regulome-search/), CADD v1.4 (https://cadd.gs.washington.edu/), and GTEx v8 (https://www.gtexportal.org/home/). Circos plot was made using TBtools (v2.210). Gene set enrichment analyses for gene ontology, pathway, and transcription factor were performed using Enrichr (last updated version: June 8, 2023) (https://maayanlab.cloud/Enrichr/), ChEA3 (https://maayanlab.cloud/chea3/), and MATLAB R2019a. Healthy and heart failure network analyses were performed previously in R v3.5.2. Cell boundary extraction and morphology analysis from single cell images was performed using MATLAB R2019a.

## Acknowledgements

The authors would like to acknowledge Dr. David Amar (Stanford University), Dr. Srigokul Upadhyayula (University of California, Berkeley), Dr. Haiyan Huang (University of California, Berkeley), Dr. Ziad Obermeyer (University of California, Berkeley), and Dr. Lorin Crawford (Microsoft Research; Brown University) for their critical comments and discussions on this work, Elmer Enriquez (Stanford nanofabrication facility) for the technical support of microfluidic device fabrication, and Dr. Anna Shcherbina (Stanford University) and Dr. Manuel Rivas (Stanford University) for their technical help with the UK Biobank data. The authors would also like to thank the anonymous reviewers and editor for their invaluable feedback and discussions, which have greatly improved this work. This work was supported by the Chan Zuckerberg Biohub – San Francisco through the Intercampus Research Awards (2019 - 2022) (R.A., J.R.P., J.B.B., A.J.B., E.A.A., B.Y.); the National Institutes of Health (NIH) through grant numbers 1R01HL144843 (E.A.A.), R01GM152718 (B.Y.), K08HL143185 (V.N.P.), R01HL105993 (W.H.W.T., K.B.M., T.P.C., E.A.A.), F32HL160067 (C.S.W.), K08HL167699 (C.S.W.), and L30HL159413 (C.S.W.); the American Heart Association (AHA) through grant numbers 23POST1023278 (Q.W.) and 23CDA1042900 (C.S.W.); the National Science Foundation (NSF) through grants DMS-1613002 (B.Y.), IIS 1741340 (B.Y.), and the Graduate Research Fellowship Program DGE-2146752 (T.M.T); and a Weill Neurohub grant (B.Y.).

## Contributions

Q.W., T.M.T., C.S.W., J.W.H., A.J.B., R.A., J.B.B., J.R.P., V.N.P., B.Y., and E.A.A. conceived and designed research. J.W.H. performed LVMi extraction from cardiac MRI images. T.M.T., A.A., X.L., M.B., and K.K. performed exploratory data investigations leading to development of lo-siRF. T.M.T., A.A., and B.Y. developed the lo-siRF pipeline; T.M.T. performed the lo-siRF analysis. Q.W. and T.M.T. performed the FUMA SNP2GENE process using lo-siRF-prioritized SNVs and GWAS-filtered SNVs.

Q.W. evaluated functional annotation results from FUMA, performed ANNOVAR enrichment test for each lo-siRF loci and functional gene mapping. Q.W. performed integrative biological enrichment analyses and evaluated the co-associations between lo-siRF hypothesized gene-gene interactions and the enriched GOs, pathways, and TFs. E.T.C., C.S.M., W.H.W.T., K.B.M., T.P.C., V.N.P., and E.A.A. contributed to the data collection and construction of WGCNA healthy and heart failure coexpression networks. Q.W. and E.T.C. evaluated connectivity differences of epistatic genes hypothesized by lo- siRF between cardiac co-expression networks of failing and non-failing hearts. Q.W. designed and created microfluidic devices; M.Y., S.C.S. and V.N.P. created gene-silenced hiPSC-CM lines; Q.W. and M.Y. performed the microfluidic single cell imaging experiments. Q.W., O.R., and A.M.K. performed single cell image analysis and morphological feature extraction. Q.W., T.M.T., A.M.K., O.R., C.S.W., V.N.P., J.W.H., B.Y., and E.A.A. interpreted results of experiments; Q.W., T.M.T., and A.M.K. prepared figures; Q.W., T.M.T., C.S.W. and J.W.H. drafted manuscript; All authors contributed to editing and revising manuscript.

## Extended Data Figure Legends

**Fig. S1:**
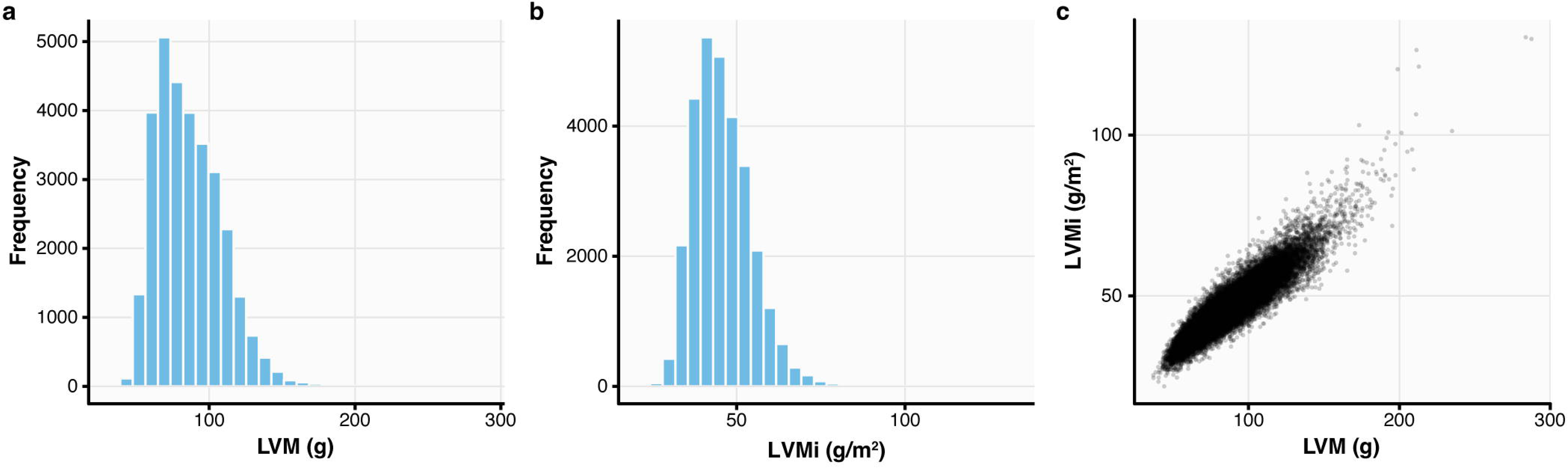
Distribution of LVM and LVMi measurements for 29,661 UK Biobank participants. Left ventricular mass (LVM, **a**) and LVM indexed to body surface area (LVMi, **b**) measurements were extracted from cardiac magnetic resonance imaging for 29,661 unrelated White British individuals via deep learning^20^. **c,** A high Pearson correlation of 0.92 was observed between these LVM and LVMi measurements.

**Fig. S2:**
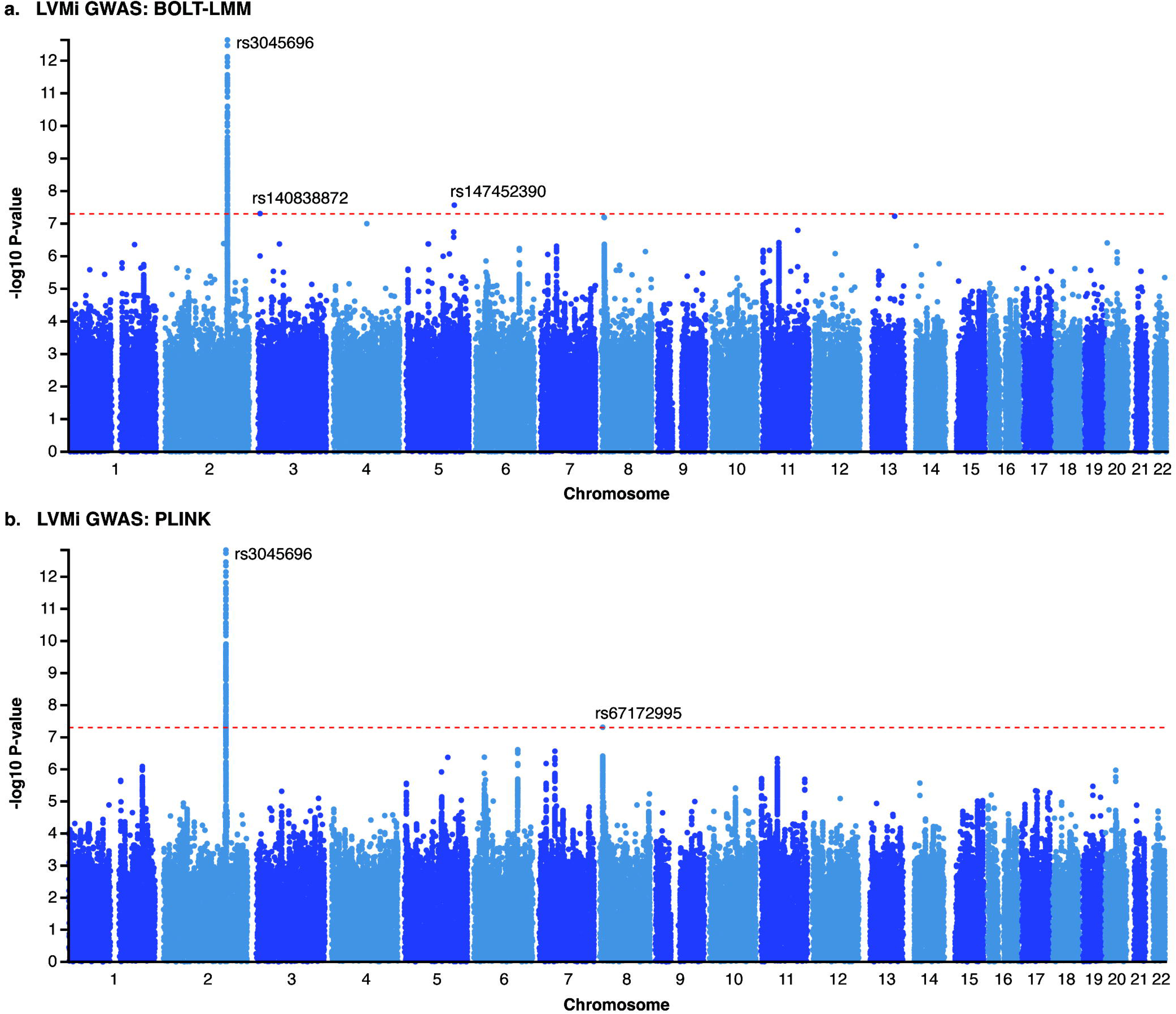
LVMi GWAS using BOLT-LMM and PLINK. Two-sided GWAS *p*-values from BOLT-LMM (**a**) and PLINK (**b**) identified LVMi-associated SNVs, of which the top lead SNV rs3045696 showed the highest significance in both analyses. Other labeled lead SNVs were significant in only one of the two methods. The red dashed line indicates the genome-wide significance threshold (*p* < 5E-8, two-sided). Notably, rs3045696 and rs67172995 were also stably prioritized by lo-siRF as epistasis interactor variants. Further details are provided in Supplementary Data 1.

**Fig. S3:**
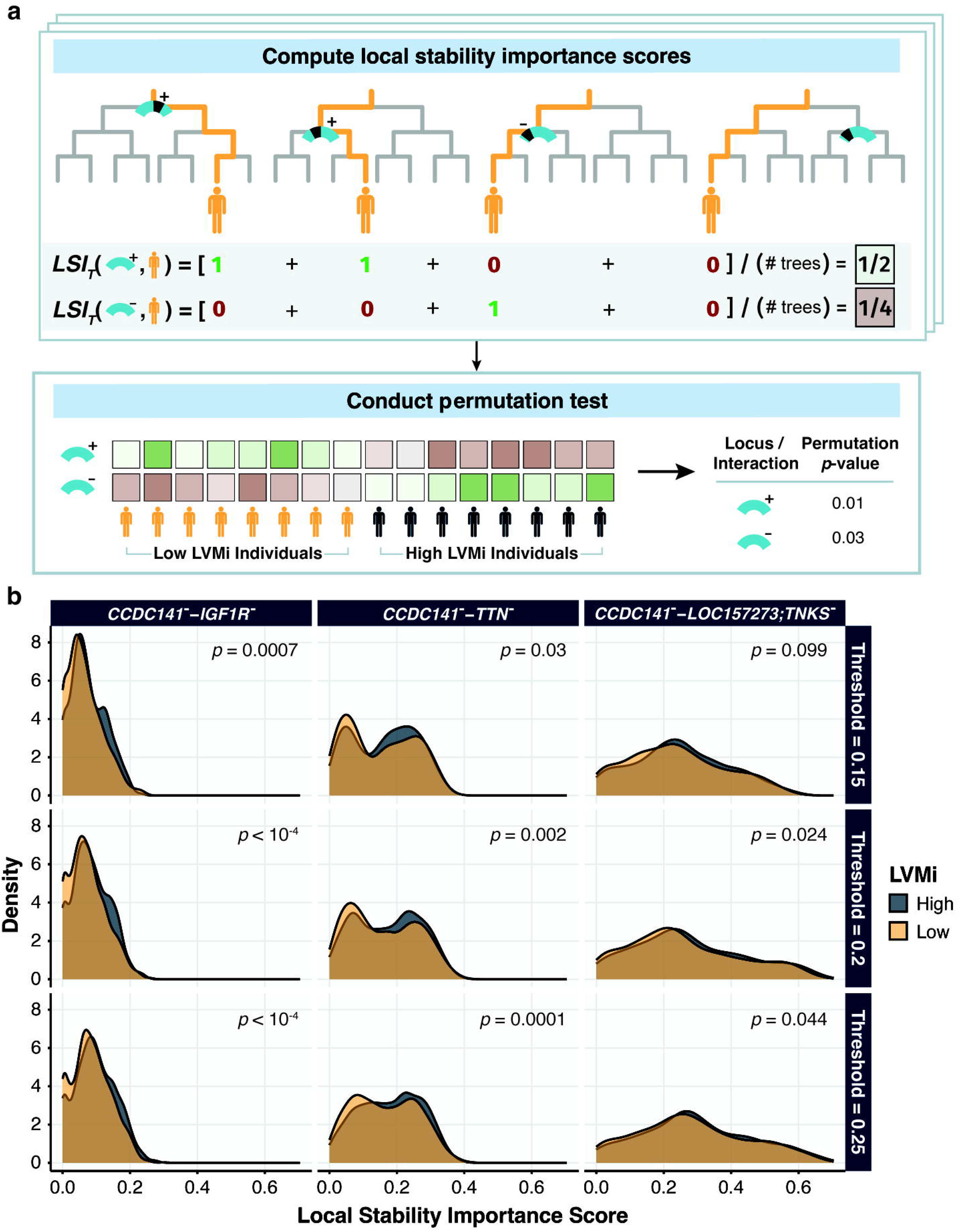
Differences in local stability scores between high and low LV mass highlight the importance of the lo-siRF-prioritized interactions between genetic loci. **a**, Schematic of local stability importance score computation. Given a locus (light blue transcript), the local stability importance score for an individual is defined as the proportion of trees for which at least one SNV (shaded black region) in the locus is used in the individual’s decision path. This computation (top) was performed for each individual (denoted by the stacked boxes). Then, a two-sided permutation test was conducted to assess the difference in these local stability importance scores between the low and high LVMi individuals (bottom). **b**, Differences in the distribution of local stability importance scores suggest that the identified interactions between genetic loci are important for differentiating individuals with high (dark gray) and low (orange) LVMi in the siRF fit. This result, evaluated on the validation data, is stable across the three binarization thresholds and is quantified by a nominal two-sided *p*-value from a permutation test given in the top right corner of each subplot. Given the use of 10,000 permutations, *p*-values cannot be reported with greater precision than *p* = 1E-4.

**Fig. S4:**
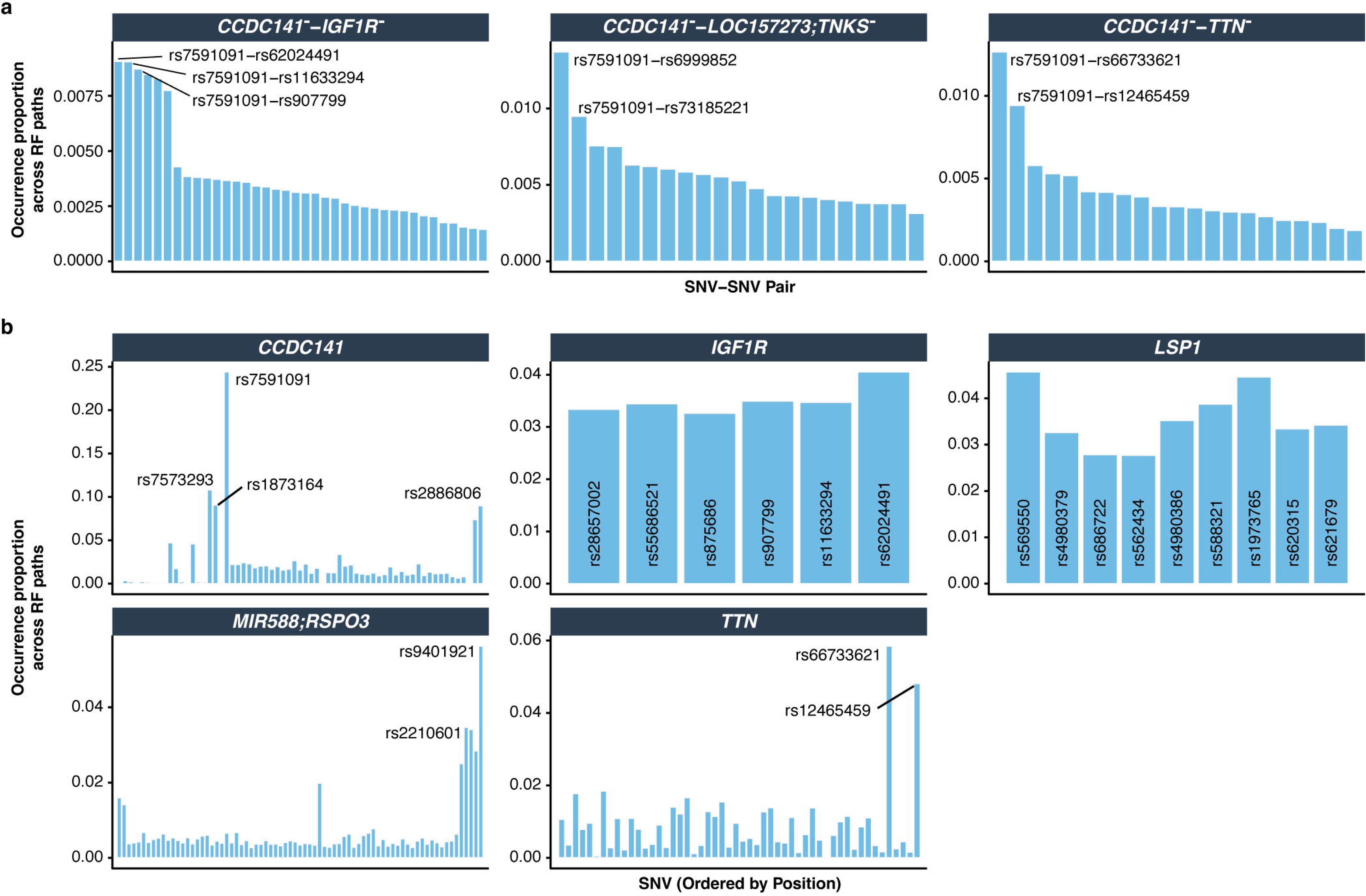
Top SNVs from lo-siRF-prioritized loci and interactions between loci. The most important SNVs and SNV-SNV pairs, as measured by their proportion of occurrence in the siRF fit, are annotated for the top lo-siRF-prioritized interactions between loci in **a** and top genetic loci in **b**. The y-axis shows the proportion of decision paths in siRF, for which the SNV or SNV-SNV pair occurs, averaged across all three binarization thresholds. In each of the interactions between genetic loci, rs7591091 in the *CCDC141* locus appears most frequently, suggesting a key role in cardiac hypertrophy.

**Fig. S5:**
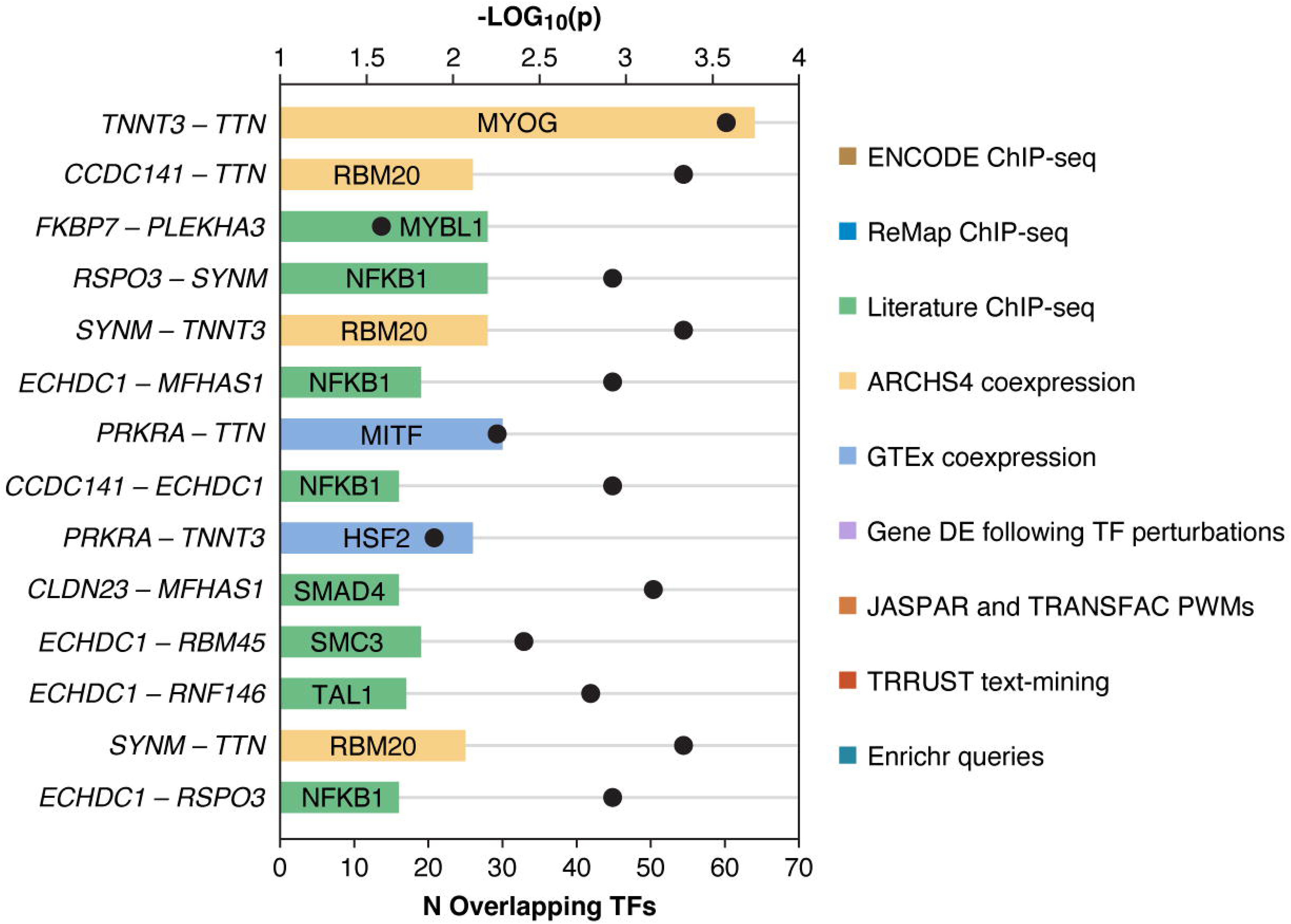
Genes mapped from epistatic loci share transcription factors and splicing regulators. Gene pairs exhibiting strong co-association with transcription factors (TFs) and RNA-binding regulators (*p* < 0.05, two-sided Fisher’s exact test, Fig. 4e) are displayed. Horizontal bars indicate the number of shared TFs or RNA-binding regulators (bottom axis), with the top-ranked TF or regulator, determined by the lowest two-sided enrichment *p*-value (dots, top axis), labeled on each bar. Bars are color-coded by the corresponding gene set library. Detailed information can be found in Supplementary Data 7.

**Fig. S6:**
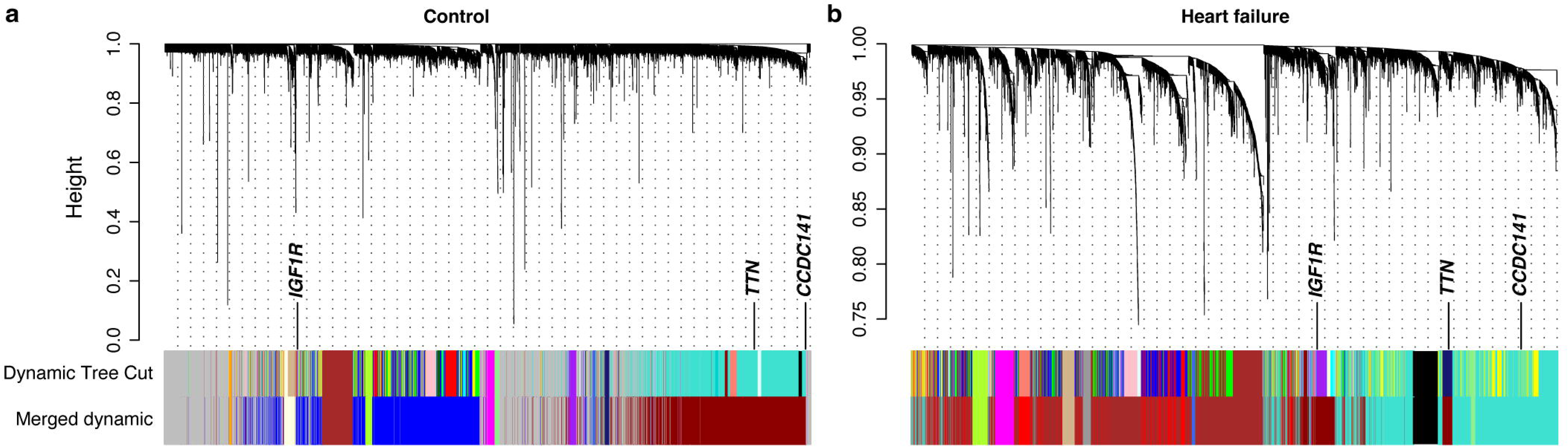
Dendrograms from WGCNA network analyses. Dendrograms from WGCNA control (**a**) and heart failure (**b**) networks show distinctive gene module structures and modular assignments for *CCDC141*, *IGF1R*, and *TTN*.

**Fig. S7:**
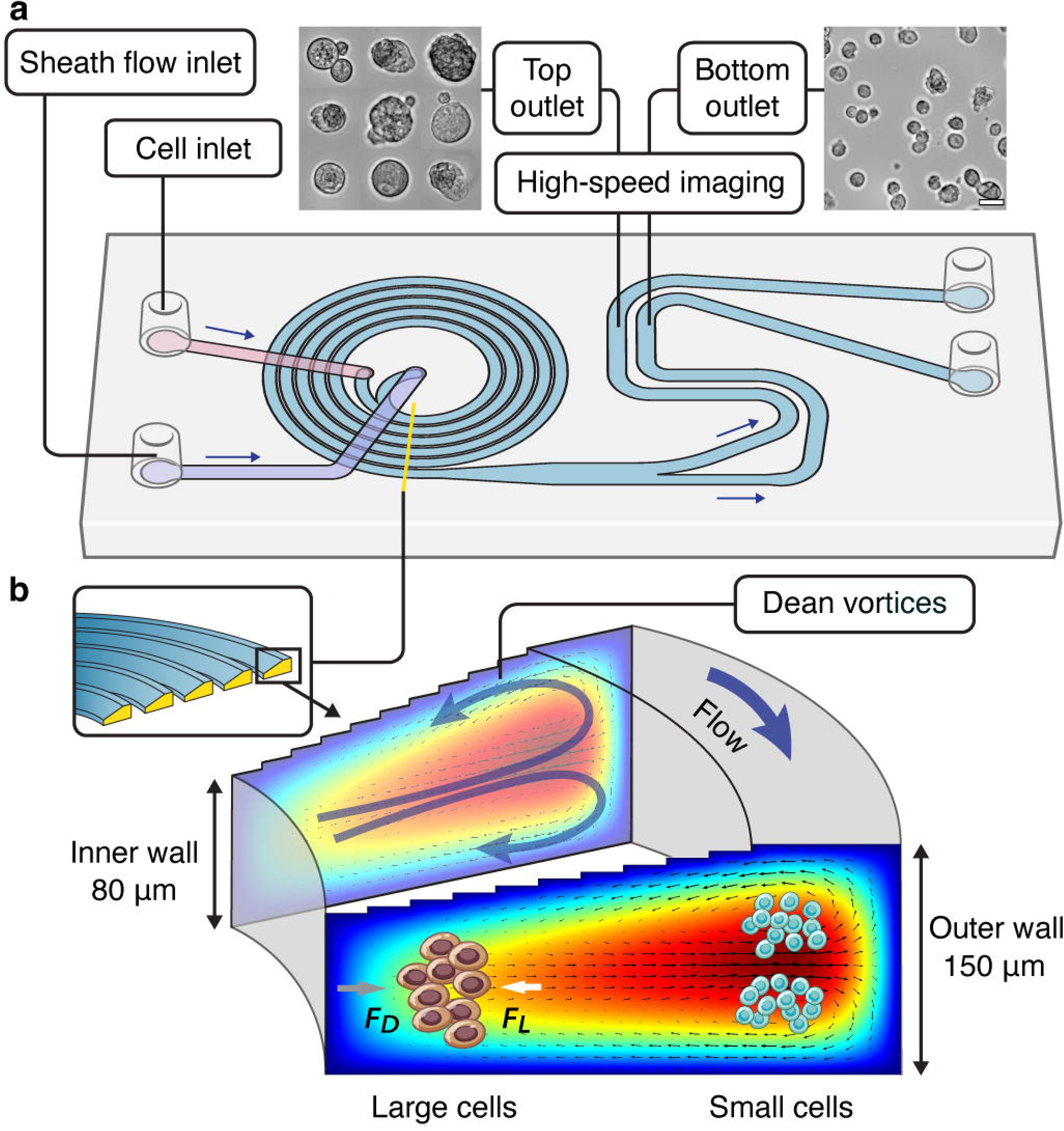
Spiral-shaped inertial microfluidic channel for cell focusing and imaging. a, Schematic of an inertial microfluidic cell focusing device. Cell suspensions and fresh medium were introduced into the microfluidic device through the cell and sheath flow inlets, respectively, using a syringe pump and flowed down the 5-loop spiral microchannel with the same flow rate (1.2 mL/min). Inserted microscopy images show that randomly dispersed cells were separated by size and bifurcated into the top (large cells) or bottom (small cells) outlets. Scale bar, 10 μm. Outlet channels are connected to straight observation channels where flowing cells were further focused in the channel height direction and imaged using a high-speed camera for morphological feature extraction (Fig. 6c). **b**, Schematic of the cell focusing principle. The spiral microchannel has a cross-section with a slanted ceiling, resulting in different depths at the inner and outer side of the microchannel. This geometry induces strong Dean vortices (counter rotating vortices in the plane perpendicular to the main flow direction) in the outer half of the microchannel cross-section. The interplay between drag forces (*F_D_*) induced by Dean vortices and lift forces (*F_L_*) due to shear gradient and the channel wall drives cell transverse migration towards equilibrium positions where the net force is zero. As a result, large cells in a heterogeneous population progressively migrate closer to the inner channel wall, while smaller cells move towards the outer channel wall. Details about microchannel dimensions can be found in Methods.

**Fig. S8:**
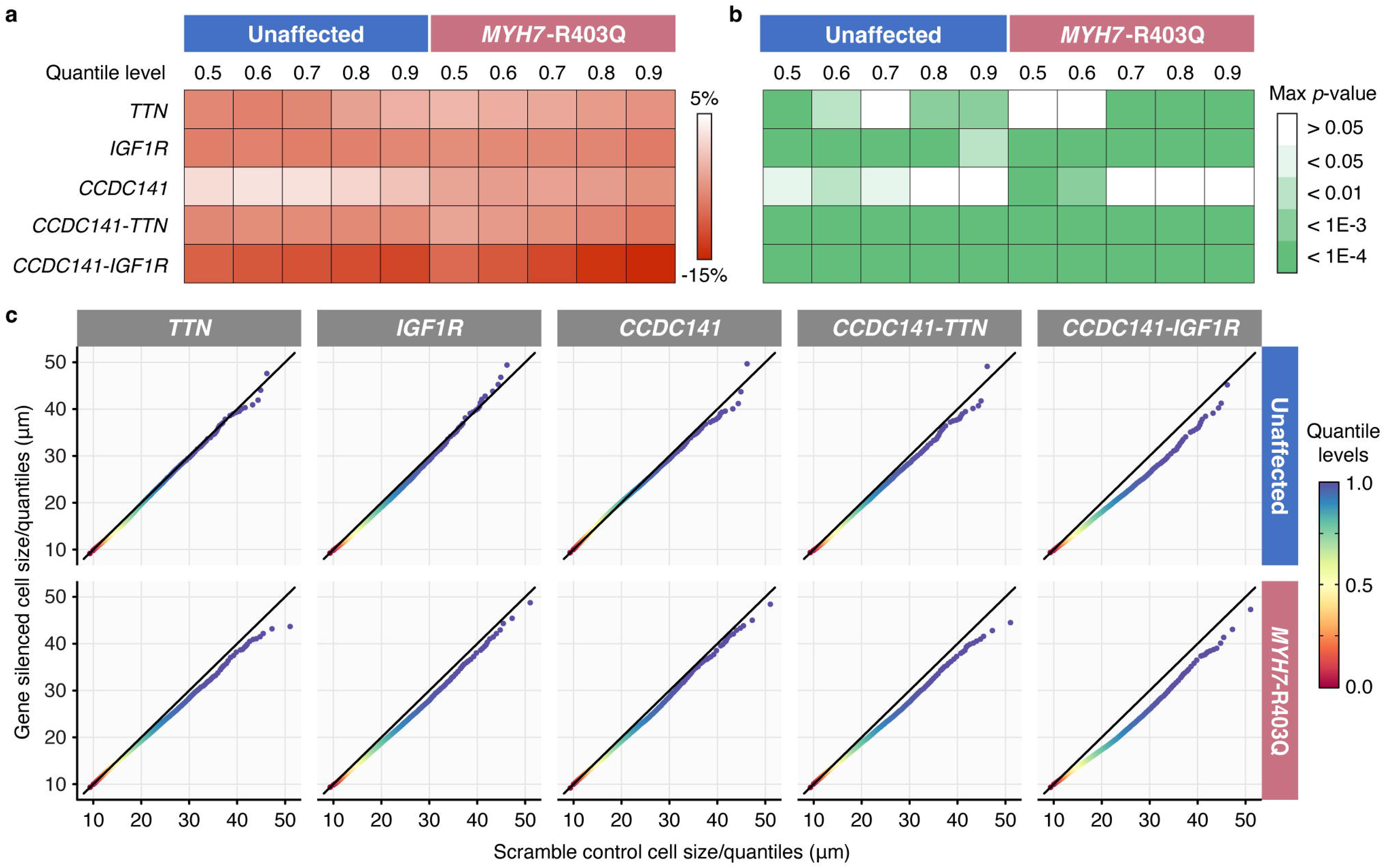
Epistatic genes non-uniformly reshape cardiomyocyte size distributions. **a**, Heatmap showing relative differences in cell sizes at various quantile levels between gene-silenced and scramble control conditions for unaffected and *MYH7*-R403Q variant cardiomyocytes. Larger quantiles correspond to larger cells in the cell size distribution. Dark red indicates strong size reduction at the specified quantile level in gene-silenced cells relative to scramble controls. The corresponding statistical differences (**b**) were evaluated by the maximum (two-sided) *p*-values across *n* = 3 (*n* = 4 for unaffected cells silencing *CCDC141*) independent cell batches using a bootstrap quantile test with 10,000 bootstrapped samples (exact *p* values per batch are available in Source Data). **c**, QQ-plots of averaged cell size quantiles across cell batches compare gene-silenced cells against scramble controls in unaffected (top row) and *MYH7*-R403Q variant (bottom row) cardiomyocytes, revealing a clear size-dependent effect of silencing *CCDC141*-*IGF1R* on correcting cardiomyocyte hypertrophy.

**Fig. S9:**
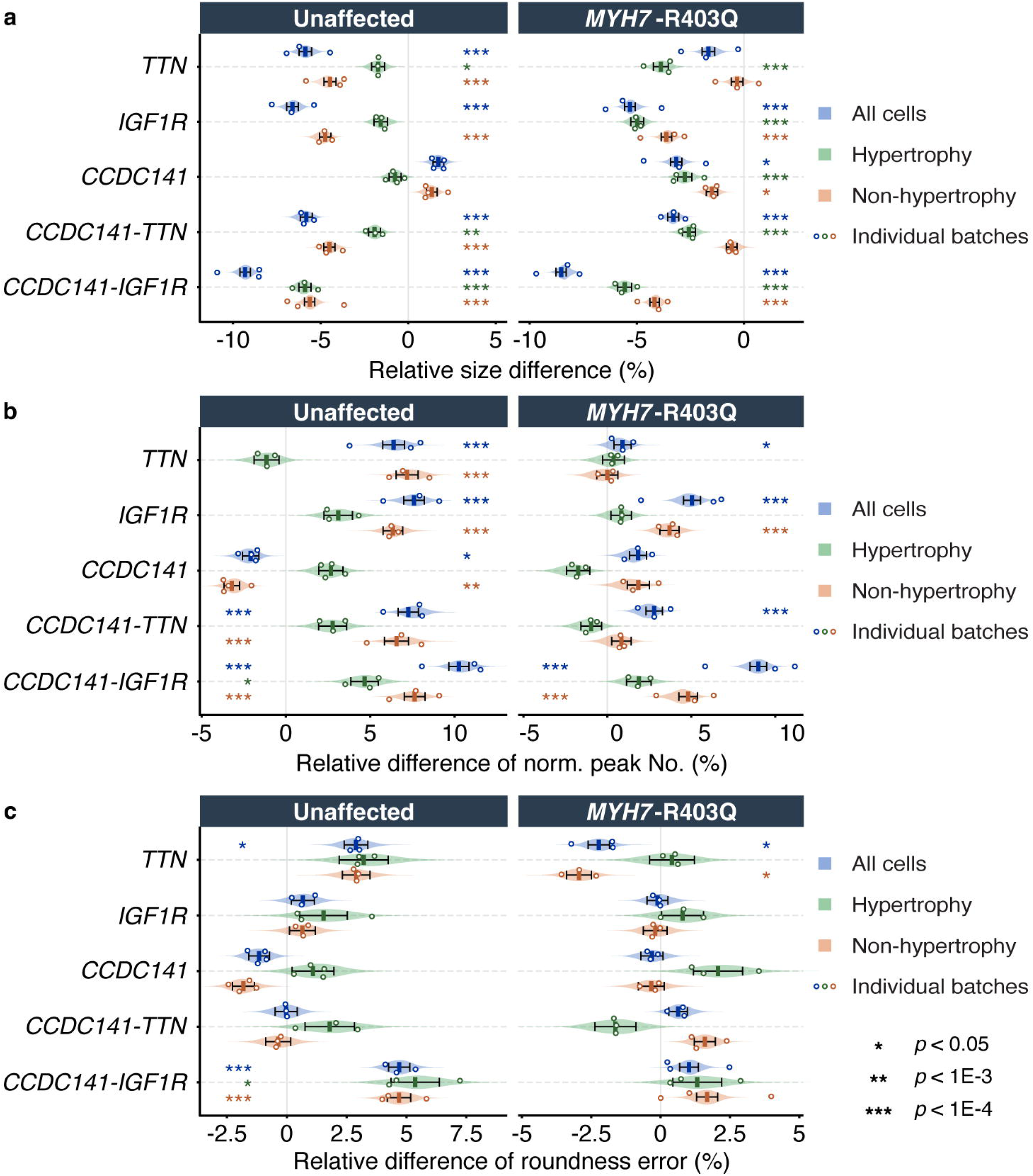
Effects of lo-siRF-prioritized genes and gene-gene interactions on hypertrophic and non-hypertrophic cell morphology. Relative differences in median cell size (**a**), normalized peak number (**b**), and roundness error (**c**) were analyzed separately for cells size-sorted into the top (hypertrophic cells, green) and bottom (non-hypertrophic cells, orange) microchannel outlets (Extended Data Fig. 7), as well as for all cells combined (blue). For both unaffected (left) and *MYH7*-R403Q variant (right) cardiomyocytes, the gene-silencing induced relative differences in each morphological feature (**a**-**c**) were quantified as (*m_S_* - *m_C_*)/*m_C_* x 100%, where *m_S_* and *m_C_* denote median values in gene-silencing and scrambled control conditions, respectively. Squares represent mean relative differences across *n* = 3 independent cell batches (*n* = 4 for unaffected cells silencing *CCDC141*), with overlaid points indicating medians of individual replicates. Violins display mean relative differences from 1,000 bootstrap samples, with error bars indicating standard deviations. Asterisks indicate significant differences compared to scramble controls, based on the maximum *p*-values of two-sided Mann-Whitney U test across all cell batches (\**p* < 0.05, \*\**p* < 1E-3, \*\*\**p* < 1E-4). Exact *p*-values and corresponding cell counts per condition per batch are provided in Source Data.

