## Supplementary Notes for "Deciphering epistatic genetic regulation of cardiac hypertrophy"

**Supplementary Note 1: PCS documentation for low-signal signed iterative random forest analysis**

An HTML (webpage style) document that can be downloaded and viewed in the browser or via the website https://yu-group.github.io/epistasis-cardiac-hypertrophy/. Guided by the PCS framework for veridical data science, we transparently document and justify the many human judgment calls and modeling decisions (e.g., choice of dimension reduction and binarization thresholds) that were made throughout the lo-siRF pipeline. Furthermore, we provide additional exploration and stability analyses to ensure that our findings are stable and robust across these choices. These additional analyses include an investigation of possible pleiotropic effects of the identified epistasis on both LV mass and blood pressure as well as comparisons between the lo-siRF-prioritizations and those from common existing methods. We also provide simulations, showing that lo-siRF produces calibrated *p*-values under a null response model and investigates the performance of lo-siRF under both a marginal effect and interaction effect simulation models.

**Supplementary Note 2: Implementation of alternative epistasis detection methods as a comparison to lo-siRF**

*2.1 Exhaustive regression-based pairwise interaction scan*[1,2]

For each pair of SNVs that passed the GWAS filter in lo-siRF step 1, we fit the follow regression:

$$y_i = \beta_0 + \beta_1 g_{i1} + \beta_2 g_{i2} + \beta_{12} g_{i1} g_{i2} + \gamma^T z_i + \epsilon_i, \qquad \text{(S1)}$$

where $y_i$ is the rank-based inverse normal-transformed LVMi for individual $i$, $g_{ij}$ is the genotype of SNV $j$ for individual $i$, $z_i$ is a vector of covariates for individual $i$ (i.e., sex, age, height, body weight, and the first five principal components of ancestry), and $\epsilon_i$ is the random error or noise term for individual $i$. Under this regression model, we tested the null hypothesis of $\beta_{12} = 0$ versus the alternative hypothesis of $\beta_{12} \neq 0$ via the traditional t-test. We also repeated this exhaustive interaction search using the binarized LVMi response for each of the three different binarization thresholds (15%, 20%, and 25%). For the binarized LVMi, we used a logistic regression in lieu of the linear regression and tested for a non-zero $\beta_{12}$ coefficient via the traditional Wald z-test. Results are discussed in Supplementary Note 1.

##### *2.2 MAPIT[3]*

MAPIT leverages a variance component model to first identify candidate variants with non-zero marginal epistatic effects, defined as the total pairwise interaction effect between the variant and all other variants[3]. By focusing on these marginal epistatic effects, MAPIT can advantageously search for epistatic variants without enduring the computational and statistical burdens associated with pinpointing their epistatic partners. We performed MAPIT using the mvMAPIT (v2.0.3) R package. For inputs, we used the 1405 GWAS-filtered SNVs with minor allele frequency > 0.05, adjusted for sex, age, height, body weight, and the first five principal components of ancestry, and used the rank-based inverse normal-transformed LVMi as the response. We used the default settings in the mvmapit function and chose the “normal” test to minimize the computational burden. Results are discussed in Supplementary Note 1.

##### *2.3 MAPIT[3] + Gene Set Enrichment Analysis[4,5]*

Because MAPIT was originally developed to identify epistatic variants rather than loci (as in lo-siRF), we conducted a locus-level analysis using MAPIT[3] in conjunction with gene set enrichment analysis (GSEA)[4,5]. In this analysis, we took the union of the top 10,000 GWAS hits from PLINK and BOLT-LMM and performed MAPIT on these SNVs (using the same settings as above) to obtain a ranked list of SNVs according to their MAPIT *p*-values. We then identified loci showing statistically significant enrichment at the top of this ranking using the pre-ranked gene list tool in GSEA (v4.3.3) with 1,000 permutations, the classic (unweighted) enrichment statistic, a maximum set size of 500, a minimum set size of 1, while keeping all other options at their default settings. Results are discussed in Supplementary Note 1.

##### *2.4 Alternative set-based genome-wide association tests*

To investigate the importance of the *IGF1R* locus using existing set-based association methods, we performed SKAT-O[6] using the subset of 1405 GWAS-filtered SNVs with minor allele frequency > 0.05 as input and the rank-based inverse normal-transformed LVMi as the response. We also adjusted for sex, age, height, body weight, and the first five principal components of ancestry in the SKAT-O null model. This analysis was carried out using the SKAT R package. In addition to SKAT-O, we also ran the gene-based test as computed by MAGMA[7] using the LVMi PLINK GWAS results as input. This MAGMA analysis was carried out using FUMA with the default settings. Results of these two analyses are detailed in Supplementary Note 1.

**Supplementary Note 3: Dimensions and fabrication of the microfluidic device**

*3.1 Microchannel geometry and dimensions*

The microfluidic device (Extended Data Fig. 7) contains 5 loops of spiral microchannel with a radius increasing from 3.3 mm to 7.05 mm. The microchannel has a cross-section with a slanted ceiling, resulting in 80 μm and 150 μm depths at the inner and outer side of the channel, respectively. The channel width is fixed to 600 μm. The 495 μm wide slanted region of the channel ceiling is composed of ten 7 μm deep stairs. The device has two inlets at the spiral center to introduce cell suspensions and sheath flow of fresh medium. At the outlet region, the channel is expanded in width and split into two outlet channels with a width of 845 μm for the top outlet and 690 μm for the bottom outlet. Depths of the two outlet channels are designed to create equal hydraulic resistance. The top and bottom outlet channels are connected to 80 μm and 50 μm deep straight observation channels for high-throughput cell imaging.

*3.2 Microdevice fabrication*

The spiral microchannel was fabricated by CNC micromachining a piece of laser-cut poly (methyl methacrylate) (PMMA) sheet, which was bonded with a PMMA chip machined only with the inlet channels and another blank PMMA chip using a solvent-assisted thermal binding process to form the enclosed channel[8]. Before bonding, PMMA chips were cleaned with acetone, methanol, isopropanol and deionized water in sequence. Droplets of a solvent mixture (47.5% DMSO, 47.5% water, 5% methanol) were evenly spread over the cleaned chips. The PMMA chips were assembled appropriately and clamped using a customized aluminum fixture, and then heated in a ThermoScientific Lindberg Blue M oven at 96 ℃ for 2 hrs. After bonding, fluid reservoirs (McMaster) were then attached to the chips using a two-part epoxy (McMaster). Microchannels were flushed with 70% ethanol followed by DI water for sterilization.

**Supplementary Note 4: *CCDC141-IGF1R* and *CCDC141-TTN* interactions non-additively reshape cardiomyocyte size distribution**

Given the diverse expressivity of cellular hypertrophy among diseased cells (indicated by the heavily positively-skewed size distribution in Fig. 7a), we further analyzed size changes of large and small cells focused into different microchannel outlets (Extended Data Fig. 7) separately. Our results show that silencing *TTN* ($p$ = 7E-14, two-sided Mann-Whitney U test) or *CCDC141-TTN* ($p$ = 1E-5) predominantly targets cells that exhibit cellular hypertrophy (green in Extended Data Fig. 9a, right) but has a trivial impact on small cells (orange in Extended Data Fig. 9a, right).

To explore this diverse expressivity further, we evaluated how gene silencing reshapes the cell size distribution (Extended Data Fig. 8a and $p$-values in 8b). QQ-plots comparing the quantiles of gene-silenced cells against their scrambled controls suggest that digenic gene silencing leads to a stronger effect on larger cells, as indicated by the greater deviation of points from the solid black line in Extended Data Fig. 8c). For example, the upper quantiles of diseased cells silencing *CCDC141-IGF1R* are much lower (the 0.9 quantile decreased by 14.4%, $p$ < 1E-4, two-sided bootstrap quantile test with 10,000 bootstrapped samples) than the corresponding scrambled quantiles, suggesting that silencing this gene pair favors rescuing hypertrophic cells over small cells. However, this size-dependent effect is mitigated in cells with monogenic silencing of *CCDC141* or *IGF1R*. This discrepancy in how these two genes affect cell size reinforces the hypothesized non-additive interaction between *CCDC141* and *IGF1R* (e.g., $p$ < 1E-4 for non-additivity, 0.9 quantile). Importantly, stable non-additive interaction effects for hypertrophic cells (quantile levels higher than 0.6) are also observed for the *CCDC141-TTN* pair in both cell lines (Supplementary Data 8). Overall, these experimental results suggest both an epistatic interaction between *CCDC141* and the other two genes (*IGF1R* and *TTN*) as well as independent effects on modifying cardiomyocyte hypertrophy.

**Supplementary Note 5: Simulations examining the effect of varying gene-silencing efficiencies on epistasis testing**

An HTML (webpage style) document that can be downloaded and viewed in the browser or via the website https://yu-group.github.io/epistasis-cardiac-hypertrophy/simulations_efficiency. We describe the simulation study used to examine how different gene-silencing efficiencies impact the significance testing for epistasis. These simulations cover various silencing efficiencies and different signal-to-noise regimes. Results from this simulation study are also provided and discussed in detail.

### Supplementary Table Legends

**Supplementary Table 1: Characteristics of 29,661 analyzed participants in the UK Biobank.**

Summary statistics of the 29,661 unrelated White British individuals analyzed in this study. Means and standard deviations (in parentheses) are reported for continuous measurements (LVMi, LVM, age, height, and weight) alongside the number of individuals (N) and the percentage of individuals with various cardiac hypertrophy-related diseases or on blood pressure medication. We define hypertensive individuals as anyone with self-reported hypertension, high blood pressure as diagnosed by a doctor, or any ICD10 billing code diagnosis in I10-I16; aortic stenosis as self-reported aortic stenosis or an ICD10 billing code diagnosis of I35; heart failure as self-reported heart failure or an ICD10 billing code diagnosis of I50; and type II diabetes as self-reported type II diabetes or an ICD10 billing code diagnosis of E11.

**Supplementary Table 2: Thresholds defining low and high LVMi groups used in siRF fit**

For each of the three binarization thresholds used in lo-siRF (corresponding to the bottom/top $15^{th}$, $20^{th}$, and $25^{th}$ quantiles), we provide the sex-specific LVMi cutoffs for the low and high LVMi groups. All thresholds were measured in g/$m^2$.

**Supplementary Table 3: Prediction accuracies of methods for predicting the continuous LVMi phenotype without binarization**

Commonly used machine learning methods yield validation $R^2$ values that are slightly less than 0 for predicting the continuous LVMi phenotype without binarization. These prediction models are no better than constantly predicting the mean LVMi (which would yield an $R^2$ value of 0) and thus do not pass the prediction check under the PCS framework. This motivates the need for an alternative approach, such as binarization. Abbreviations: RMSE, root mean squared error; MAE, mean absolute error.

**Supplementary Table 4: Prediction accuracies of methods across different LVMi binarization thresholds.**

Maximum prediction accuracies highlighted in bold. The siRF model performs better or on par with other commonly used machine learning methods when predicting the binarized LVMi phenotype. This result holds across all three binarization thresholds and three different classification metrics, i.e., classification accuracy, area under the receiver operator curve (AUROC), and area under the precision-recall curve (AUPRC). In accordance with the prediction check component of the PCS framework, siRF is an appropriate fit for the given data.

**Supplementary Table 5: Top signed loci and interactions between loci, prioritized by lo-siRF across LVMi binarization thresholds.**

A list of the top signed loci and interactions between loci, prioritized by lo-siRF, that were stably important across all three LVMi binarization thresholds (Supplementary Table 2). *P*-values for each binarization threshold were computed using a two-sided permutation test in lo-siRF. The loci and interactions between loci are ranked by the mean nominal lo-siRF *p*-value (i.e., the average *p*-value across the three binarization thresholds). Note that since the permutation tests were run using 10,000 permutations, the obtained *p*-values cannot be made more precise than 1E-4.

**Supplementary Table 6: List of prediction methods and hyperparameters used in prediction check.**

A list of each model alongside their implementations (software, package, and function) and hyperparameters used during the prediction check in lo-siRF. All hyperparameters were tuned using 5-fold cross-validation.

**Supplementary Table 7: Summary of siRF evaluation metrics for top interactions between loci.** Though prediction accuracy is weak (indicated by precision scores close to 0.5), the lo-siRF-prioritized interactions are stable across binarization thresholds and across bootstrap replicates (indicated by all types of stability scores being close or equal to 1). Here, prevalence measures the proportion of high LVMi individuals for which the interaction appears. Precision measures the probability of having high LVMi given that the interaction is active. The class difference in prevalence is the prevalence of the interaction in high LVMi individuals minus the prevalence in low LVMi individuals. Feature selection dependence evaluates whether the interaction is collectively or individually associated with the responses. The stability of each of these metrics evaluates how stable the respective scores are across 50 bootstrap replicates. The overall stability score (last column) is the proportion of times that the interaction is identified by siRF across 50 bootstrapped replicates. Higher scores for each listed metric indicate greater importance.

### Supplementary Data Legends

**Supplementary Data 1: GWAS results for left ventricular mass.**

A summary of the left ventricular mass (LVM) and left ventricular mass index (LVMi) genome-wide association study results using PLINK and BOLT-LMM. This includes a list of the LVM/LVMi risk loci, lead SNVs, independent significant SNVs, and positionally mapped genes from FUMA GWAS.

**Supplementary Data 2. List of GWAS-filtered SNVs used in the lo-siRF pipeline.**

A list of the 1405 SNVs that passed the genome-wide association study dimension reduction step in the lo-siRF pipeline. This table includes the rsID, chromosome, hg19 position, allelic information, corresponding genetic loci in lo-siRF, and inferred (exonic) function.

**Supplementary Data 3. The lo-siRF-prioritized SNVs and SNV-SNV epistatic interactions.**

A summary of the 283 lo-siRF-prioritized SNVs that show stable association with left ventricular mass index (LVMi), which are aggregated into six left ventricular hypertrophy risk loci (Table 1) according to ANNOVAR gene annotation in lo-siRF. The spreadsheet also includes separate tabs showing the prioritized SNV-SNV pairs for each of the three locus-to-locus interactions (*CCDC141-IGF1R*, *CCDC141-TTN*, and *CCDC141-LOC157273;TNKS*) stably identified by all the three LVMi binarization thresholds in lo-siRF.

**Supplementary Data 4. Functional annotations for lo-siRF-prioritized SNVs and their LD-linked SNVs extracted by FUMA, along with the functionally mapped genes.**

The spreadsheet includes a list of the 283 lo-siRF-prioritized SNVs and the FUMA-extracted candidate SNVs in LD with any of the 283 lo-siRF-prioritized SNVs, as well as their corresponding functional annotations from ANNOVAR, ChromHMM core-15 chromatin state, CADD score, RegulomeD score, eQTL and sQTL information from GTEx, and GWAS statistics. The spreadsheet also includes a list of protein-coding genes functionally linked from the 283 lo-siRF-prioritized SNVs and their LD-linked SNVs by positional, eQTL, and chromatin interaction mapping in FUMA.

**Supplementary Data 5. ANNOVAR Enrichment analysis results.**

ANNOVAR enrichment of functional consequences of all SNVs (both the lo-siRF-prioritized SNVs and their LD-linked SNVs extracted by FUMA, details in Supplementary Data 4) associated to each of the six lo-siRF-prioritized risk loci for left ventricular hypertrophy.

**Supplementary Data 6. Gene ontology and pathway co-association network.**

Co-association network between genes that are functionally linked to the lo-siRF-prioritized epistatic and hypostatic loci (Supplementary Data 4) and their shared GO or pathway terms. GOs and pathways are enriched from multiple annotated gene set libraries from Enrichr (Fig. 4c).

**Supplementary Data 7. Transcription factor co-association network.**

The spreadsheet includes two lists of transcription factors and RNA-binding regulators enriched from lo-siRF-prioritized and lo-siRF-deprioritized genes (Fig. 4d), respectively. It also includes a summary of co-associations between lo-siRF-prioritized genes and their shared regulatory factors (Fig. 4e). Transcription factors and RNA-binding regulators in the co-association network are enriched from nine distinct annotated gene set libraries from Enrichr and ChEA3.

**Supplementary Data 8. Single cardiomyocyte morphology assessment and statistical analysis.**

A summary of the epistatic relationships in cardiomyocyte morphology confirmed by gene perturbation experiment (Fig. 7). The spreadsheet includes an evaluation of epistatic effects on cell size (at various quantile levels), cell textural irregularity, and cell boundary irregularity, supported by multiple statistical analysis and assessment of non-additivity for the investigated gene-gene interactions.
