## Supplementary webpage 1 for "Deciphering epistatic genetic regulation of cardiac hypertrophy": supplementary_note1.html

PCS documentation for low-signal signed iterative random forests (lo-siRF)


### PCS documentation for low-signal signed iterative random forests (lo-siRF)

###### September 27, 2024

- 1 Overview
- 2 Choice of Phenotypic Data
- 3 Dimension Reduction Step
- 4 Binarization Step
- 5 Prediction Step
- 6 Prioritization Step
- 7 Non-hypertensive Analysis
- 8 Comparison to Existing Methods
- 9 Lo-siRF Permutation Test Simulations
- 10 Final Remarks
- 11 Bibliography

### 1 Overview

In Q. Wang et al. (2023), we developed an end-to-end pipeline to demonstrate the role of epistasis in the genetic control of cardiac hypertrophy.
This pipeline consisted of four major phases:
(1) the derivation of estimates of left ventricular mass via deep learning,
(2) the computational prioritization of epistatic drivers based on data from the UK Biobank (Bycroft et al. 2018),
(3) functional interpretation of the hypothesized epistatic genetic loci, and
(4) experimental confirmation of epistasis in cardiac transcriptome and cardiac cellular morphology.

Here, we expand upon the computational prioritization of epistatic drivers phase.
In this phase, we leveraged large-scale genetic and cardiac MRI data from the UK Biobank and developed the low-signal signed iterative random forest (lo-siRF) to prioritize genetic loci and interactions between loci for downstream investigations.
As lo-siRF is heavily rooted in the Predictability, Computability, and Stability (PCS) framework for veridical (trustworthy) data science (Yu and Kumbier 2020), we conducted extensive stability analyses to help minimize the impact of human judgment calls and modeling decisions on our scientific conclusions.
In this supplementary PCS documentation, we will:

1. Transparently document and justify the many human judgment calls and modeling decisions that were inevitably made throughout our statistical analysis.
2. Provide additional exploration and insight into the main results presented in Q. Wang et al. (2023) alongside the stability analyses.

In what follows, we organize this documentation by step in the lo-siRF pipeline (see Q. Wang et al. (2023)), beginning with the choice of phenotypic data, followed by the dimension reduction step, binarization step, prediction step, and concluding with the prioritization step.
We then investigate the possibility of pleiotropy between LV mass and blood pressure, conducting a stability analysis where we examine the impact of excluding hypertensive individuals from the lo-siRF analysis.
Lastly, we compare the lo-siRF prioritized loci and interactions between loci with those identified using existing methods and conclude with final remarks.

⚠
**Origins and intended usage of lo-siRF:**
As discussed in Q. Wang et al. (2023), lo-siRF was originally developed as a first-stage recommendation (or hypothesis generation) tool to prioritize genetic loci and interactions between loci for downstream analyses and experimental validation.
Importantly, the output of lo-siRF is a prioritization or ranking order of loci and interactions between loci, not a measure of statistical significance.
We emphasize that recommendations or prioritizations from lo-siRF should not be interpreted as causal findings without further investigation.
Given the numerous challenges inherent to epistasis discovery (e.g., high heterogeneity, small effect sizes, and unmeasured confounding factors, discussed further in Q. Wang et al. (2023)), assessing causality solely from data is incredibly complex and difficult to do *reliably*.
Lo-siRF was thus designed and intended to be used as a single step within a larger scientific discovery pipeline.
In Q. Wang et al. (2023), we relied on extensive follow-up investigations to validate the lo-siRF-prioritized genetic loci and interactions between loci, and we encourage practitioners to do the same.

### 2 Choice of Phenotypic Data

In the first step of lo-siRF, we began with a careful choice of phenotypic data.
This first, but often overlooked, choice is critical to the reliability and interpretation of results.
If the data quality is poor, conclusions may be unreliable.
If the data is far-removed from the domain problem of interest, conclusions may not be relevant.
Given the importance of this choice of phenotypic data, we conducted extensive data explorations for different phenotypes in addition obtaining clinician input regarding its clinical relevance.

**Initial choice of phenotype:**
Our overarching goal was to study the role of epistasis in cardiac hypertrophy.
One important disease that is closely related to cardiac hypertrophy is hypertrophic cardiomyopathy (HCM).
HCM is a common heart disease, impacting around 1 in 500 people, where the heart muscle becomes enlarged or thickened.
Given the high prevalence and possible life-threatening consequences of this disease, we first set out to explore this HCM phenotype, defined as a 0-1 indicator variable of whether or not the individual was diagnosed with HCM (i.e., any ICD10 code of I42.1 or I42.2 in the UK Biobank).
However, when fitting various machine learning prediction models, including L1 or L2-regularized logistic regression, random forest (RF), iterative RF (iRF), and support vector machines, using single-nucleotide variant (SNV) data from the UK Biobank (details in Q. Wang et al. (2023)) to predict HCM diagnosis, the balanced classification accuracy on the held-out validation data did not consistently exceed 50% when trained on different bootstrapped training samples.
In other words, the fitted prediction models were no better than random guessing.
This made us wary of any interpretations extracted from these prediction models since it is unclear whether these models are capturing any relevant phenotypic signal or simply noise.
Given this incredibly weak prediction signal, we deemed that these models did not pass the prediction check (the “P” in PCS) (Yu and Kumbier 2020).
For this reason, we chose not to proceed further with the HCM phenotype and did not conduct any analysis with respect to interpreting the aforementioned models for HCM.

**Pivoting to a cardiac MRI-derived phenotype:**
We instead pivoted to an alternative phenotype for cardiac hypertrophy based upon deep-learning-enabled left ventricular (LV) structural analysis using cardiac MRIs.
This choice was heavily motivated by our struggles with HCM.
Specifically, when looking back at the HCM diagnosis data, we noticed that the number of HCM cases in the UK Biobank data (only 236 HCM cases out of 337,535 unrelated White British individuals) was much lower than the 1 in 500 ratio that is expected.
Although the UK Biobank population is known to be healthier than the general population (Fry et al. 2017), the observed discrepancy is large, leading us to speculate about potential under-diagnosis issues with the HCM phenotype.
Consequently, rather than relying on a clinician’s diagnosis which may suffer from issues such as under-diagnosis, we chose to build upon recent advances in deep-learning-enabled phenotyping (Bai et al. 2018).
Doing so enables us to extract a measure of left ventricular hypertrophy, specifically, the left ventricular mass (LVM), from anyone with a cardiac MRI.

**Arriving at LVMi:**
However, LVM is known to be heavily influenced by body weight, height, and gender (Engel, Schwartz, and Homma 2016; Mizukoshi et al. 2016).
Thus, if we were to proceed solely with LVM in our analysis, we risk identifying genetic loci and interactions between loci that are primarily associated with body weight, height, and gender, rather than left ventricular hypertrophy.
Indeed, if we visualize the relationship between LVM, height, and weight within each gender group (Figure 2.1), we see strong positive correlations.

This motivates the need to adjust for height and weight in order to help decouple the effects of height/weight and LVM.
As done in Khurshid et al. (2023), one common way to address this is by analyzing LVM indexed by body surface area (LVMi), defined as:

\[\begin{align\*}
\text{LVMi} = \text{LVM } / \text{ Body surface area},
\end{align\*}\]
where body surface area is calculated based on height and body weight via the Du Bois formula (Du Bois and Du Bois 1916).

After normalizing by body surface area (and hence, height and weight), the associations between LVMi and height/weight are much weaker within each gender group, exhibiting correlations closer to 0 (Figure 2.1).
Thus, by focusing our statistical analysis on LVMi, we lessen the possibility that the identified genetic loci and interactions between loci are only selected because of their association with body height and/or weight.
Still, there are differences in LVMi between males and females, with males tending to have higher LVMi measurements.
As seen later, we will account for this in the modeling stage by using gender-specific binarization thresholds to binarize the LVMi responses into the high and low LVMi groups.

**Study Cohort:**
Before proceeding, we note that another important human judgment call within our pipeline is the choice of study cohort.
Specifically, we restricted our analysis to the unrelated White British individual population with both SNV and cardiac MRI data in the UK Biobank, resulting in a cohort of 29,661 individuals.
We chose to focus this analysis on the unrelated White British population, anticipating the very low-signal problem under study (e.g., due to the generally small effect sizes of common genetic variants and the high phenotypic diversity of cardiac hypertrophy).
By studying a more homogeneous cohort such as the unrelated White British population, we hope to first establish a proof-of-concept foothold on the role of epistasis in cardiac hypertrophy, and we view the study and generalization to the broader population as a very crucial next step.

Figure 2.1: Pair plot, illustrating the relationships between LVM, LVMi, height, and weight within each gender group (males in green and females in orange). Density distributions are shown along the diagonal.

### 3 Dimension Reduction Step

#### Overview

Having carefully chosen the phenotypic data (i.e., the cardiac MRI-derived LVMi) under study, the next step in lo-siRF is to do dimension reduction to reduce the number of SNVs under consideration using a genome-wide association study (GWAS).
This dimension reduction step is necessary to mitigate the computational and statistical challenges of searching across millions of possible SNVs (which in the case of searching for epistasis results in an even larger combinatorial explosion of possibilities).

**Why GWAS?**
We chose to do dimension reduction by running a GWAS as it
(1) has been shown to be an incredibly useful tool for better understanding the genetic basis of numerous diseases in this field (Visscher et al. 2017; Pirruccello et al. 2020; Meyer et al. 2020; Harper et al. 2021; Khurshid et al. 2023),
(2) is one of the few methods that can handle an input of millions of SNVs in a computationally-tractable manner, and
(3) is a common, widely-accepted preprocessing step in other related statistical genomics tasks such as fine-mapping (Schaid, Chen, and Larson 2018).

**Limitations:**
We acknowledge, however, that using a GWAS to reduce the number of SNVs under consideration has limitations.
Because a GWAS prioritizes SNVs that have strong marginal associations with the phenotype under study, we may be removing SNVs that are associated with LVMi through an epistatic interaction but not through a marginal association.
Alleviating this limitation is certainly interesting and an important direction to pursue in future work.
Regardless, in this current work, we use lo-siRF primarily as a way to generate recommendations for experimental validation of epistasis.
We thus are most interested in generating *reliable* recommendations for a limited number of experiments, rather than attempting to find all possible interactions that drive the disease.
Given our particular use-case for lo-siRF, we accept the limitations of using a GWAS for dimension reduction and again view this work as a reliable *first* step (and certainly not the last) for better understanding the role of epistasis in cardiac hypertrophy.

**How was the dimension reduction performed?**
We performed a GWAS on the training data for the rank-based inverse normal-transformed LVMi using two algorithms, PLINK (Purcell et al. 2007) and BOLT-LMM (Loh et al. 2015).
For each of the two GWAS runs, we ranked the SNVs by significance (i.e., the GWAS p-value).
We then took the union of the top 1000 SNVs (without clumping) from each of the two GWAS runs and proceeded with this resulting set of 1405 SNVs for the remainder of the lo-siRF pipeline.
We refer to Q. Wang et al. (2023) for the details on the PLINK and BOLT-LMM implementations and use this document instead to justify several of our human judgment calls in this dimension reduction step.

- **Why PLINK and BOLT-LMM?** Since BOLT-LMM and PLINK rely on different statistical models and it is unclear a priori which model is better suited for our problem, we employed both software packages to mitigate the dependence of downstream conclusions on this arbitrary choice of model.
- **Why the rank-based inverse normal-transformed LVMi?** Because the distribution of LVMi measurements is right skewed (see Figure 2.1), and the p-value computation in PLINK and BOLT-LMM rely on normality assumptions, the rank-based inverse normal-transform was taken as in Khurshid et al. (2023).
- **Why was the union taken?** As mentioned previously, relying on a GWAS to do dimension reduction can be limiting since it is primarily assessing marginal associations between the SNVs and the phenotype. There are also other restrictive modeling assumptions (e.g., linearity) that may add to these limitations. Due to this concern that we may be imposing too many unnecessary restrictions in order for SNVs to pass the GWAS filter, we chose to take the union of the top SNVs from BOLT-LMM and from PLINK, rather than taking the intersection which would have made it more difficult or restrictive for SNVs to pass the GWAS filter.
- **How was the threshold chosen?** We chose the threshold of 1000 SNVs per GWAS method (without clumping) as it (1) struck a balance between the amount of information loss and the computational cost of our downstream modeling and (2) yielded the highest validation prediction accuracy compared to choosing other possible thresholds (500 and 2000) with and without clumping in the prediction modeling stage. We note that this choice of the top 1000 SNVs includes all SNVs that passed the genome-wide significance level (p-value = 5E-8) as well as additional SNVs that did not pass the genome-wide significance level. In particular, taking the top 1000 SNVs corresponds to using the p-value threshold of 1.7E-5 and 6.7E-6 for PLINK and BOLT-LMM, respectively.

**Remark:** We note that alternative dimension reduction strategies may be used in replacement of our proposed procedure. For example, one may consider using a different GWAS software (e.g., REGENIE (Mbatchou et al. 2021)), a different phenotype transformation, or perhaps a more sophisticated approach tailored for interactions and epistasis detection (e.g., MAPIT (Crawford et al. 2017)).

**Organization:** Under the `GWAS Results Summary` tab, we provide an investigation into the BOLT-LMM and PLINK GWAS results for LVM and LVMi. Under the `GWAS-filtered SNVs`, we explore the selected 1405 SNVs that passed the GWAS filter for the LVMi phenotype.

#### GWAS Results Summary

Below, we provide an overview of the genome-wide association study (GWAS) results for both the LVM and LVMi phenotypes. While LVMi is the primary phenotype of interest, we provide the LVM GWAS results for completeness. We further investigate the similarities in findings between the LVM and LVMi GWAS analyses via a stability analysis.

- Under the `BOLT-LMM` and `PLINK` tabs:
  - We provide a brief look into the GWAS results, including the Manhattan plots for each phenotype (LVMi and LVM).
  - We also provide a table of the genomic risk loci, lead SNVs, and independent significant SNVs from each GWAS, determined using FUMA (Watanabe et al. 2017) with the default settings (i.e., maximum p-value of lead SNVs = 5E-8, maximum p-value cutoff = 0.05, \(r^2\) threshold to define independent significant SNVs = 0.6, 2nd \(r^2\) threshold to define lead SNVs = 0.1, minimum minor allele frequency = 0, maximum distance between LD blocks to merge into a locus = 250kb, using the UKB release2b 10k White British reference panel population).
  - Furthermore, we used FUMA to map SNVs to genes based on position, again with the default settings (maximum distance = 10kb), and provide this table of mapped genes below.
- Lastly, under the `Stability Analysis` tab, we investigated the similarities and differences between the BOLT-LMM and PLINK results as well as between the LVM and LVMi results.

##### BOLT-LMM

###### LVMi

Figure 3.1: GWAS Manhattan plot for the LVMi phenotype using BOLT-LMM. Red dashed line indicates the genome-wide significance level (p = 5E-8).

###### Genomic Risk Loci

Figure 3.2: **List of genomic risk loci from FUMA for the LVMi BOLT-LMM GWAS.** uniqID = Unique ID of SNVs consists of chr:position:allele1:allele2 where alleles are alphabetically ordered. rsID = rsID of the top lead SNV for the given genomic locus. chr = Chromosome of top lead SNV. pos = Position of top lead SNV on hg19. p = P-value of top lead SNV. start = Start position of the locus. end = End position of the locus. nGWASSNVs = Number of the GWAS-tagged candidate SNVs within the genomic locus. nIndSigSNVs = Number of the independent significant SNVs in the genomic locus. IndSigSNVs = rsID of independent significant SNVs in the genomic locus. nLeadSNVs = The number of lead SNVs in the genomic locus. LeadSNVs = rsID of lead SNVs in the genomic locus.

###### Lead SNVs

Figure 3.3: **List of lead SNVs from FUMA for the LVMi BOLT-LMM GWAS.** GenomicLocus = Index of assigned genomic locus (note: multiple independent lead SNVs can be assigned to the same genomic locus). uniqID = Unique ID of SNVs consists of chr:position:allele1:allele2 where alleles are alphabetically ordered. rsID = rsID of the SNV. chr = Chromosome. pos = Position on hg19. p = P-value from GWAS. nIndSigSNVs = Number of independent significant SNVs which are in LD of the lead SNV at \(r^2\) = 0.1. IndSigSNVs = rsID of independent significant SNVs which are in LD of the lead SNV at \(r^2\) = 0.1.

###### Independent Significant SNVs

Figure 3.4: **List of independent significant SNVs from FUMA for the LVMi BOLT-LMM GWAS.** GenomicLocus = Index of assigned genomic locus (note: multiple independent lead SNVs can be assigned to the same genomic locus). uniqID = Unique ID of SNVs consists of chr:position:allele1:allele2 where alleles are alphabetically ordered. rsID = rsID of the SNV. chr = Chromosome. pos = Position on hg19. p = P-value from GWAS. nGWASSNVs = Number of GWAS-tagged SNVs which are in LD of the independent significant SNV given \(r^2\) = 0.6.

###### Mapped Genes

Figure 3.5: **List of mapped genes (via positional mapping) from FUMA for the LVMi BOLT-LMM GWAS.** ensg = ENSG ID. symbol = Gene Symbol. chr = Chromosome. start = Starting position of the gene. end = Ending position of the gene. strand = Strand of the gene. type = Gene biotype from Ensembl. entrezID = entrez ID (if available). HUGO = HUGO (HGNC) gene symbol. pLI = pLI score (the probability of being loss-of-function intolerant) from ExAC database; the higher the score is, the more intolerant to loss-of-function mutations the gene is. ncRVIS = Non-coding residual variation intolerance score; the higher the score is, the more intolerant to non-coding variation the gene is. posMapSNVs = Number of SNVs mapped to gene based on positional mapping. posMapMaxCADD = The maximum CADD score of mapped SNVs by positional mapping. minGwasP = The minimum P-value of mapped SNVs. IndSigSNVs = rsID of the independent significant SNVs that are in LD with the mapped SNVs. GenomicLocus = Index of genomic loci where mapped SNVs are from.

###### LVM

Figure 3.6: GWAS Manhattan plot for the LVM phenotype using BOLT-LMM. Red dashed line indicates the genome-wide significance level (p = 5E-8).

###### Genomic Risk Loci

Figure 3.7: **List of genomic risk loci from FUMA for the LVM BOLT-LMM GWAS.** uniqID = Unique ID of SNVs consists of chr:position:allele1:allele2 where alleles are alphabetically ordered. rsID = rsID of the top lead SNV for the given genomic locus. chr = Chromosome of top lead SNV. pos = Position of top lead SNV on hg19. p = P-value of top lead SNV. start = Start position of the locus. end = End position of the locus. nGWASSNVs = Number of the GWAS-tagged candidate SNVs within the genomic locus. nIndSigSNVs = Number of the independent significant SNVs in the genomic locus. IndSigSNVs = rsID of independent significant SNVs in the genomic locus. nLeadSNVs = The number of lead SNVs in the genomic locus. LeadSNVs = rsID of lead SNVs in the genomic locus.

###### Lead SNVs

Figure 3.8: **List of lead SNVs from FUMA for the LVM BOLT-LMM GWAS.** GenomicLocus = Index of assigned genomic locus (note: multiple independent lead SNVs can be assigned to the same genomic locus). uniqID = Unique ID of SNVs consists of chr:position:allele1:allele2 where alleles are alphabetically ordered. rsID = rsID of the SNV. chr = Chromosome. pos = Position on hg19. p = P-value from GWAS. nIndSigSNVs = Number of independent significant SNVs which are in LD of the lead SNV at \(r^2\) = 0.1. IndSigSNVs = rsID of independent significant SNVs which are in LD of the lead SNV at \(r^2\) = 0.1.

###### Independent Significant SNVs

Figure 3.9: **List of independent significant SNVs from FUMA for the LVM BOLT-LMM GWAS.** GenomicLocus = Index of assigned genomic locus (note: multiple independent lead SNVs can be assigned to the same genomic locus). uniqID = Unique ID of SNVs consists of chr:position:allele1:allele2 where alleles are alphabetically ordered. rsID = rsID of the SNV. chr = Chromosome. pos = Position on hg19. p = P-value from GWAS. nGWASSNVs = Number of GWAS-tagged SNVs which are in LD of the independent significant SNV given \(r^2\) = 0.6.

###### Mapped Genes

Figure 3.10: **List of mapped genes (via positional mapping) from FUMA for the LVM BOLT-LMM GWAS.** ensg = ENSG ID. symbol = Gene Symbol. chr = Chromosome. start = Starting position of the gene. end = Ending position of the gene. strand = Strand of the gene. type = Gene biotype from Ensembl. entrezID = entrez ID (if available). HUGO = HUGO (HGNC) gene symbol. pLI = pLI score (the probability of being loss-of-function intolerant) from ExAC database; the higher the score is, the more intolerant to loss-of-function mutations the gene is. ncRVIS = Non-coding residual variation intolerance score; the higher the score is, the more intolerant to non-coding variation the gene is. posMapSNVs = Number of SNVs mapped to gene based on positional mapping. posMapMaxCADD = The maximum CADD score of mapped SNVs by positional mapping. minGwasP = The minimum P-value of mapped SNVs. IndSigSNVs = rsID of the independent significant SNVs that are in LD with the mapped SNVs. GenomicLocus = Index of genomic loci where mapped SNVs are from.

##### PLINK

###### LVMi

Figure 3.11: GWAS Manhattan plot for the LVMi phenotype using Plink. Red dashed line indicates the genome-wide significance level (p = 5E-8).

###### Genomic Risk Loci

Figure 3.12: **List of genomic risk loci from FUMA for the LVMi Plink GWAS.** uniqID = Unique ID of SNVs consists of chr:position:allele1:allele2 where alleles are alphabetically ordered. rsID = rsID of the top lead SNV for the given genomic locus. chr = Chromosome of top lead SNV. pos = Position of top lead SNV on hg19. p = P-value of top lead SNV. start = Start position of the locus. end = End position of the locus. nGWASSNVs = Number of the GWAS-tagged candidate SNVs within the genomic locus. nIndSigSNVs = Number of the independent significant SNVs in the genomic locus. IndSigSNVs = rsID of independent significant SNVs in the genomic locus. nLeadSNVs = The number of lead SNVs in the genomic locus. LeadSNVs = rsID of lead SNVs in the genomic locus.

###### Lead SNVs

Figure 3.13: **List of lead SNVs from FUMA for the LVMi Plink GWAS.** GenomicLocus = Index of assigned genomic locus (note: multiple independent lead SNVs can be assigned to the same genomic locus). uniqID = Unique ID of SNVs consists of chr:position:allele1:allele2 where alleles are alphabetically ordered. rsID = rsID of the SNV. chr = Chromosome. pos = Position on hg19. p = P-value from GWAS. nIndSigSNVs = Number of independent significant SNVs which are in LD of the lead SNV at \(r^2\) = 0.1. IndSigSNVs = rsID of independent significant SNVs which are in LD of the lead SNV at \(r^2\) = 0.1.

###### Independent Significant SNVs

Figure 3.14: **List of independent significant SNVs from FUMA for the LVMi Plink GWAS.** GenomicLocus = Index of assigned genomic locus (note: multiple independent lead SNVs can be assigned to the same genomic locus). uniqID = Unique ID of SNVs consists of chr:position:allele1:allele2 where alleles are alphabetically ordered. rsID = rsID of the SNV. chr = Chromosome. pos = Position on hg19. p = P-value from GWAS. nGWASSNVs = Number of GWAS-tagged SNVs which are in LD of the independent significant SNV given \(r^2\) = 0.6.

###### Mapped Genes

Figure 3.15: **List of mapped genes (via positional mapping) from FUMA for the LVMi Plink GWAS.** ensg = ENSG ID. symbol = Gene Symbol. chr = Chromosome. start = Starting position of the gene. end = Ending position of the gene. strand = Strand of the gene. type = Gene biotype from Ensembl. entrezID = entrez ID (if available). HUGO = HUGO (HGNC) gene symbol. pLI = pLI score (the probability of being loss-of-function intolerant) from ExAC database; the higher the score is, the more intolerant to loss-of-function mutations the gene is. ncRVIS = Non-coding residual variation intolerance score; the higher the score is, the more intolerant to non-coding variation the gene is. posMapSNVs = Number of SNVs mapped to gene based on positional mapping. posMapMaxCADD = The maximum CADD score of mapped SNVs by positional mapping. minGwasP = The minimum P-value of mapped SNVs. IndSigSNVs = rsID of the independent significant SNVs that are in LD with the mapped SNVs. GenomicLocus = Index of genomic loci where mapped SNVs are from.

###### LVM

Figure 3.16: GWAS Manhattan plot for the LVM phenotype using Plink. Red dashed line indicates the genome-wide significance level (p = 5E-8).

###### Genomic Risk Loci

Figure 3.17: **List of genomic risk loci from FUMA for the LVM Plink GWAS.** uniqID = Unique ID of SNVs consists of chr:position:allele1:allele2 where alleles are alphabetically ordered. rsID = rsID of the top lead SNV for the given genomic locus. chr = Chromosome of top lead SNV. pos = Position of top lead SNV on hg19. p = P-value of top lead SNV. start = Start position of the locus. end = End position of the locus. nGWASSNVs = Number of the GWAS-tagged candidate SNVs within the genomic locus. nIndSigSNVs = Number of the independent significant SNVs in the genomic locus. IndSigSNVs = rsID of independent significant SNVs in the genomic locus. nLeadSNVs = The number of lead SNVs in the genomic locus. LeadSNVs = rsID of lead SNVs in the genomic locus.

###### Lead SNVs

Figure 3.18: **List of lead SNVs from FUMA for the LVM Plink GWAS.** GenomicLocus = Index of assigned genomic locus (note: multiple independent lead SNVs can be assigned to the same genomic locus). uniqID = Unique ID of SNVs consists of chr:position:allele1:allele2 where alleles are alphabetically ordered. rsID = rsID of the SNV. chr = Chromosome. pos = Position on hg19. p = P-value from GWAS. nIndSigSNVs = Number of independent significant SNVs which are in LD of the lead SNV at \(r^2\) = 0.1. IndSigSNVs = rsID of independent significant SNVs which are in LD of the lead SNV at \(r^2\) = 0.1.

###### Independent Significant SNVs

Figure 3.19: **List of independent significant SNVs from FUMA for the LVM Plink GWAS.** GenomicLocus = Index of assigned genomic locus (note: multiple independent lead SNVs can be assigned to the same genomic locus). uniqID = Unique ID of SNVs consists of chr:position:allele1:allele2 where alleles are alphabetically ordered. rsID = rsID of the SNV. chr = Chromosome. pos = Position on hg19. p = P-value from GWAS. nGWASSNVs = Number of GWAS-tagged SNVs which are in LD of the independent significant SNV given \(r^2\) = 0.6.

###### Mapped Genes

Figure 3.20: **List of mapped genes (via positional mapping) from FUMA for the LVM Plink GWAS.** ensg = ENSG ID. symbol = Gene Symbol. chr = Chromosome. start = Starting position of the gene. end = Ending position of the gene. strand = Strand of the gene. type = Gene biotype from Ensembl. entrezID = entrez ID (if available). HUGO = HUGO (HGNC) gene symbol. pLI = pLI score (the probability of being loss-of-function intolerant) from ExAC database; the higher the score is, the more intolerant to loss-of-function mutations the gene is. ncRVIS = Non-coding residual variation intolerance score; the higher the score is, the more intolerant to non-coding variation the gene is. posMapSNVs = Number of SNVs mapped to gene based on positional mapping. posMapMaxCADD = The maximum CADD score of mapped SNVs by positional mapping. minGwasP = The minimum P-value of mapped SNVs. IndSigSNVs = rsID of the independent significant SNVs that are in LD with the mapped SNVs. GenomicLocus = Index of genomic loci where mapped SNVs are from.

##### Stability Analysis

To further investigate the choice of GWAS method and phenotype, we conducted a deeper exploration into the similarities and differences between the BOLT-LMM and PLINK results as well as between the LVM and LVMi results in the stability analysis below. The primary goal of this stability analysis is to answer the following two questions:

1. **Are there large differences between the GWAS p-values from BOLT-LMM and PLINK?**

- Figure 3.21: For each SNV, we plot the p-value (log-transformed) obtained via BOLT-LMM (x-axis) and PLINK (y-axis) for each of the LVM (left) and LVMi (right) phenotypes. We see that the most significant GWAS hits tend to be stable, no matter the choice of algorithm, but as the strength of the association decreases, there is greater variation in the p-value between BOLT-LMM and PLINK. This is to say that there is more stability around the SNVs with the strongest associations with LVMi and less stability around the SNVs with moderate or weak associations with LVMi. Another reason for taking the union of the top 1000 SNVs from each GWAS algorithm in the lo-siRF pipeline was to allow these moderate to weak association SNVs the chance of being identified in a multivariate model (i.e., via iterative random forest).

2. **What are the FUMA GWAS findings that are stable across GWAS methods (BOLT-LMM and PLINK) and choice of phenotype (LVM and LVMi)?** As advocated by the stability principle (“S” in PCS) (Yu and Kumbier 2020; Yu 2013), we view stability across these arbitrary choices as a minimum requirement for making a scientific discovery.

- In Figure 3.22, we summarize the SNVs, identified by FUMA, to be the top lead SNVs (per genomic risk locus), lead SNVs, and/or independent significant SNVs for each GWAS method (BOLT-LMM and PLINK) and choice of phenotype (LVM and LVMi). Some takeaways here:
  - rs3045696, an intronic *TTN* variant, is the only stable top lead SNV, identified across all four GWAS runs.
  - Besides rs3045696, rs1873164, an intronic *CCDC141* variant, is the only stable lead SNV, identified by FUMA across all four GWAS runs.
  - rs7591091 (an intronic *CCDC141* variant with the highest frequency of occurrence in lo-siRF, seen later), along with rs967507, rs12622500, and rs10178003, were stably identified as independent significant SNVs by FUMA across all four GWAS runs.
  - Two other SNVs (rs62178977, rs73973171) were also identified as independent significant SNVs by FUMA using both BOLT-LMM and PLINK for the LVMi phenotype.
- In Figure 3.23, we summarize the genes that were positionally-mapped and prioritized by FUMA across the different GWAS methods (BOLT-LMM and PLINK) and choice of phenotype (LVM and LVMi).
  - The genes that were stably mapped by FUMA across all four GWAS runs are *PRKRA*, *DFNB59*, *FKBP7*, *PLEKHA3*, *TTN*, and *CCDC141*. We note that all of these genes are located near each other on chromosome 2 within ~600kb. The start position of PRKRA is 179296141 while the end position of CCDC141 is 179914813, with all other aforementioned genes in between.
  - No other genes were identified when using both BOLT-LMM and PLINK for the LVMi phenotype.

Figure 3.21: Comparison of the BOLT-LMM and PLINK GWAS p-values for each phenotype (LVM left, LVMi right). Each point represents a SNV with its BOLT-LMM p-value on the x-axis and its PLINK p-value on the y-axis.

Figure 3.22: Summary of annotated SNVs from FUMA across different GWAS methods (BOLT-LMM and PLINK) and across different choices of phenotype (LVMi and LVM).

Figure 3.23: Summary of mapped genes from FUMA across different GWAS methods (BOLT-LMM and PLINK) and across different choices of phenotype (LVMi and LVM).

#### GWAS-filtered SNVs

In this section, we briefly explore the selected 1405 SNVs that passed the GWAS filter for the LVMi phenotype. We make two observations that will impact how we interpret the lo-siRF fit downstream:

- There are strong correlations within blocks of neighboring SNVs due to linkage disequilibrium (LD) (Figure 3.24). This is a large contributing factor for why we ultimately extracted feature importances from the lo-siRF fit at the level of genetic loci, rather than for each individual SNV.
- There are large differences in the number of SNVs that passed the GWAS filter from each genetic loci (Figure 3.25). We hence must account for this in the lo-siRF locus-level importance score.

We will expand upon this locus-level importance score in a later section (see `Prioritization Step`).

Additional notes:

- Given the interest in *CCDC141* and *TTN* in our work and their close proximity to each other on chromosome 2, we point out a couple pairwise (Pearson) correlations of particular relevance:
  - \(| Cor(\text{rs7591091}, \text{ rs66733621}) | = 0.35\)
  - \(| Cor(\text{rs7591091}, \text{ rs3045696}) | = 0.34\)
  - rs7591091 is the top SNV from the CCDC141 locus in lo-siRF and is one of the top CCDC141 GWAS hits (in fact, is identified to be a stable independent significant SNV (see `GWAS Results Summary` tab)). rs3045696, an intronic *TTN* SNV, was stably identified to be a top lead SNV in the GWASes, and rs66733621 is the top SNV from the TTN locus in lo-siRF. This is to say that the correlation between the CCDC141 and TTN loci is moderate and may possibly suggest independent roles of CCDC141 and TTN in their detected interaction. A further discussion of this is provided in Q. Wang et al. (2023). [Note: we use the term *top SNV from a genetic locus in lo-siRF* to mean the SNV in the specified genetic locus with the highest frequency of occurrence across decision paths in the siRF fit.]

##### LVMi - Correlation Between GWAS-filtered SNVs

Figure 3.24: Correlation heatmap for the GWAS-filtered SNVs. We plot the magnitude of the Pearson correlation between all possible pairs of SNVs that passed the GWAS filter. Darker red indicates higher correlation between the two SNVs. SNVs along the axes are ordered according to genomic location. Loci with more than two constituent SNVs are labeled.

##### LVMi - Table of GWAS-filtered SNVs

Figure 3.25: Number of SNVs per genetic locus that passed the GWAS filter. Note: a semicolon is used to denote the intergenic region between two genes.

### 4 Binarization Step

#### Overview

In the next step of lo-siRF, we chose to binarize the (continuous) LVMi phenotype measurements into two categories: a low LVMi group and a high LVMi group before fitting the prediction models.

**Why did we choose to binarize?** One of our main motivating factors to pursue binarization revolved around the prediction check criteria in the PCS framework (Yu and Kumbier 2020). This predictability principle advocates for the use of prediction accuracy as a reality check. This is to say that at a minimum, models should fit the data well, as measured by prediction accuracy, before believing that the model is capturing something relevant to reality or the real-world phenomena under study. In particular, before we thought to binarize the phenotype, we first attempted to train prediction models using the GWAS-filtered SNVs to predict the original (continuous) LVMi phenotype measurements. While this seemed like the most natural next step, these prediction results illuminated an incredibly weak prediction signal, with validation \(R^2\) values hovering very close to 0 (see Figure 4.1 in the `Prediction Results without Binarization` tab). Given that an \(R^2\) value of 0 can be achieved by simply predicting the mean LVMi response, this raised the question of whether these prediction models are capturing any biologically-relevant signal in the data. Consequently, the interpretation of these models may not be an accurate representation of reality. Put differently, these models do not satisfy the prediction check criteria under the PCS framework.

Given these difficulties in accurately predicting LVMi, we drew inspiration from how scientists often reduce complex problems into simplified versions. Here, the complex version of our problem aims to understand how SNVs are predictive of one’s precise (continuous-valued) LVMi measurement. This is challenging, as demonstrated by the poor prediction accuracy discussed previously. On the other hand, arguably the most simplified version of this problem is to understand how SNVs are predictive of whether one has an extremely high or extremely low LVMi measurement. Intuitively, if there are in fact genetic differences between these two LV chamber states, then differentiating between the two groups — those with extremely high LVMi and those with extremely low LVMi — should be easier than the original regression problem (i.e., predicting the raw continuous-valued LVMi response). Indeed, we will see this to be the case, observing that the resulting classification models consistently performed better than random guessing unlike the original regression models. More specifically, we observed balanced classification accuracies \(>55\%\), area under the receiver operator characteristic (AUROC) \(>0.58\), and area under the precision-recall curve (AUPRC) \(>0.55\) for the constructed binary classification task (see the `Prediction Step` tab). This prediction performance instills greater trustworthiness that the interpretation of these models after binarization are more relevant to the biological phenomena than the case without binarization.

**How is the binarization done?** We transformed the original regression problem of predicting the raw continuous-valued LVMi response into a binary classification problem as follows. For particular choice of threshold \(x\), we partitioned the raw continuous-valued LVMi phenotype into a low, middle, and high LVMi group corresponding to the \([0, x]\), \((x, 1-x)\), and \([1-x, 1]\) quantile ranges, respectively. We then omitted the individuals in the middle quantile range and trained the prediction models to differentiate between the low and high LVMi individuals. However, due to the gender-specific biological variation of LVMi shown earlier in the exploratory data analysis (see `Choice of Phenotypic Data`), we performed this binarization procedure for males and females separately. That is, we computed the quantiles for males and females separately. Finally, given that this threshold is artificially introduced, we run the remainder of the lo-siRF pipeline using three different reasonable binarization thresholds (15%, 20%, 25%) that balance the improvement in prediction signal and amount of data lost. In particular, we note that if we had chosen a binarization threshold of 50% so that no data is thrown out, then the balanced classification accuracy of the best-performing prediction model (signed iterative random forest) falls to \(51%\), which is very similar to random guessing. In the end, we will consolidate the results that were stable across all three binarization thresholds (15%, 20%, 25%), described later.

**Limitations of binarization:** We recognize that this binarization procedure has limitations and that these limitations should be carefully considered in context of the problem under study. For instance, the utility of this binarization is highly dependent on whether understanding the differences between the low and high quantiles is relevant to the study aims. In our case, we believe that the connection between cardiac hypertrophy and those with high LVMi helps to justify the use of binarizing based on the low and high quantiles. If one does not expect a monotonic relationship between the genetic features and the phenotype, then binarization may not be appropriate. In addition, binarization also requires the choice of binarization threshold. This choice is almost certainly a subjective choice and thus necessitates a stability analysis, that is, analyzing the stability of findings across different choices of binarization thresholds. In our case, we only prioritize loci/interactions that were stably important across three different choices of binarization threshold (15%, 20%, and 25%). Finally, binarization also reduces the sample size and systematically omits individuals who are not in the tails of the phenotype distribution. As a result, there may be additional interactions that are missed in lo-siRF. While we do not view this as a major concern if the goal is to recommend a small set of reliable experimental targets, this limitation may certainly impact studies with different end goals.

We expand on the prediction modeling with the binarized LVMi phenotype in the next section (see `Prediction Step`).

#### Prediction Results without Binarization

Here, we present a summary of the prediction modeling results, where we use the GWAS-filtered SNVs to predict the raw LVMi phenotype measurements (i.e., without any binarization). Specifically, we measure the validation set prediction accuracy from several popular machine learning regression algorithms (kernel ridge regression, Lasso regression, ridge regression, random forest (RF), and signed iterative random forest (siRF)) across four different prediction performance metrics - the Pearson correlation, mean absolute error (MAE), R-squared, and root-mean-squared error (RMSE) between the observed LVMi response and the predicted LVMi response. We also visualize how the predicted responses differ across the different prediction models in the provided pair plots. Though RF appears to yield the best prediction performance relative to the other methods under consideration, the RF prediction performance is still very poor as both the correlation and R-squared metrics are very close to 0.

##### Prediction Accuracies

Figure 4.1: Summary of prediction performance on the validation set for the LVMi phenotype (without binarization) across various prediction methods and evaluation metrics.

##### Prediction Pair Plot

Figure 4.2: Pair plot of the predicted LVMi responses from various prediction models. Each point represents an individual in the validation set, and the coordinates represent the predicted LVMi response from two different prediction methods, which are specified by the x and y-axes labels in each subplot. Density distributions of the predicted LVMi responses are shown along the diagonal.

### 5 Prediction Step

After choosing to binarize the LVMi phenotype, we next fit various prediction models using the GWAS-filtered SNVs to predict the binarized LVMi phenotype and assess their validation prediction accuracy.

**Which models were chosen and why?** While there are many methods, each with their unique advantages and disadvantages, for detecting epistasis from data (e.g., see Niel et al. 2015 and references therein), we chose to focus on siRF (Kumbier et al. 2023) as our primary interaction search engine for various reasons.

1. Unlike many methods that make linear assumptions or require pre-specification of interaction order (i.e., the number of features involved in an interaction), siRF can identify *higher-order*, *nonlinear* Boolean interactions in a computationally-tractable manner without the need to specify the interaction order of interest.
2. Another advantage of siRF for detecting interactions is the resemblance between the localized thresholding behavior of its decision trees and the threshold (or on-off switch-like) behavior frequently observed in biomolecular interactions (Nelson, Lehninger, and Cox 2008). In other words, the form of the siRF model appears to be well-suited for the biological phenomena under study.
3. Furthermore, the prediction accuracy of the siRF model fit can be easily assessed. siRF also exhibited higher prediction accuracy than other commonly used prediction methods across almost all evaluation metrics examined and binarization thresholds (Figure 5.1). As discussed previously, prediction accuracy can serve as an indicator of whether or not the model is an accurate representation of reality. This prediction check is a necessary pre-requisite for interpreting the prediction model and is especially important in low-signal regimes such as this study.
4. siRF and its predecessor iRF have demonstrated great success in detecting reliable interactions in previous real-world studies, such as in epistasis-controlled hair color (Behr et al. 2020), obesity (Allen et al. 2022), schizophrenia (De Rosa et al. 2022), and predictive gene expression networks (Cliff et al. 2019; Walker et al. 2022).
5. Finally, due to the weak phenotypic signal and the strong correlations between neighboring SNVs due to linkage disequilibrium (Figure 3.24), we believed it was most appropriate to recommend interactions between genetic loci, rather than interactions betweeen individual SNVs. While it is not always clear how to perform this locus-level interaction search when the data is given at the SNV-level, we can build upon the siRF structure to aggregate feature importances across multiple SNVs using our new locus-level importance score, described in Q. Wang et al. (2023).

Though we are most interested in interpreting the siRF prediction model to identify candidate epistatic interactions, we also fit several other popular prediction models, including RF, L1-regularized logistic regression (Lasso), L2-regularized logistic regression (ridge), support vector machines (SVM), multilayer perceptron, AutoGluon TabularPredictor, and a polygenic risk score. These prediction models serve as baselines to ensure that siRF indeed fits the data well, relative to these other popular prediction models, and that siRF passes the prediction check in accordance with the PCS framework (Yu and Kumbier 2020).

**Summary of prediction results:** For all of the classification algorithms under investigation, we see that the validation prediction accuracy metrics, measured via classification accuracy, area under the receiver operator characteristic curve (AUROC), and area under the precision-recall curve (AUPRC), are all comfortably above the random-guessing baseline of 0.5 across three different binarization thresholds. This suggests that the prediction models are able to ascertain some phenotypic signal from the SNVs under study. That is, the SNVs used in the prediction models have some predictive power in differentiating those with high LVMi versus those with low LVMi. Moreover, siRF consistently yielded the highest predictive power across the various binarization thresholds and prediction metrics (with one exception being the classification accuracy for the 15% binarization threshold, where ridge is slightly better than siRF). From this, we conclude that siRF passed the prediction check and will proceed to interpret the siRF fit to recommend and prioritize interactions between genetic loci for downstream analyses and experiments.

Beyond the validation prediction accuracies, we also provide the confusion matrices, ROC and precision-recall curves, and pair plots comparing the predicted responses across methods for additional post-hoc exploration.

#### Prediction Accuracy Summary

Figure 5.1: Summary of prediction performance on the validation set for the binarized LVMi phenotype across various prediction methods, binarization thresholds, and evaluation metrics. Abbreviations: siRF, signed iterative random forest; RF, random forest; Lasso, L1-regularized logistic regression; Ridge, L2-regularized logistic regression; SVM, support vector machine; MLP, multilayer perceptron; Autogluon (medium/good), Autogluon TabularPredictor fitted with the medium/good quality preset; PGS, polygenic risk score.

#### Binarization Threshold = 0.15

##### Confusion Tables

Figure 5.2: Confusion matrix from various prediction models for predicting the binarized LVMi phenotype (binarization threshold = 0.15).

##### ROC/PR Curves

Figure 5.3: ROC curve for predicting the binarized LVMi (threshold = 0.15) across various prediction methods. TPR = true positive rate; FPR = false positive rate.

Figure 5.4: Precision-recall curve for predicting the binarized LVMi (threshold = 0.15) across various prediction methods.

##### Prediction Pair Plot

Figure 5.5: Pair plots of the predicted binarized LVMi responses (threshold = 0.15) across various prediction methods. Each point represents an individual in the validation set, and the coordinates represent the predicted probability of having high LVMi from two different prediction methods, which are specified by the x and y-axes labels in each subplot. Density distributions of the predicted probabilities are shown along the diagonal.

#### Binarization Threshold = 0.2

##### Confusion Tables

Figure 5.6: Confusion matrix from various prediction models for predicting the binarized LVMi phenotype (binarization threshold = 0.2).

##### ROC/PR Curves

Figure 5.7: ROC curve for predicting the binarized LVMi (threshold = 0.2) across various prediction methods. TPR = true positive rate; FPR = false positive rate.

Figure 5.8: Precision-recall curve for predicting the binarized LVMi (threshold = 0.2) across various prediction methods.

##### Prediction Pair Plot

Figure 5.9: Pair plots of the predicted binarized LVMi responses (threshold = 0.2) across various prediction methods. Each point represents an individual in the validation set, and the coordinates represent the predicted probability of having high LVMi from two different prediction methods, which are specified by the x and y-axes labels in each subplot. Density distributions of the predicted probabilities are shown along the diagonal.

#### Binarization Threshold = 0.25

##### Confusion Tables

Figure 5.10: Confusion matrix from various prediction models for predicting the binarized LVMi phenotype (binarization threshold = 0.25).

##### ROC/PR Curves

Figure 5.11: ROC curve for predicting the binarized LVMi (threshold = 0.25) across various prediction methods. TPR = true positive rate; FPR = false positive rate.

Figure 5.12: Precision-recall curve for predicting the binarized LVMi (threshold = 0.25) across various prediction methods.

##### Prediction Pair Plot

Figure 5.13: Pair plots of the predicted binarized LVMi responses (threshold = 0.25) across various prediction methods. Each point represents an individual in the validation set, and the coordinates represent the predicted probability of having high LVMi from two different prediction methods, which are specified by the x and y-axes labels in each subplot. Density distributions of the predicted probabilities are shown along the diagonal.

### 6 Prioritization Step

#### Overview

Having passed the prediction check, we finally proceed to interpret the siRF fit in the last step of lo-siRF. In this step, we developed a novel stability-driven feature importance score to prioritize both genetic loci and interactions between genetic loci from siRF. We refer to Q. Wang et al. (2023) for the detailed description of this new importance score and here provide brief insights into the motivation and intuition behind this method.

**Main idea:** At a high-level, this new importance score (1) aggregates weak, unstable SNV-level importances from siRF into stronger, more stable locus-level importances and (2) can be leveraged to evaluate both the marginal importance of a genetic locus in siRF as well as the importance of higher-order interactions between genetic loci in siRF. The need for this locus-level feature importance score stems from two main reasons: the high correlation between SNVs and the weak phenotypic signal. Though the prediction models (including siRF) are all trained using SNV data, it is often very challenging to interpret the importance of an individual SNV from these models due to the high correlation between SNVs (Toloşi and Lengauer 2011; Hooker, Mentch, and Zhou 2021), which we observed in our previous exploratory data analysis (see `Choice of Phenotypic Data` tab). To further complicate matters, the weak phenotypic signal increases the instability of existing feature importance methods in that we see the feature rankings change drastically when using different bootstrap training samples. Given these challenges with SNV-level feature importances, it is then no surprise that siRF, out-of-the-box, does not find any stable interactions between SNVs (i.e., the siRF stability score is around 0 for all interactions between SNVs). To address these challenges, our new feature importance score leverages the correlation structure between SNVs, groups SNVs into genetic loci, and aggregates weak, unstable SNV-level importances from siRF into stronger, more stable locus-level importances.

**Motivation behind new feature importance score:** To better understand the motivation and construction of our new locus-level feature importance score, it can be helpful to first review the original importance score used in siRF (Kumbier et al. 2023). Briefly, in the original version of siRF, the importance of a signed interaction is based upon *how stable or frequently it occurs* in decisions paths across the random forest – that is, based upon the number of decision paths for which every signed feature in the signed interaction was used in at least one decision split (see Kumbier et al. (2023) for details). Following a similar logic, one natural path forward to calculate a locus-level importance from a forest that was fit on SNV data is as follows:

1. Take the fitted forest, where each decision split used some SNV to make the split.
2. Map each SNV used in a decision split to its corresponding genetic loci (e.g., using annotation software like ANNOVAR (K. Wang, Li, and Hakonarson 2010)).
3. Compute the importance of a signed locus-level interaction based upon the frequency of occurrence of the interaction as before, but using the mapped genetic loci in lieu of the SNVs. That is, compute the number of decision paths for which every signed locus in the signed locus-level interaction was used in at least one decision split.

However, this *frequency of occurrence* measure is heavily tied to the number of SNVs that belong to each genetic locus. A genetic locus that consists of more SNVs will naturally have a higher frequency of occurrence than a genetic locus with only a few SNVs even though both genetic loci may be equally important (or unimportant). In other words, the original siRF importance score, measuring the frequency of occurrence, is not comparable across genetic loci. This bias is problematic given that we have previously seen some genetic loci may consist of only 1 SNV while other genetic loci have >50 SNVs (see `GWAS-filtered SNVs` tab in the Dimension Reduction Step).

**Intuition behind new feature importance score:** To alleviate this bias, we leverage the idea of a *local* feature importance score, where we compute a *per-individual* score that measures the importance of a locus/interaction for making the prediction of *each* individual. This is in contrast to a *global* feature importance score, which is a measure of importance for the entire group of individuals under study. While a global feature importance score gives rise to a *single* score for the entire population under study, we can compute a local feature importance score for *each* individual in the population. By computing a local feature importance score for each individual (i.e., this is the local stability importance (LSI) score from Q. Wang et al. (2023)), we can compare the importance of the locus/interaction across *individuals* and avoid the across-*loci* comparison that was problematic in the aforementioned discussion. This comparison is done via a permutation test in Q. Wang et al. (2023). Moreover, since we are comparing the local feature importances between individuals *within* or *per* loci/interaction in this permutation test, the local feature importances are all measured on the same scale and comparable, thereby mitigating the bias towards genetic loci with more SNVs.

**How to aggregate SNVs:** We mapped each SNV to its corresponding genetic locus using ANNOVAR (K. Wang, Li, and Hakonarson 2010) according to the hg19 refSeq Gene annotations. Given our ultimate goal of recommending/prioritizing genetic loci and interactions between loci for downstream analyses and experimental validation, we believe this choice provides an appropriate starting point to obtain actionable recommendations. However, we note that there are many alternative ways of aggregating SNVs. For example, it is possible to aggregate SNVs according to LD/haplotype blocks or to include additional gene-based or functional annotations in the aggregation scheme.

**Organization:** Under the `SNV-level Importances` tab, we provide the SNV-level feature importances from the fitted prediction algorithms (described in the `Prediction Step` section) for completeness. These SNV-level feature importances are very unstable across methods and across bootstrap perturbations for the reasons discussed above. We then summarize the results from the lo-siRF pipeline including the rankings of genetic loci and interaction between genetic loci using our new locus-level importance score under the `Summary of lo-siRF Results` tab. A final technical note on the advantages of incorporating signed information from siRF in this feature importance computation is provided under the `Advantages of Signed Information` tab.

#### SNV-level Feature Importances

Having observed that the aforementioned classification algorithms do have some predictive power to predict those with high versus low LVMi, a natural next step is to investigate which SNVs were used in these prediction models. Below, we highlight the top 100 SNVs, ranked by importance in each of the prediction models. For completeness, we provide the SNV importances for the unbinarized LVMi (regression) problem as well as the binarized LVMi (classification) problem across the three different binarization thresholds.

**How is importance defined?**

- For ridge and Lasso, the importance of an SNV is defined as the magnitude of its coefficient in the prediction model.
- For RF, the importance of an SNV is determined by the mean decrease in impurity (MDI) score.
- For siRF, the importance of an SNV is determined by the MDI from the forest in the final iteration.
- Due to the computational complexity, we did not compute the SNV importances from the SVM fit as there is no built-in model-based feature importance method (in R), and any permutation-based approach would be computationally expensive.

**A word of caution:** When interpreting these SNV importances, it is crucial to keep in mind that there are very large, strongly correlated blocks of SNVs within this data, which often renders the feature rankings unmeaningful and unstable (Toloşi and Lengauer 2011; Hooker, Mentch, and Zhou 2021). In particular, it is well-known that the MDI variable importance metric used in RF/iRF/siRF is biased against correlated features (Hooker, Mentch, and Zhou 2021). In other words, SNVs within large LD blocks tend to have artificially low MDI values. Given these idiosyncrasies, it is unsurprising that SNVs from TTN and CCDC141 – two genetic loci with many SNVs under LD – do not have high importances according to MDI. We provide these SNV importance tables only for completeness and to partially motivate the need to aggregate SNV-level importances into locus-level importances in lo-siRF.

##### Unbinarized (Regression)

###### Lasso

Figure 6.1: Lasso - Top 100 SNVs, ranked by importance, for predicting LVMi.

###### Ridge

Figure 6.2: Ridge - Top 100 SNVs, ranked by importance, for predicting LVMi.

#### RF

Figure 6.3: RF - Top 100 SNVs, ranked by importance, for predicting LVMi.

###### siRF

Figure 6.4: siRF - Top 100 SNVs, ranked by importance, for predicting LVMi.

##### Binarization Threshold = 0.15

###### Lasso

Figure 6.5: Lasso - Top 100 SNVs, ranked by importance, for predicting the binarized LVMi (threshold = 0.15).

###### Ridge

Figure 6.6: Ridge - Top 100 SNVs, ranked by importance, for predicting the binarized LVMi (threshold = 0.15).

#### RF

Figure 6.7: RF - Top 100 SNVs, ranked by importance, for predicting the binarized LVMi (threshold = 0.15).

###### siRF

Figure 6.8: siRF - Top 100 SNVs, ranked by importance, for predicting the binarized LVMi (threshold = 0.15).

##### Binarization Threshold = 0.2

###### Lasso

Figure 6.9: Lasso - Top 100 SNVs, ranked by importance, for predicting the binarized LVMi (threshold = 0.2).

###### Ridge

Figure 6.10: Ridge - Top 100 SNVs, ranked by importance, for predicting the binarized LVMi (threshold = 0.2).

#### RF

Figure 6.11: RF - Top 100 SNVs, ranked by importance, for predicting the binarized LVMi (threshold = 0.2).

###### siRF

Figure 6.12: siRF - Top 100 SNVs, ranked by importance, for predicting the binarized LVMi (threshold = 0.2).

##### Binarization Threshold = 0.25

###### Lasso

Figure 6.13: Lasso - Top 100 SNVs, ranked by importance, for predicting the binarized LVMi (threshold = 0.25).

###### Ridge

Figure 6.14: Ridge - Top 100 SNVs, ranked by importance, for predicting the binarized LVMi (threshold = 0.25).

#### RF

Figure 6.15: RF - Top 100 SNVs, ranked by importance, for predicting the binarized LVMi (threshold = 0.25).

###### siRF

Figure 6.16: siRF - Top 100 SNVs, ranked by importance, for predicting the binarized LVMi (threshold = 0.25).

#### Summary of lo-siRF Results

Finally, we summarize the prioritization results from the lo-siRF pipeline using our new importance score to rank genetic loci and interactions between genetic loci. In this new importance score, we (1) computed a local (i.e., on a per-individual basis) stability importance score that aggregates weak SNV-level importances into stronger locus-level importances using the siRF fit and (2) conducted a permutation test to assess whether the local stability importance scores for a given locus/interaction are different between individuals with high and low LVMi (conditioned on the rest of the fitted forest). The larger the difference in local stability importance scores between those with high versus low LVMi, the greater the importance of the locus/interaction for predicting LVMi. For details on the new importance score, we refer to Q. Wang et al. (2023).

Below, we recap the siRF prediction accuracy for predicting the binarized LVMi across the three binarization thresholds (15%, 20%, 25%) (Figure 6.17). Then, in Figure 6.18, we summarize the prioritization rankings of genetic loci and interaction between loci according to the new importance score. We provide the nominal permutation p-values along with the permutation test statistic, that is, the difference in local feature stability scores between the high and low LVMi groups, with the standard deviation of this permutation distribution in parentheses.

Note that the permutation test was not performed for all loci/interactions (details in Q. Wang et al. (2023)). Given our primary aim of generating reliable hypotheses for follow-up experimental validation, we chose to only test loci/interactions that were *predictive* and *stable* in the siRF fit, following the PCS framework for veridical data science (Yu and Kumbier 2020). Specifically, for each binarization threshold, we only tested:

- the top 25 loci with the highest average local stability importance scores, or in other words, those that occurred most stably/frequently (and thus likely predictive) in the siRF fit, and
- the interactions between loci that were identified by siRF and achieved a stability score > 0.5, stability score for mean increase in precision > 0, and stability score for feature selection dependence > 0. We note that in addition to the explicit stability checks here, the requirement on the mean increase in precision serves as a predictability check, and the requirement on the feature selection dependence serves to differentiate marginal additive effects from non-additive interaction effects (see Kumbier et al. (2023) for details).

By implementing this additional predictability and stability check, we are requiring that the prioritized loci/interactions meet a higher standard, with the hope that this ultimately generates more reliable (albeit more conservative) experimental recommendations.

In addition to the provided summary of results,

- Under the `siRF Interaction Table` tab, we provide the full siRF locus-level interaction table output for each binarization threshold (Figures 6.19, 6.21, and 6.23). This table includes several metrics for evaluating the stability and strength of the interaction. For details on these metrics, we refer to Kumbier et al. (2023).
  - For comparison, we also provide the analogous siRF output for interactions at the SNV level (Figures 6.20, 6.22, and 6.24). Here, we see that if we do not perform the SNV-to-locus aggregation, the stability scores for the SNV-level interactions are very close to 0. This means that the siRF-identified SNV-level interactions rarely appear when siRF is refitted using different bootstrapped training samples. As discussed previously, this instability is partially due to the correlation among SNVs and the weak phenotypic signal. This high instability also motivates the need for an alternative approach via our new importance score.
- Under the `Local Stability Importance Plots - Interactions` and `Local Stability Importance Plots - Loci`, we provide visualizations of the density distribution of local stability importance scores between the low and high LVMi groups as well as the frequency distribution of SNVs used in the siRF fit (i.e., which SNVs occurred most frequently in the decision paths across the siRF fit).
  - In the density distribution of local stability importance scores, observable differences between the low and high LVMi groups are evidence that the genetic locus or interaction between loci is important for differentiating between individuals with high versus low LVMi. The larger the difference, the greater the importance of the locus or interaction between loci.
  - In the SNV frequency distributions, we show the frequency of occurrence for each SNV or SNV pair in the siRF fit. This is the number of decision paths for which the SNV or SNV pair appeared in the siRF fit. SNVs or SNV pairs with the highest number of occurrences in the RF decision paths (y-axis) contributed the most to the overall locus/interaction’s importance in the siRF fit.

##### Summary

Figure 6.17: Summary of the validation prediction performance from siRF across various LVMi binarization thresholds.

Figure 6.18: Summary of the lo-siRF permutation test results across various LVMi binarization thresholds including the test statistic (i.e., difference in local feature stability scores between the high and the low LVMi groups (SD in parentheses) and the resulting nominal p-values. Blank cells indicate that the locus/interaction was not selected for testing for the given binarization threshold. Loci/interactions are ranked according to the mean nominal p-value across all three binarization thresholds. Interactions between genetic loci are highlighted in blue.

##### siRF Interaction Table

#### 0.15

Figure 6.19: Summary of the siRF locus-level interaction table output for the binarized LVMi phenotype (binarization threshold = 0.15). For all metrics excluding the nominal lo-siRF p-value, higher values indicate a stronger interaction. Details regarding these metrics in the siRF interaction table output can be found in Kumbier et al. (2018).

Figure 6.20: Summary of the siRF SNV-level interaction table output for the binarized LVMi phenotype (binarization threshold = 0.15). Stability scores for all interactions are close to 0, suggesting unreliable SNV-level interactions. Details regarding these metrics in the siRF interaction table output can be found in Kumbier et al. (2018).

#### 0.2

Figure 6.21: Summary of the siRF locus-level interaction table output for the binarized LVMi phenotype (binarization threshold = 0.2). For all metrics excluding the nominal lo-siRF p-value, higher values indicate a stronger interaction. Details regarding these metrics in the siRF interaction table output can be found in Kumbier et al. (2018).

Figure 6.22: Summary of the siRF SNV-level interaction table output for the binarized LVMi phenotype (binarization threshold = 0.2). Stability scores for all interactions are close to 0, suggesting unreliable SNV-level interactions. Details regarding these metrics in the siRF interaction table output can be found in Kumbier et al. (2018).

#### 0.25

Figure 6.23: Summary of the siRF locus-level interaction table output for the binarized LVMi phenotype (binarization threshold = 0.25). For all metrics excluding the nominal lo-siRF p-value, higher values indicate a stronger interaction. Details regarding these metrics in the siRF interaction table output can be found in Kumbier et al. (2018).

Figure 6.24: Summary of the siRF SNV-level interaction table output for the binarized LVMi phenotype (binarization threshold = 0.25). Stability scores for all interactions are close to 0, suggesting unreliable SNV-level interactions. Details regarding these metrics in the siRF interaction table output can be found in Kumbier et al. (2018).

##### Local Stability Importance Plots - Interactions

#### 0.15

Figure 6.25: Distribution of local stability importance scores between the high and low LVMi groups (binarization threshold = 0.15) for each signed interaction between loci (specified by subplot) that passed the siRF stability filter. The nominal p-value from the permutation test, assessing distributional differences between the high and low LVMi groups, is provided in the top right corner of each subplot.

Figure 6.26: Frequency of occurrence of SNV-SNV combinations in the siRF fit for the binarized LVMi phenotype (binarization threshold = 0.15). SNV-SNV combinations with the highest number of occurrences in the siRF decision paths contribute the most to the overall locus-level interaction importance in the siRF fit.

#### 0.2

Figure 6.27: Distribution of local stability importance scores between the high and low LVMi groups (binarization threshold = 0.2) for each signed interaction between loci (specified by subplot) that passed the siRF stability filter. The nominal p-value from the permutation test, assessing distributional differences between the high and low LVMi groups, is provided in the top right corner of each subplot.

Figure 6.28: Frequency of occurrence of SNV-SNV combinations in the siRF fit for the binarized LVMi phenotype (binarization threshold = 0.2). SNV-SNV combinations with the highest number of occurrences in the siRF decision paths contribute the most to the overall locus-level interaction importance in the siRF fit.

#### 0.25

Figure 6.29: Distribution of local stability importance scores between the high and low LVMi groups (binarization threshold = 0.25) for each signed interaction between loci (specified by subplot) that passed the siRF stability filter. The nominal p-value from the permutation test, assessing distributional differences between the high and low LVMi groups, is provided in the top right corner of each subplot.

Figure 6.30: Frequency of occurrence of SNV-SNV combinations in the siRF fit for the binarized LVMi phenotype (binarization threshold = 0.25). SNV-SNV combinations with the highest number of occurrences in the siRF decision paths contribute the most to the overall locus-level interaction importance in the siRF fit.

##### Local Stability Importance Plots - Loci

#### 0.15

Figure 6.31: Distribution of local stability importance scores between the high and low LVMi groups (binarization threshold = 0.15) for each signed locus (specified by subplot). Here, we show only the top 25 signed loci, ranked by their average local stability importance scores. The nominal p-value from the permutation test, assessing distributional differences between the high and low LVMi groups, is provided in the top right corner of each subplot.

Figure 6.32: Frequency of occurrence of SNVs in the siRF fit for the binarized LVMi phenotype (binarization threshold = 0.15). SNVs with the highest number of occurrences in the siRF decision paths contribute the most to the overall locus-level importance in the siRF fit. SNVs are ordered on the x-axis by genomic location.

#### 0.2

Figure 6.33: Distribution of local stability importance scores between the high and low LVMi groups (binarization threshold = 0.2) for each signed locus (specified by subplot). Here, we show only the top 25 signed loci, ranked by their average local stability importance scores. The nominal p-value from the permutation test, assessing distributional differences between the high and low LVMi groups, is provided in the top right corner of each subplot.

Figure 6.34: Frequency of occurrence of SNVs in the siRF fit for the binarized LVMi phenotype (binarization threshold = 0.2). SNVs with the highest number of occurrences in the siRF decision paths contribute the most to the overall locus-level importance in the siRF fit. SNVs are ordered on the x-axis by genomic location.

#### 0.25

Figure 6.35: Distribution of local stability importance scores between the high and low LVMi groups (binarization threshold = 0.25) for each signed locus (specified by subplot). Here, we show only the top 25 signed loci, ranked by their average local stability importance scores. The nominal p-value from the permutation test, assessing distributional differences between the high and low LVMi groups, is provided in the top right corner of each subplot.

Figure 6.36: Frequency of occurrence of SNVs in the siRF fit for the binarized LVMi phenotype (binarization threshold = 0.25). SNVs with the highest number of occurrences in the siRF decision paths contribute the most to the overall locus-level importance in the siRF fit. SNVs are ordered on the x-axis by genomic location.

#### Advantages of Signed Information

Though the signed information is not pertinent to our goal of prioritizing/recommending candidate markers for experiments, the signed information from siRF provides more granular information that can improve our interpretation of the fit. To see this, suppose that we did not keep track of the sign of the feature when splitting (i.e., that we do not keep track of whether we split on low or high values of the feature). Suppose also that we would like to measure the importance of feature \(X\_1\), which was used as the first (root) split in tree \(T\). In the first step in our new importance score computation, we compute the local stability importance (LSI) score of \(X\_1\) for each individual \(i\). If we do not keep track of the sign of the \(X\_1\) split, then \(X\_1\) appears on every individual’s decision path so that \(LSI\_T(X\_1, i) = 1\) for every individual \(i\). In the next step of our new importance score computation, we perform a permutation test to assess whether there is a difference in the LSI scores for individuals with high LVMi and those with low LVMi. Since \(LSI\_T(X\_1, i) = 1\) for every \(i\), this permutation test will return a nominal p-value of 1. This suggests that \(X\_1\) is an unimportant feature in this model. However, this is counterintuitive given that \(X\_1\) was split on first in the tree and should be considered one of the most important features in this tree. By keeping track of the signed information of the splits as done in siRF, we can utilize this more granular information to mitigate the issue discussed above and improve our interpretation of the siRF fit.

Moreover, to facilitate the interpretation of this signed information, it can be beneficial to aggregate SNV features that are *positively* correlated (as opposed to negatively correlated) into a single locus (or group). Since the encoding of SNVs as a value taking on 0, 1, or 2 is rather arbitrary (e.g., a value of 0 can be re-encoded to take on the value of 2 by changing the reference allele), we can choose the encoding of the SNV to try to avoid strong negative correlations within a locus. More specifically, for each genetic locus which consists of SNVs \(X\_1, \ldots, X\_g\), we choose to “flip” the encoding of the SNV \(X\_j\) (\(j = 2, \ldots, g\)) if the Pearson correlation between the observed values of \(X\_j\) and \(X\_1\) is below -0.15. Here, we use the term “flip” to mean the application of the function that maps 0 to 2, 1 to 1, and 2 to 0. This correlation threshold of -0.15 was chosen by visually inspecting the resulting pairwise correlations between \(X\_i\) and \(X\_j\) for all \(i, j = 1, \ldots, g\) and checking that these pairwise correlations are not strongly negative. However, while we focus on the lo-siRF prioritization results using this correlation threshold of -0.15, the results appear to be robust to other reasonable correlation threshold choices (e.g., -0.05, -0.25). Further analysis and improvements to this procedure are very interesting for future work.

### 7 Non-hypertensive Analysis

#### Overview

The lo-siRF analysis thus far has not accounted for the possibility of pleiotropy, particular between LV mass and blood pressure given the known connections between LV hypertrophy and hypertension (Yildiz et al. 2020). To evaluate this possibility of pleiotropic effects of the identified variants on both LV mass and blood pressure, we conducted two analyses.

1. We assessed the marginal association between each of the lo-siRF-prioritized variants and hypertension (see `Association between SNVs and Hypertension` tab).
2. We repeated the lo-siRF analysis using only the subset of UK Biobank individuals who were not reported to have hypertension (see `Lo-siRF Results Summary` tab).

Here, we define hypertensive diseases as anyone with self-reported hypertension, high blood pressure as diagnosed by a doctor, or any ICD10 billing code diagnosis in I10-I16. Out of the 29,661 UK Biobank participants in the original lo-siRF analysis, 7,371 individuals had hypertension, leaving 22,290 individuals for the non-hypertensive analysis.

#### Association between SNVs and Hypertension

To assess their marginal association with hypertension, we fit a separate logistic regression model for each of the 1405 GWAS-filtered SNVs, regressing hypertension (i.e., a binary indicator of whether or not one has hypertension) on the genotype of the SNV, while adjusting for the first five principal components of ancestry, sex, age, height, and body weight.

Overall, we found that none of the 1405 SNVs met the genome-wide (p < 5E-8) nor the suggestive (p < 1E-5) significance level (Figure 7.1 and Figure 7.2). Given our interest in the lo-siRF-prioritized interactions, which involve the *TTN*, *CCDC141*, *IGF1R*, and *LOC157273;TNKS* loci, we highlight the SNVs in each locus with the smallest p-values:

- *TTN*: rs35112591 (p = 0.0328)
- *CCDC141*: rs6725028 (p = 0.0403)
- *IGF1R*: rs62024491 (p = 0.376)
- *LOC157273;TNKS*: rs73185221 (0.0491)

The top lo-siRF-prioritized variants in each loci (see Figure 2f in Q. Wang et al. (2023)) exhibited even weaker marginal associations with hypertension;

- *TTN*: rs66733621 (p = 0.776)
- *CCDC141*: rs7591091 (p = 0.446)
- *IGF1R*: rs62024491 (p = 0.376)
- *LOC157273;TNKS*: rs6999852 (0.304)

We note however that the *MIR588;RSPO3* locus with lead SNV rs2022479 gave the smallest p-value of 5E-5. This, in combination with the findings in the non-hypertension lo-siRF analysis next, may suggest a possible contribution of the MIR588;RSPO3 locus on mediating LV hypertrophy through regulating blood pressure.

Figure 7.1: Manhattan plot for the association between the 1405 GWAS-filtered SNVs used in the original lo-siRF analysis and hypertension.

Figure 7.2: **List of top 100 SNVs (out of the 1405 GWAS-filtered SNVs used in the original lo-siRF analysis) with the highest association with hypertension.** rsID = rsID of the SNV. Chr = Chromosome. Pos = Position on hg19. Lo-siRF Locus = assigned locus of the variant in the original lo-siRF analysis. p = P-value measuring the association between the SNV and hypertension.

#### Non-hypertensive Lo-siRF Results Summary

Using the same set of 1405 GWAS-filtered SNVs as in the original lo-siRF analysis, we repeated steps 2-4 of the lo-siRF analysis using only the non-hypertension UK Biobank cohort.

- In Figure 7.3, we confirm that the siRF fit on the non-hypertensive cohort performs better than random guessing and satisfies the prediction check according to the PCS framework (Yu and Kumbier 2020). We thus choose to proceed with interpreting the siRF fit.
- Overall, we found that the top 3 interactions between loci from our original lo-siRF analysis — namely, *CCDC141-IGF1R*, *CCDC141-TTN*, and *CCDC141-LOC157273;TNKS* — were again prioritized by lo-siRF when excluding the hypertensive individuals. That is, each of the aforementioned interactions yielded a nominal lo-siRF p-value less than our pre-specified threshold of 0.1 (in fact, all were < 0.01) for each of the three binarization thresholds (see Figure 7.4 below). This is to say that the top lo-siRF-prioritized interactions are stable with respect to the inclusion or exclusion of hypertensive individuals and provides evidence that these interactions are not solely mediating LV hypertrophy through the regulation of blood pressure.
- All of the previously prioritized loci, with the exception of the *MIR588;RSPO3* locus, were also prioritized in this non-hypertensive analysis. While the nominal lo-siRF p-value for the *MIR588;RSPO3* locus was 0.019 and 0.007 for the 20% and 25% binarization thresholds, the nominal lo-siRF p-value was 0.187 (> 0.1 threshold) for the 15% binarization threshold and thus deemed not stable and was not prioritized in the non-hypertensive lo-siRF analysis. Though the *MIR588;RSPO3* locus almost met the prioritization criteria here, this result, combined with the suspect marginal association results, suggests a possibility of pleiotropic locus contributing to variations of both blood pressure and LV mass.
- We provide the full list of loci and interactions between loci from the non-hypertensive lo-siRF analysis in Figure 7.5. This list includes many loci and interactions that were not stable in the original lo-siRF analysis.

Figure 7.3: Summary of the validation prediction performance from siRF on the non-hypertensive cohort across various LVMi binarization thresholds.

Figure 7.4: Summary of the non-hypertensive lo-siRF permutation test results across various LVMi binarization thresholds including the test statistic (i.e., difference in local feature stability scores between the high and the low LVMi groups (SD in parentheses) and the resulting nominal p-values. Results are shown for the loci/interactions that were previously prioritized in the original lo-siRF analysis (including both hypertensive and non-hypertensive individuals). Loci/interactions are ranked according to the mean nominal p-value across all three binarization thresholds. Interactions between genetic loci are highlighted in blue. This table is a subset of the table below.

Figure 7.5: Summary of the non-hypertensive lo-siRF permutation test results across various LVMi binarization thresholds including the test statistic (i.e., difference in local feature stability scores between the high and the low LVMi groups (SD in parentheses) and the resulting nominal p-values. Blank cells indicate that the locus/interaction was not selected for testing for the given binarization threshold. Loci/interactions are ranked according to the mean nominal p-value across all three binarization thresholds. Interactions between genetic loci are highlighted in blue.

### 8 Comparison to Existing Methods

#### Epistasis Detection Methods

In this section, we compare the lo-siRF-prioritized interactions between loci to those interactions prioritized by other existing epistasis detection methods, namely, an exhaustive pairwise interaction search via linear (or logistic) regression, MAPIT (Crawford et al. 2017), and a locus-level version of MAPIT where we combined MAPIT and gene set enrichment analysis (GSEA) (Subramanian et al. 2005; Mootha et al. 2003).

##### Regression-based pairwise interaction scan

Below, we present the results from conducting an exhaustive pairwise interaction scan using linear (or logistic) regression over all GWAS-filtered SNVs with a minor allele frequency > 0.05.
In this exhaustive interaction search, there are four different transformations of the LVMi response/phenotype that are of interest — namely, the rank-based inverse normal-transformed (RINT) LVMi (fitted using linear regression) and the binarized LVMi at three different thresholds (0.15, 0.2, and 0.25) (fitted using logistic regression).
Details regarding the methodology can be found in Q. Wang et al. (2023).

- In Figure 8.1, we examined the stability of the top ranked SNV-SNV interactions (i.e., those with the smallest p-values for the interaction term) across the different choices of LVMi response (i.e., RINT or one of the binarized versions). While there is some positive correlation across the different binarization runs, we found that there is very little correlation between the prioritized interactions when using the RINT-transformed LVMi versus the binarized LVMi as the response.
  - We note that the exhaustive interaction search was also conducted using all 1405 GWAS-filtered SNVs without the minor allele frequency filter. However, this led to even greater instability. We hence omit these results and focus on the results after restricting to SNVs with a minor allele frequency > 0.05.
- In Figure 8.2, we provide a table of the SNV-SNV interactions and their corresponding interaction term p-values for each choice of LVMi response. We show only those SNV-SNV interactions with a p-value < 0.01 for at least one of the LVMi responses.
  - We note that none of the SNV-SNV interactions met the \(\alpha = 0.05\) significance threshold after performing a Bonferroni multiple testing adjustment.
  - Still, one could in theory use the p-values for ranking SNV-SNV interactions. For the unbinarized RINT-transformed LVMi response, the top interaction was between rs10227393 (chromosome 7; *TPK1;CNTNAP2* locus) and rs8076248 (chromosome 17; *CFAP52* locus). For the binarized LVMi response using both the 15% and 25% thresholds, the top interaction was between rs3176323 (chromosome 6; *CDKN1A* locus) and rs12795076 (chromosome 11; *KIRREL3* locus). To our knowledge, these loci are not known to be closely related to hypertrophy. Only for the binarized LVMi response with the 20% threshold do we obtain a top interaction involving one of the lo-siRF-prioritized loci. Here, the top interaction is between rs12618871 (chromosome 2; *CCDC141* locus) and rs9267356 (chromosome 6; *MICB* locus).

These differences in the top interactions across the different LVMi responses further highlight the instability of the regression-based pairwise interaction scan. This instability poses issues in interpreting results, as it can be unclear which results to trust. We suspect that at least part of the instability is a result of the low signal-to-noise ratio in this problem and highlights the need for more robust and stability-driven methods, such as lo-siRF, for detecting epistasis in low signal-to-noise regimes.

Figure 8.1: **Pair plot of the LVMi interaction effect p-values from an exhaustive regression-based SNVxSNV interaction scan using different transformations of the LVMi phenotype.** The transformations of interest include the rank-based inverse normal-transformed LVMi and the binarized LVMi using three different binarization thresholds (15%, 20%, 25%). The x- and y-axes show the interaction effect p-values after a -log10 transformation. Each point represents an SNV-SNV pair. We observe that the interaction effect p-values are generally unstable and depend on the choice of transformation of the phenotype (or response).

Figure 8.2: **List of SNV-SNV interactions from an exhaustive regression-based SNVxSNV interaction scan using different transformations of the LVMi phenotype.** The transformations of interest include the rank-based inverse normal-transformed LVMi and the binarized LVMi using three different binarization thresholds (15%, 20%, 25%). The table sorts the SNV-SNV interactions based on the mean p-value across the different transformations of the LVMi phenotype. To sort the SNV-SNV interactions based upon their p-value from a particular transformation, please use the toggles next to the column names. Only interactions that yielded a p-value < 0.01 for at least one transformation of the LVMi phenotype are shown for brevity.

##### MAPIT

Given the instabilities observed in the exhaustive regression-based interaction search, we applied MAPIT (Crawford et al. 2017) to our dataset as an additional comparison method. At a high-level, MAPIT aims to identify variants with non-zero marginal epistatic effects, defined as the total pairwise interaction effect between the given variant and all other variants in the data.

Similar to before, we used the GWAS-filtered SNVs with minor allele frequency > 0.05 as input to MAPIT. For the response, we used the rank-based inverse normal-transformed LVMi only since the MAPIT extension to binary traits has not yet been implemented. Details regarding the methodology can be found in Q. Wang et al. (2023).

- Note: we also tried applying MAPIT to the full set of 1405 GWAS-filtered SNVs (without the additional minor allele frequency filter). However, the resulting top SNVs were predominantly those with the smallest minor allele frequencies, and we omit these results.

The top 5 SNVs from MAPIT with the smallest p-values were located in the *MAT2B;LINC02143*, *MICB*, *BRMS1L;LINC00609*, and *CDKN1A* loci (Figure 8.3). Interestingly, the highest-ranked SNVs from the *CCDC141*, *TTN*, and *IGF1R* loci were ranked 109 (p = 0.028), 75 (p = 0.003), and 87 (p = 0.011), respectively (compared to p < 8E-5 for the top 5 SNVs), according to MAPIT.

As is, the results from MAPIT and lo-siRF are not directly comparable since they are operating at different biological scales (i.e., MAPIT aims to identify epistatic variants while lo-siRF aims to identify epistatic loci). Variant-level approaches are known to be under-powered in traits where the effect of each variant is very small and/or noisy while group-level (or locus-level) approaches aggregate these signals across a collection of variants in hope of detecting more stable and biologically-relevant loci. In addition to this difference, there are a number of other potential reasons why MAPIT and lo-siRF may yield different, but non-contradictory results. For example, MAPIT and lo-siRF assume different underlying model assumptions and may capture different forms of interactions. In short, MAPIT encodes interactions as pairwise multiplicative effects (more typical in the statistical epistasis literature) whereas lo-siRF takes a more machine learning approach, encoding pairwise and higher-order interactions through decision paths (or boolean thresholding operations) in a decision tree. Other potential differences may stem from the different handling of variants with minor allele frequencies below 0.05 or the slightly different phenotypes used (i.e., lo-siRF used the binarized LVMi phenotype while MAPIT unfortunately requires a continuous phenotype and thus used the rank-based inverse normal-transformed LVMi).

Figure 8.3: **Top epistatic variants from MAPIT, ranked by p-value.** We show only the epistatic variants with p-value < 0.05. rsID = rsID of the variant. Chr = Chromosome. Pos = Position on hg19. Lo-siRF Locus = assigned locus of the variant in the lo-siRF analysis. P-value = p-value from MAPIT.

##### MAPIT+GSEA

Since the results of MAPIT and lo-siRF are on different biological scales (i.e., SNVs versus loci), we also conducted a locus-level analysis using MAPIT (Crawford et al. 2017) in conjunction with gene set enrichment analysis (GSEA) (Subramanian et al. 2005; Mootha et al. 2003). In this pipeline, we took the union of the top 10,000 GWAS hits from PLINK and BOLT-LMM, ran MAPIT on these SNVs to obtain a ranked list of SNVs according to their MAPIT p-values, and then ran GSEA on this ranked list to identify loci that are enriched at the top of this ranked list. The results of this MAPIT+GSEA analysis are provided below. We highlight three notable findings from this analysis: When ranking the loci by their normalized enrichment score,

- The top-ranked locus (by MAPIT+GSEA) was the intergenic region between *IGFBP3* and *LOC730338* (denoted as *IGFBP3;LOC730338*). *IGFBP3* encodes an insulin-like growth factor (IGF) binding protein, which is known to interact closely with *IGF1R* (Baxter 2023).
- The three interacting loci, prioritized by lo-siRF, were all ranked within the top 30 by the MAPIT+GSEA analysis (*LOC157273;TNKS*: rank 2; *IGF1R*: rank 7; *TTN*: rank 23; *CCDC141*: rank 28 out of 863 total loci). Their corresponding nominal p-value, FDR q-value, and FWER p-value were 0 (or < 1e-3 given that we used 1000 permutations). We note however that the magnitudes of these p-values are not directly comparable to those from lo-siRF given the different modeling assumptions.
- Aside from *IGFBP3;LOC730338*, *LOC157273;TNKS*, *IGF1R*, and *ADAMTS16*, other loci ranked in the top 10 by MAPIT+GSEA do not appear to be closely related to cardiac hypertrophy or cardiovascular diseases in the current literature. Moreover, except for *IGFBP3;LOC730338*, *LOC157273;TNKS*, *IGF1R*, *ADAMTS16*, and *FBXL17*, the remaining 5 loci in the top 10 were not among the top 1000 GWAS-filtered SNVs used in the original lo-siRF analysis. In other words, half of the top 10 loci identified by MAPIT+GSEA did not show strong marginal associations with LVMi. There is no expectation that variants involved in epistasis also have large marginal effects, so the lack of marginal associations alone is not particularly concerning. However, combined with the observation that the primary biological functions of these loci are not directly related to the heart, we are uncertain whether these epistatic findings would be validated in wet lab experiments.

Overall, in comparing lo-siRF with MAPIT+GSEA, we are encouraged to see that both methods identified loci like *LOC157273;TNKS*, *IGF1R*, *TTN*, and *CCDC141*. On the other hand, MAPIT+GSEA also identified loci deprioritized by lo-siRF, which turn out to lack biological relevance to cardiac diseases. These discrepancies may arise from the fundamental differences between the computational frameworks of lo-siRF and MAPIT+GSEA, which make direct comparisons challenging. First, MAPIT (or MAPIT+GSEA) outputs single SNVs or loci with marginal epistatic effects while lo-siRF directly identifies specific interactions between genetic loci. Secondly, lo-siRF and MAPIT are based on different modeling assumptions, so their definitions of epistasis are not directly comparable. Lastly, since performing GSEA after MAPIT has not been extensively studied, it is unclear whether the observed strengths and weaknesses of MAPIT+GSEA arise from MAPIT, GSEA, or their combination. It also remains uncertain whether using alternative SNV-to-locus grouping methods (in place of GSEA) would improve upon the epistatic loci rankings.

Figure 8.4: **Top epistatic loci from MAPIT+GSEA, ranked by normalized enrichment score.** We show only the epistatic loci with nominal p-value < 0.05.

#### Marginal Gene-based Methods

In this section, we compare the lo-siRF-prioritized loci to those loci prioritized by other existing marginal set-based methods, such as MAGMA (Leeuw et al. 2015) (Figures 8.5-8.6) and SKAT-O (Lee et al. 2012) (Figure 8.7). Details regarding the methodology can be found in Q. Wang et al. (2023).

Notably, we found that while lo-siRF prioritized *IGF1R* as one of its top recommended loci, SKAT-O and MAGMA did not rank *IGF1R* very highly. According to SKAT-O, *IGF1R* yielded a p-value of 1.0E-5 and was the 45th ranked locus, and using MAGMA, *IGF1R* yielded a p-value of 0.002 and was the 146th ranked locus. Given our extensive validation efforts demonstrating the importance of *IGF1R* for left ventricular hypertrophy, we view this as an interesting example where lo-siRF is able to identify important loci that are not necessarily prioritized by other methods, even for marginal effects.

Figure 8.5: Manhattan plot of the gene-based test as computed by MAGMA. The genome-wide significance level is shown by the red dotted line.

Figure 8.6: **Top genes from MAGMA, ranked by p-value.** We show only the top 200 genes. Locus = Gene symbol. Chr = Chromosome. Start = Start position on hg19. Stop = Stop position on hg19. Z = Z-statistic from MAGMA. P-value = p-value from MAGMA.

Figure 8.7: **Top genes from SKAT-O, ranked by p-value.** Locus = Name of locus from lo-siRF analysis. Chr = Chromosome. P-value = p-value from SKAT-O.

### 9 Lo-siRF Permutation Test Simulations

In this section, we conduct several simulations to investigate the performance and demonstrate the validity of the permutation p-values from lo-siRF.

#### P-value Calibration Simulations

**Calibration of p-values under null model**: In the first set of simulations, we aimed to assess whether the permutation p-values are calibrated under the null model where no epistasis is present. To this end, we simulated binary responses \(y\) from a null, purely-noise model, where \(y \sim Bernoulli(1/2)\). Under this null simulation setup, we generated covariates \(X\) from three different data-generating processes:

1. Independent normal: \(X\_{ij} \stackrel{iid}{\sim} N(0, 1)\) for \(i = 1, \ldots, n\) and \(j = 1, \ldots, p\).
2. Independent SNPs: \(X\_{ij}\) iid from \(\{0, 1, 2\}\) with uniform probabilities for \(i = 1, \ldots, n\) and \(j = 1, \ldots, p\).
3. Real-world SNPs: The \(n \times p\) matrix \(X\) is taken to be the GWAS-filtered SNV matrix used in lo-siRF. In this case, we we randomly subsampled the features and the columns to meet the desired dimensions of \(n\) and \(p\).

Under each of these three settings, we ran the simulated data through a simplified lo-siRF pipeline, where we:

1. Split the data into two partition (2/3 training and 1/3 validation).
2. Fit siRF on the training data to obtain a list of candidate interactions.
3. For each of the interactions from step 2, evaluate the local stability importance scores and compute the associated permutation p-value using the validation data.

In simulations (A) and (B), we searched for interactions at the level of individual features (or SNVs), rather than a group of features (or genes). This was done to simplify the simulation setup and interpretation of results, given that the features were simulated independently. For simulation setup (C), we leveraged the same SNV-to-gene mapping used previously in lo-siRF and searched for interactions at the gene level.

We also note that the simplified lo-siRF pipeline omits the interaction filtering step based upon their siRF stability scores. This was done because the interaction filtering step excludes almost all non-relevant candidate interactions under the simulation setup. In particular, under the null simulation model, the interaction filtering (using the recommended thresholds from the original siRF paper (Kumbier et al. 2023)) removed all but 2 interactions across 200 simulated data replicates. Given our goal of assessing whether the permutation p-values are calibrated, we will focus our evaluation on the permutation p-values from all candidate interactions in this setting regardless of whether the interactions passed the stability filtering.

**Results**: In Figure 9.1 below, we plot the proportion of rejected null hypotheses across different significance levels \(\alpha\) under the aforementioned null simulation setup. We found that the observed type I error of our permutation test closely aligns with the significance level \(\alpha\), thereby confirming that the p-values are calibrated. Moreover, these results hold across different sample sizes, number of features, and all three data-generating processes for X.

Figure 9.1: Proportion of rejected null hypotheses across varying significance levels under the null simulation response model are represented by the solid black line. Dotted line represents a perfectly calibrated hypothesis test (i.e., the y = x line). Each row corresponds to a different data-generating process for X, and each column corresponds to a different choice of number of samples. Results are aggregated across 200 simulation replicates.

#### Marginal and Interaction Effect Simulations

**Marginal and Interaction Effect Simulations**: Moving beyond this basic calibration investigation and the null, purely-noise model, we were more interested in studying the broader performance of lo-siRF and the permutation test. To this end, we conducted two additional simulations using boolean-type rules. We note that the boolean-type rules were chosen as they have been extensively studied in previous RF-based work and are thought to closely resemble the switch-like and stereospecific nature of interactions among biomolecules (Nelson, Lehninger, and Cox 2008; Kumbier et al. 2023; Behr et al. 2020). Specifically, we first simulated the \(X\) data from independent SNVs, where \(X\_{ij}\) is simulated iid from \(\{0, 1, 2\}\) with uniform probabilities. Next, we simulated binary responses \(y \mid X \sim Bernoulli(p(X))\) via:

4. Marginal effect model: \(p(X) = 0.4 \cdot \mathbf{1}\{X\_1 > 0\} + 0.4 \cdot \mathbf{1}\{X\_2 > 0\}\)

- In this model, there are no true interactions.

5. Interaction effect model: \(p(X) = 0.4 \cdot \mathbf{1}\{X\_1 > 0\} \mathbf{1}\{X\_2 > 0\} + 0.4 \cdot \mathbf{1}\{X\_3 > 0\} \mathbf{1}\{X\_4 > 0\}\)

- In this model, the true interactions are \(X\_1:X\_2\) and \(X\_3:X\_4\).

We set the number of features to 100 and varied the number of samples across n = 1000, 1500, 2000. For each simulation model and choice of sample size, we ran 200 simulation replicates.

**Results**: The simulation results are summarized in Table 9.2 below. In what follows, we define “partial interaction” as a candidate interaction that involves at least one true signal feature from an interaction that appeared in the simulation model — specifically, at least one of \(X\_1\) or \(X\_2\) in the marginal effect simulation (D), or at least one of \(X\_1\), \(X\_2\), \(X\_3\), or \(X\_4\) in the interaction effect simulation (E). We define “true interaction” as a candidate interaction that involves both \(X\_1\) and \(X\_2\) or both \(X\_3\) and \(X\_4\) in the interaction effect simulation.

The main takeaways from these simulations are:

1. Under both the marginal effect and interaction effect scenarios, the candidate interactions from siRF almost always include at least one true signal feature (and equivalently, is a partial interaction).

   - Specifically, under both the marginal effect (D) and interaction effect (E) scenarios, over 99% of the candidate interactions identified by siRF were found to be “partial interactions” (the 5th column in Table 9.2). Moreover, under the interaction effect (E) simulation, over 38% of the candidate interactions identified by siRF were “true interactions” (the 4th column in Table 9.2).
2. The interaction filtering step within siRF, where we filter out candidate interactions that do not meet the siRF stability thresholds, is both necessary and effective at distinguishing between interactions and marginal effects.

   - First, the siRF interaction filtering is necessary to differentiate between interactions and marginal effects since the permutation test itself is not designed to distinguish between “true interactions” and “partial interactions.”

     - Because the permutation test within lo-siRF formally assesses whether the local stability importance of an interaction differs between the high and low LVMi groups, any candidate interaction involving at least one true signal feature (i.e., any partial interaction) is expected to show a difference between the two groups and result in a significant p-value. Our results support this expectation. In both the interaction and marginal simulation models, an overwhelming majority (>99%) of the permutation tests led to a significant p-value (p < 0.05) (the last column in Table 9.2). This is expected since each of these tested interactions was a partial interaction and involved at least one true signal feature (the 8th column in Table 9.2).
     - We thus relied on the siRF interaction filtering step to distinguish between marginal and true interaction effects. This siRF interaction filtering step is detailed in the Methods section *Lo-siRF step 4.3: Permutation test for difference in local stability importance scores* in Q. Wang et al. (2023). Specifically, the feature selection dependence threshold (discussed in greater detail in the original siRF paper (Kumbier et al. 2023)) was originally designed to differentiate between additive (e.g., marginal) and non-additive (e.g., interaction) effects. This metric showed strong performance in empirical simulations from the original siRF paper.
   - As in the original siRF paper, the siRF interaction filtering demonstrated similarly strong effectiveness for distinguishing between true interactions and true marginal effects under our simulation setup.

     - In particular, under the interaction simulation model, the siRF interaction filtering step reduced the number of interactions from over 3000 candidates (the 3rd column in Table 9.2), of which \(\sim\) 40% were true interactions (4th column), to ~1300 candidates (6th column). Of these remaining \(\sim\) 1300 candidates, 87%, 99%, and 100% were true interactions in the n = 1000, 1500, and 2000 simulations, respectively (the 7th column).
     - Furthermore, under the marginal simulation model, the siRF interaction filtering step reduced the number of interactions from over 4400 candidates (the 3rd column in Table 9.2) to fewer than 100 candidates (4th column). That is, less than 2% of the candidate interactions remain after the siRF interaction filtering step in the marginal effect simulation setting.
   - Thus, even though the permutation test rejects most tests involving partial interactions, in lo-siRF, the permutation test occurs after the siRF interaction filtering, which we have shown to effectively eliminate the vast majority of non-signal or partial interactions.

Figure 9.2: Summary of marginal and interaction effect simulations. # Candidate Interactions from siRF: The total number of interactions identified by siRF before applying the stability filtering step. Prop. (Num.) of True Interactions in Candidate Set: The proportion (or number) of true interactions out of the total number of candidate interactions from siRF. Prop. (Num.) of Partial Interactions in Candidate Set: The proportion (or number) of partial interactions out of the total number of candidate interactions from siRF. Prop. (Num.) of Stable Interactions in Candidate Set: The proportion (or number) of interactions that passed the stability filter step out of the total number of candidate interactions from siRF. Prop. (Num.) of True Interactions in Stable Set: The proportion (or number) of true interactions out of the total number of interactions that passed the stability filter. Prop. (Num.) of Partial Interactions in Stable Set: The proportion (or number) of partial interactions out of the total number of interactions that passed the stability filter. # Interactions Rejected (p < 0.05): The number of stable (or tested) interactions with a permutation test p-value below 0.05.

### 10 Final Remarks

We reiterate that lo-siRF was originally developed as a first-stage recommendation (or hypothesis generation) tool within a broader pipeline in order to prioritize genetic loci and interactions between loci for downstream analyses and experimental validation. Importantly, the output of lo-siRF is a prioritization/ranking order, not a proper statistical test of significance. We rely on follow-up investigations and in this study rigorous gene-silencing experiments in order to validate the prioritized genetic loci and interaction between loci. With that, many of the modeling decisions and human judgment calls were made with this original use case in mind. For other use cases or problems, it is highly likely that different choices may be better suited. We also acknowledge that though many stability checks were conducted to help ensure that our conclusions are stable across different reasonable data and/or modeling perturbations, these stability checks are inevitably constrained by computational limitations, and additional stability analyses can be conducted. We hope that by documenting our modeling choices, stability analyses, and providing insights into our motivations/reasons, this may help to both clarify the limitations of our current work and to facilitate the trustworthy (veridical) use of lo-siRF (and variants thereof) in other scientific problems.

Kumbier, Karl, Sumanta Basu, Erwin Frise, James B Brown, Susan Celniker, and Bin Yu. 2023. “Signed Iterative Random Forests to Identify Enhancer-Associated Transcription Factor Binding.” *arXiv Preprint arXiv:1810.07287*.

Lee, Seunggeun, Mary J Emond, Michael J Bamshad, Kathleen C Barnes, Mark J Rieder, Deborah A Nickerson, David C Christiani, Mark M Wurfel, and Xihong Lin. 2012. “Optimal Unified Approach for Rare-Variant Association Testing with Application to Small-Sample Case-Control Whole-Exome Sequencing Studies.” *The American Journal of Human Genetics* 91 (2): 224–37.

Leeuw, Christiaan A de, Joris M Mooij, Tom Heskes, and Danielle Posthuma. 2015. “MAGMA: Generalized Gene-Set Analysis of GWAS Data.” *PLoS Computational Biology* 11 (4): e1004219.

Loh, Po-Ru, George Tucker, Brendan K Bulik-Sullivan, Bjarni J Vilhjalmsson, Hilary K Finucane, Rany M Salem, Daniel I Chasman, et al. 2015. “Efficient Bayesian Mixed-Model Analysis Increases Association Power in Large Cohorts.” *Nature Genetics* 47 (3): 284–90.

Mbatchou, Joelle, Leland Barnard, Joshua Backman, Anthony Marcketta, Jack A Kosmicki, Andrey Ziyatdinov, Christian Benner, et al. 2021. “Computationally Efficient Whole-Genome Regression for Quantitative and Binary Traits.” *Nature Genetics* 53 (7): 1097–1103.

Meyer, Hannah V, Timothy JW Dawes, Marta Serrani, Wenjia Bai, Paweł Tokarczuk, Jiashen Cai, Antonio de Marvao, et al. 2020. “Genetic and Functional Insights into the Fractal Structure of the Heart.” *Nature* 584 (7822): 589–94.

Mizukoshi, Kei, Masaaki Takeuchi, Yasufumi Nagata, Karima Addetia, Roberto M Lang, Yoshihiro J Akashi, and Yutaka Otsuji. 2016. “Normal Values of Left Ventricular Mass Index Assessed by Transthoracic Three-Dimensional Echocardiography.” *Journal of the American Society of Echocardiography* 29 (1): 51–61.

Mootha, Vamsi K, Cecilia M Lindgren, Karl-Fredrik Eriksson, Aravind Subramanian, Smita Sihag, Joseph Lehar, Pere Puigserver, et al. 2003. “PGC-1\(\alpha\)-Responsive Genes Involved in Oxidative Phosphorylation Are Coordinately Downregulated in Human Diabetes.” *Nature Genetics* 34 (3): 267–73.

Nelson, David L, Albert L Lehninger, and Michael M Cox. 2008. *Lehninger Principles of Biochemistry*. Macmillan.

Niel, Clément, Christine Sinoquet, Christian Dina, and Ghislain Rocheleau. 2015. “A Survey about Methods Dedicated to Epistasis Detection.” *Frontiers in Genetics* 6: 285.

Pirruccello, James P, Alexander Bick, Minxian Wang, Mark Chaffin, Samuel Friedman, Jie Yao, Xiuqing Guo, et al. 2020. “Analysis of Cardiac Magnetic Resonance Imaging in 36,000 Individuals Yields Genetic Insights into Dilated Cardiomyopathy.” *Nature Communications* 11 (1): 2254.

Purcell, Shaun, Benjamin Neale, Kathe Todd-Brown, Lori Thomas, Manuel AR Ferreira, David Bender, Julian Maller, et al. 2007. “PLINK: A Tool Set for Whole-Genome Association and Population-Based Linkage Analyses.” *The American Journal of Human Genetics* 81 (3): 559–75.

Schaid, Daniel J, Wenan Chen, and Nicholas B Larson. 2018. “From Genome-Wide Associations to Candidate Causal Variants by Statistical Fine-Mapping.” *Nature Reviews Genetics* 19 (8): 491–504.

Subramanian, Aravind, Pablo Tamayo, Vamsi K Mootha, Sayan Mukherjee, Benjamin L Ebert, Michael A Gillette, Amanda Paulovich, et al. 2005. “Gene Set Enrichment Analysis: A Knowledge-Based Approach for Interpreting Genome-Wide Expression Profiles.” *Proceedings of the National Academy of Sciences* 102 (43): 15545–50.

Toloşi, Laura, and Thomas Lengauer. 2011. “Classification with Correlated Features: Unreliability of Feature Ranking and Solutions.” *Bioinformatics* 27 (14): 1986–94.

Visscher, Peter M, Naomi R Wray, Qian Zhang, Pamela Sklar, Mark I McCarthy, Matthew A Brown, and Jian Yang. 2017. “10 Years of GWAS Discovery: Biology, Function, and Translation.” *The American Journal of Human Genetics* 101 (1): 5–22.

Walker, Angelica M, Ashley Cliff, Jonathon Romero, Manesh B Shah, Piet Jones, Joao Gabriel Felipe Machado Gazolla, Daniel A Jacobson, and David Kainer. 2022. “Evaluating the Performance of Random Forest and Iterative Random Forest Based Methods When Applied to Gene Expression Data.” *Computational and Structural Biotechnology Journal* 20: 3372–86.

Wang, Kai, Mingyao Li, and Hakon Hakonarson. 2010. “ANNOVAR: Functional Annotation of Genetic Variants from High-Throughput Sequencing Data.” *Nucleic Acids Research* 38 (16): e164–64.

Wang, Qianru, Tiffany M Tang, Nathan Youlton, Chad S Weldy, Ana M Kenney, Omer Ronen, J Weston Hughes, et al. 2023. “Epistasis Regulates Genetic Control of Cardiac Hypertrophy.”

Watanabe, Kyoko, Erdogan Taskesen, Arjen Van Bochoven, and Danielle Posthuma. 2017. “Functional Mapping and Annotation of Genetic Associations with FUMA.” *Nature Communications* 8 (1): 1826.

Yildiz, Mehmet, Ahmet Afşin Oktay, Merrill H Stewart, Richard V Milani, Hector O Ventura, and Carl J Lavie. 2020. “Left Ventricular Hypertrophy and Hypertension.” *Progress in Cardiovascular Diseases* 63 (1): 10–21.

Yu, Bin. 2013. “Stability.”

Yu, Bin, and Karl Kumbier. 2020. “Veridical Data Science.” *Proceedings of the National Academy of Sciences* 117 (8): 3920–29. https://doi.org/10.1073/pnas.1901326117.

Code 

- Show All Code
- Hide All Code
- Download Rmd
