## Supplementary webpage 2 for "Deciphering epistatic genetic regulation of cardiac hypertrophy": Supplementary_webpage_2.html

Examining the effect of varying gene-silencing efficiencies on epistasis testing: A simulation study


### Examining the effect of varying gene-silencing efficiencies on epistasis testing: A simulation study

###### March 18, 2024

- Simulation Experiment Recipe
- CCDC141-IGF1R, Diseased Simulation
- CCDC141-IGF1R, Healthy Simulation
- CCDC141-TTN, Diseased Simulation
- CCDC141-TTN, Healthy Simulation
- Efficiency = 1 Simulation

### Simulation Experiment Recipe

#### Objectives

In Wang et al. (2023), high-throughput single-cell gene-silencing experiments were performed to assess the presence of epistasis (i.e,. non-additive interaction effects) in the genetic regulation of cardiac structure. More specifically, these single-cell experiments were designed to assess whether the genes pairs *CCDC141* and *TTN* as well as *CCDC141* and *IGF1R* interact non-additively to impact the cell diameter of human induced pluriopotent stem cell-derived cardiomyocytes. Mathematically, this boils down to studying the following regression problem: for a gene pair, say gene 1 and gene 2, consider

\[y\_i = \beta\_0 + \beta\_1 x\_{i1} + \beta\_2 x\_{i2} + \beta\_{12} x\_{i1} x\_{i2} + \epsilon\_i,\]

where \(y\_i\) is the diameter (\(\mu m\)) of cell \(i\), \(x\_{ij}\) is an indicator of whether cell \(i\) belonged to an experimental batch where gene \(j\) was silenced, and \(\epsilon\_i\) is the random error or noise term for cell \(i\). Under this model, the quantity of interest is \(\beta\_{12}\), or the coefficient corresponding to the non-additive interaction effect between genes 1 and 2, and we would like to assess the null hypothesis that \(\beta\_{12} = 0\) (i.e., no epistatic effect) versus the alternative hypothesis that \(\beta\_{12} \neq 0\) (i.e., a non-zero epistatic effect).

One possible complication in the statistical analysis of gene-silencing experiments, however, is that the gene-silencing efficiency may not be 100% and may vary across different experimental batches of cells. In this simulation study, we aim to assess whether variation in silencing efficiencies lead to spurious epistasis signals.

Overall, we found unsurprisingly that the signal-to-noise ratio plays a role in understanding the effect of varying silencing efficiencies. But notably, in the very low signal-to-noise regimes that we expect to be realistic in practice, we found that the differences in our observed gene-silencing efficiencies do not typically lead to high false positive rates. Furthermore, when the strength of the interaction effect is small but not 0, the resulting quantile regression p-values are frequently not significant. We realize however that this simulation models the gene-silencing efficiencies in a particular way, and model misspecification of not only the silencing efficiencies but the model form itself is rightfully a concern. We thus simply view this simulation study as a helpful starting point and acknowledge that additional simulations can be done to explore these avenues.

In what follows, we provide additional details regarding the simulation setup (see tabs above) as well as the results of the simulation study (see section tabs on the left side panel).

#### Data Generation

##### CCDC141-IGF1R, Diseased

To simulate data from batches with different gene-silencing efficiencies, we generated data via the following model:

\[y\_i = \beta\_0 + (\beta\_1 x\_{i1} + \beta\_2 x\_{i2} + \beta\_{12} x\_{i1} x\_{i2}) \cdot z\_i + \epsilon\_i,\]

where \(y\_i\) is the diameter (\(\mu m\)) of cell \(i\), \(x\_{ij}\) is an indicator of whether cell \(i\) belonged to an experimental batch where gene \(j\) was silenced, \(z\_i\) is an indicator of whether the gene(s) were successfully silenced in cell \(i\), generated via \(z\_i \mid x\_{i1} = j, x\_{i2} = k \stackrel{ind.}{\sim} Bernoulli(\pi\_{jk})\), and \(\epsilon\_i \stackrel{iid}{\sim} N(0, \sigma^2)\) is the random error or noise term for cell \(i\).

We simulated \(n = 500\) cells for each of the gene-silencing conditions, that is,

- 500 scrambled control cells (i.e., \(x\_{i1} = x\_{i2} = 0\)),
- 500 cells wherein gene 1 was silenced (i.e., \(x\_{i1} = 1\) and \(x\_{i2} = 0\)),
- 500 cells wherein gene 2 was silenced (i.e., \(x\_{i1} = 0\) and \(x\_{i2} = 1\)), and
- 500 cells wherein gene 1 and gene 2 were silenced (i.e., \(x\_{i1} = x\_{i2} = 1\)).

Unless otherwise specified, simulations were run across 100 replicates, and the following simulation parameter values were used:

- \(\beta\_0 = 0\)
- \(\beta\_1 = -1\)
- \(\beta\_2 = -1\)
- \(\beta\_{12} = 0\)
- \(\sigma^2 = 1\)

We then ran simulations that varied across the interaction effect strength \(\beta\_{12} \in \{-1, -0.5, -0.2, -0.1, 0\}\) and the noise level \(\sigma \in \{0.1, 0.2, 0.5, 1, 1.5, 2\}\).

So that we may best connect to the observed experimental data in Wang et al. (2023), we took the observed gene-silencing efficiencies from the experiments and used these values for the simulated gene-silencing efficiencies \(\{\pi\_{01}, \pi\_{10}, \pi\_{11}\}\). Since there were four scenarios of interest (i.e., the *CCDC141*-*TTN* interaction in the healthy and diseased (MYH7-R403Q) cell lines and the *CCDC141*-*IGF1R* interaction in the healthy and diseased (MYH7-R403Q) cell lines), we repeated the simulation study under four different choices of \(\{\pi\_{01}, \pi\_{10}, \pi\_{11}\}\).

For this particular data-generating process, we set the simulated gene-silencing efficiencies to be those from the *CCDC141*-*IGF1R* experiment in the healthy cell line so that

- \(\pi\_{01} = 0.92\)
- \(\pi\_{10} = 0.83\)
- \(\pi\_{11} = 0.695\)

*Remark*: We note that the above model is a simplistic interpretation of how gene-silencing efficiencies may effect the response of interest and that there are other ways to incorporate gene-silencing efficiencies into the model that may be more realistic. We view this simulation study as a helpful *starting* point for understanding the effect of varying gene-silencing efficiencies on epistasis modeling.

**Function**

```
## function(n, beta0, beta1, beta2, beta12,
##                                eff1 = 1, eff2 = 1, int_eff = 1, 
##                                err_fun = rnorm, ...) {
##   eff1_vec <- rbinom(n, 1, eff1)
##   eff2_vec <- rbinom(n, 1, eff2)
##   int_eff_vec <- rbinom(n, 1, int_eff)
##   
##   y0 <- beta0 + err_fun(n, ...)
##   y1 <- beta0 + beta1 * eff1_vec + err_fun(n, ...)
##   y2 <- beta0 + beta2 * eff2_vec + err_fun(n, ...)
##   y12 <- beta0 + (beta1 + beta2 + beta12) * int_eff_vec + err_fun(n, ...)
##   
##   x <- matrix(0, nrow = 4 * n, ncol = 2)
##   x[(n + 1):(2 * n), 1] <- 1
##   x[(2 * n + 1):(3 * n), 2] <- 1
##   x[(3 * n + 1):(4 * n), 1:2] <- 1
##   y <- c(y0, y1, y2, y12)
##   
##   out <- list(
##     x = x,
##     y = y,
##     eff1_vec = eff1_vec,
##     eff2_vec = eff2_vec,
##     int_eff_vec = int_eff_vec
##   )
## }
```

**Input Parameters**

```
## $n
## [1] 500
## 
## $beta0
## [1] 0
## 
## $beta1
## [1] -1
## 
## $beta2
## [1] -1
## 
## $beta12
## [1] 0
## 
## $eff1
## [1] 0.92
## 
## $eff2
## [1] 0.83
## 
## $int_eff
## [1] 0.695
## 
## $err_fun
## function (n, mean = 0, sd = 1) 
## .Call(C_rnorm, n, mean, sd)
## <bytecode: 0x7feab78c3da0>
## <environment: namespace:stats>
## 
## $sd
## [1] 1
```

##### CCDC141-IGF1R, Healthy

In this data-generating process, we simulated data as described above but set the simulated gene-silencing efficiencies to be the observed efficiencies from the *CCDC141*-*IGF1R* experiment in the diseased cell line:

- \(\pi\_{01} = 0.66\)
- \(\pi\_{10} = 0.9\)
- \(\pi\_{11} = 0.725\)

**Function**

```
## function(n, beta0, beta1, beta2, beta12,
##                                eff1 = 1, eff2 = 1, int_eff = 1, 
##                                err_fun = rnorm, ...) {
##   eff1_vec <- rbinom(n, 1, eff1)
##   eff2_vec <- rbinom(n, 1, eff2)
##   int_eff_vec <- rbinom(n, 1, int_eff)
##   
##   y0 <- beta0 + err_fun(n, ...)
##   y1 <- beta0 + beta1 * eff1_vec + err_fun(n, ...)
##   y2 <- beta0 + beta2 * eff2_vec + err_fun(n, ...)
##   y12 <- beta0 + (beta1 + beta2 + beta12) * int_eff_vec + err_fun(n, ...)
##   
##   x <- matrix(0, nrow = 4 * n, ncol = 2)
##   x[(n + 1):(2 * n), 1] <- 1
##   x[(2 * n + 1):(3 * n), 2] <- 1
##   x[(3 * n + 1):(4 * n), 1:2] <- 1
##   y <- c(y0, y1, y2, y12)
##   
##   out <- list(
##     x = x,
##     y = y,
##     eff1_vec = eff1_vec,
##     eff2_vec = eff2_vec,
##     int_eff_vec = int_eff_vec
##   )
## }
```

**Input Parameters**

```
## $n
## [1] 500
## 
## $beta0
## [1] 0
## 
## $beta1
## [1] -1
## 
## $beta2
## [1] -1
## 
## $beta12
## [1] 0
## 
## $eff1
## [1] 0.66
## 
## $eff2
## [1] 0.9
## 
## $int_eff
## [1] 0.725
## 
## $err_fun
## function (n, mean = 0, sd = 1) 
## .Call(C_rnorm, n, mean, sd)
## <bytecode: 0x7feab72fd800>
## <environment: namespace:stats>
## 
## $sd
## [1] 1
```

##### CCDC141-TTN, Diseased

In this data-generating process, we simulated data as described above but set the simulated gene-silencing efficiencies to be the observed efficiencies from the *CCDC141*-*TTN* experiment in the diseased cell line:

- \(\pi\_{01} = 0.92\)
- \(\pi\_{10} = 0.99\)
- \(\pi\_{11} = 0.92\)

**Function**

```
## function(n, beta0, beta1, beta2, beta12,
##                                eff1 = 1, eff2 = 1, int_eff = 1, 
##                                err_fun = rnorm, ...) {
##   eff1_vec <- rbinom(n, 1, eff1)
##   eff2_vec <- rbinom(n, 1, eff2)
##   int_eff_vec <- rbinom(n, 1, int_eff)
##   
##   y0 <- beta0 + err_fun(n, ...)
##   y1 <- beta0 + beta1 * eff1_vec + err_fun(n, ...)
##   y2 <- beta0 + beta2 * eff2_vec + err_fun(n, ...)
##   y12 <- beta0 + (beta1 + beta2 + beta12) * int_eff_vec + err_fun(n, ...)
##   
##   x <- matrix(0, nrow = 4 * n, ncol = 2)
##   x[(n + 1):(2 * n), 1] <- 1
##   x[(2 * n + 1):(3 * n), 2] <- 1
##   x[(3 * n + 1):(4 * n), 1:2] <- 1
##   y <- c(y0, y1, y2, y12)
##   
##   out <- list(
##     x = x,
##     y = y,
##     eff1_vec = eff1_vec,
##     eff2_vec = eff2_vec,
##     int_eff_vec = int_eff_vec
##   )
## }
```

**Input Parameters**

```
## $n
## [1] 500
## 
## $beta0
## [1] 0
## 
## $beta1
## [1] -1
## 
## $beta2
## [1] -1
## 
## $beta12
## [1] 0
## 
## $eff1
## [1] 0.92
## 
## $eff2
## [1] 0.99
## 
## $int_eff
## [1] 0.92
## 
## $err_fun
## function (n, mean = 0, sd = 1) 
## .Call(C_rnorm, n, mean, sd)
## <bytecode: 0x7feab75fcfd8>
## <environment: namespace:stats>
## 
## $sd
## [1] 1
```

##### CCDC141-TTN, Healthy

In this data-generating process, we simulated data as described above but set the simulated gene-silencing efficiencies to be the observed efficiencies from the *CCDC141*-*TTN* experiment in the healthy cell line:

- \(\pi\_{01} = 0.66\)
- \(\pi\_{10} = 0.8\)
- \(\pi\_{11} = 0.93\)

**Function**

```
## function(n, beta0, beta1, beta2, beta12,
##                                eff1 = 1, eff2 = 1, int_eff = 1, 
##                                err_fun = rnorm, ...) {
##   eff1_vec <- rbinom(n, 1, eff1)
##   eff2_vec <- rbinom(n, 1, eff2)
##   int_eff_vec <- rbinom(n, 1, int_eff)
##   
##   y0 <- beta0 + err_fun(n, ...)
##   y1 <- beta0 + beta1 * eff1_vec + err_fun(n, ...)
##   y2 <- beta0 + beta2 * eff2_vec + err_fun(n, ...)
##   y12 <- beta0 + (beta1 + beta2 + beta12) * int_eff_vec + err_fun(n, ...)
##   
##   x <- matrix(0, nrow = 4 * n, ncol = 2)
##   x[(n + 1):(2 * n), 1] <- 1
##   x[(2 * n + 1):(3 * n), 2] <- 1
##   x[(3 * n + 1):(4 * n), 1:2] <- 1
##   y <- c(y0, y1, y2, y12)
##   
##   out <- list(
##     x = x,
##     y = y,
##     eff1_vec = eff1_vec,
##     eff2_vec = eff2_vec,
##     int_eff_vec = int_eff_vec
##   )
## }
```

**Input Parameters**

```
## $n
## [1] 500
## 
## $beta0
## [1] 0
## 
## $beta1
## [1] -1
## 
## $beta2
## [1] -1
## 
## $beta12
## [1] 0
## 
## $eff1
## [1] 0.66
## 
## $eff2
## [1] 0.8
## 
## $int_eff
## [1] 0.93
## 
## $err_fun
## function (n, mean = 0, sd = 1) 
## .Call(C_rnorm, n, mean, sd)
## <bytecode: 0x7feab7ce5bc0>
## <environment: namespace:stats>
## 
## $sd
## [1] 1
```

##### Efficiency = 1

In this data-generating process, we simulated data as described above but set the simulated gene-silencing efficiencies to be 1. That is, \(\pi\_{01} = \pi\_{10} = \pi\_{11} = 1\). This data-generating process is akin to the setting where the gene-silencing efficiences are perfect, and there is no variation in gene-silencing efficiences across different batches/gene-silencing conditions.

**Function**

```
## function(n, beta0, beta1, beta2, beta12,
##                                eff1 = 1, eff2 = 1, int_eff = 1, 
##                                err_fun = rnorm, ...) {
##   eff1_vec <- rbinom(n, 1, eff1)
##   eff2_vec <- rbinom(n, 1, eff2)
##   int_eff_vec <- rbinom(n, 1, int_eff)
##   
##   y0 <- beta0 + err_fun(n, ...)
##   y1 <- beta0 + beta1 * eff1_vec + err_fun(n, ...)
##   y2 <- beta0 + beta2 * eff2_vec + err_fun(n, ...)
##   y12 <- beta0 + (beta1 + beta2 + beta12) * int_eff_vec + err_fun(n, ...)
##   
##   x <- matrix(0, nrow = 4 * n, ncol = 2)
##   x[(n + 1):(2 * n), 1] <- 1
##   x[(2 * n + 1):(3 * n), 2] <- 1
##   x[(3 * n + 1):(4 * n), 1:2] <- 1
##   y <- c(y0, y1, y2, y12)
##   
##   out <- list(
##     x = x,
##     y = y,
##     eff1_vec = eff1_vec,
##     eff2_vec = eff2_vec,
##     int_eff_vec = int_eff_vec
##   )
## }
```

**Input Parameters**

```
## $n
## [1] 500
## 
## $beta0
## [1] 0
## 
## $beta1
## [1] -1
## 
## $beta2
## [1] -1
## 
## $beta12
## [1] 0
## 
## $eff1
## [1] 1
## 
## $eff2
## [1] 1
## 
## $int_eff
## [1] 1
## 
## $err_fun
## function (n, mean = 0, sd = 1) 
## .Call(C_rnorm, n, mean, sd)
## <bytecode: 0x7feab664cf30>
## <environment: namespace:stats>
## 
## $sd
## [1] 1
```

#### Methods and Evaluation

##### Methods

###### Quantile Regression (tau = 0.5)

In Wang et al. (2023), epistasis was assessed in the experimental data by fitting a quantile regression model and testing whether the interaction effect term was significantly different from 0. In this simulation study, we thus follow suit. Given the simulated data, we fit a quantile regression model:

\[y\_i = \beta\_0 + \beta\_1 x\_{i1} + \beta\_2 x\_{i2} + \beta\_{12} x\_{i1} x\_{i2} + \epsilon\_i,\]

where \(y\_i\) is the diameter (\(\mu m\)) of cell \(i\), \(x\_{ij}\) is an indicator of whether cell \(i\) belonged to an experimental batch where gene \(j\) was silenced, and \(\epsilon\_i\) is the random error or noise term for cell \(i\). Using this regression model, we performed a t-test to assess the null hypothesis that \(\beta\_{12} = 0\) (i.e., no epistatic effect) versus the alternative hypothesis that \(\beta\_{12} \neq 0\) (i.e., a non-zero epistatic effect) at the 0.5 quantile level (i.e., median).

We note that in Wang et al. (2023), percentile bootstrap hypothesis tests were conducted in addition to the t-tests, but as they gave similar conclusions in Wang et al. (2023), we omit the bootstrap hypothesis tests here to reduce the computational burden.

**Function**

```
## function(x, y, tau = 0.5, min_thr = 1e-16, ...) {
##   fit_df <- data.frame(.y = y, x)
##   fit <- quantreg::rq(formula = .y ~ X1 * X2, tau = tau, data = fit_df)
##   fit_summary <- summary(fit, se = "boot")$coefficients %>%
##     as.data.frame() %>%
##     tibble::rownames_to_column("Term") %>%
##     dplyr::rename(Coefficient = Value) %>%
##     dplyr::filter(Term == "X1:X2") %>%
##     dplyr::mutate(`Pr(>|t|)` = max(min_thr, `Pr(>|t|)`))
##   return(fit_summary)
##   # return(broom::tidy(fit_summary))
## }
```

**Input Parameters**

```
## list()
```

##### Evaluation

###### P-values Summary

N/A

**Function**

```
## function (fit_results, vary_params = NULL, nested_cols = NULL, 
##     feature_col, imp_col, group_cols = NULL, na_rm = FALSE, summary_funs = c("mean", 
##         "median", "min", "max", "sd", "raw"), custom_summary_funs = NULL, 
##     eval_id = "feature_importance") 
## {
##     group_vars <- c(".dgp_name", ".method_name", vary_params, 
##         group_cols, feature_col)
##     eval_tib <- eval_feature_importance(fit_results = fit_results, 
##         vary_params = vary_params, nested_cols = nested_cols, 
##         feature_col = feature_col, imp_col = imp_col, group_cols = group_cols) %>% 
##         dplyr::group_by(dplyr::across(tidyselect::all_of(group_vars)))
##     eval_summary <- eval_summarizer(eval_data = eval_tib, eval_id = eval_id, 
##         value_col = imp_col, summary_funs = summary_funs, custom_summary_funs = custom_summary_funs, 
##         na_rm = na_rm)
##     return(eval_summary)
## }
## <bytecode: 0x7feacc42f890>
## <environment: namespace:simChef>
```

**Input Parameters**

```
## $eval_id
## [1] "pval"
## 
## $nested_cols
## NULL
## 
## $feature_col
## [1] "Term"
## 
## $imp_col
## [1] "Pr(>|t|)"
```

#### Visualizations

##### P-values Plot

In this visualization, we plot the distribution of p-values corresponding to the interaction effect term (i.e., \(\beta\_{12}\)) across 100 simulation replicates. The red line indicates the \(\alpha = 0.05\) significance level. The y-axis is on the log10 scale. When \(\beta\_{12} = 0\), we would hope that the p-values are greater than the significance threshold.

###### CCDC141-IGF1R, Diseased Simulation - CCDC141-IGF1R, Diseased - Varying beta12

**Function**

```
## function(fit_results = NULL, eval_results, vary_params = NULL,
##                          eval_name, eval_id, add_ggplot_layers = NULL) {
##   vary_params <- unique(vary_params)
##   plt_df <- eval_results[[eval_name]] %>%
##     dplyr::filter(!is.na(!!rlang::sym(vary_params)))
##   plt <- plt_df %>%
##     tidyr::unnest(cols = tidyselect::all_of(sprintf("raw_%s", eval_id))) %>%
##     vdocs::plot_boxplot(
##       x_str = vary_params, y_str = sprintf("raw_%s", eval_id)
##     ) +
##     ggplot2::labs(y = "P-value")
##   if (vary_params == "sd") {
##     plt <- plt + ggplot2::labs(x = bquote("SD of Noise" ~ (sigma)))
##   } else if (vary_params == "beta12") {
##     plt <- plt + ggplot2::labs(x = bquote("Interaction Effect" ~ (beta[12])))
##   }
##   n_dgps <- length(unique(plt_df$.dgp_name))
##   if (n_dgps > 1) {
##     plt <- plt + ggplot2::facet_wrap(~ .dgp_name)
##   }
##   if (!is.null(add_ggplot_layers)) {
##     for (ggplot_layer in add_ggplot_layers) {
##       plt <- plt + ggplot_layer
##     }
##   }
##   return(plt)
## }
## <bytecode: 0x7feab36244e0>
```

**Input Parameters**

```
## $eval_name
## [1] "P-values Summary"
## 
## $eval_id
## [1] "pval"
## 
## $add_ggplot_layers
## $add_ggplot_layers[[1]]
## <ScaleContinuousPosition>
##  Range:  
##  Limits:    0 --    1
## 
## $add_ggplot_layers[[2]]
## mapping: yintercept = ~yintercept 
## geom_hline: na.rm = FALSE
## stat_identity: na.rm = FALSE
## position_identity 
## 
## $add_ggplot_layers[[3]]
## <ggproto object: Class CoordCartesian, Coord, gg>
##     aspect: function
##     backtransform_range: function
##     clip: on
##     default: FALSE
##     distance: function
##     expand: TRUE
##     is_free: function
##     is_linear: function
##     labels: function
##     limits: list
##     modify_scales: function
##     range: function
##     render_axis_h: function
##     render_axis_v: function
##     render_bg: function
##     render_fg: function
##     setup_data: function
##     setup_layout: function
##     setup_panel_guides: function
##     setup_panel_params: function
##     setup_params: function
##     train_panel_guides: function
##     transform: function
##     super:  <ggproto object: Class CoordCartesian, Coord, gg>
```

###### CCDC141-IGF1R, Diseased Simulation - CCDC141-IGF1R, Diseased - Varying sd

**Function**

```
## function(fit_results = NULL, eval_results, vary_params = NULL,
##                          eval_name, eval_id, add_ggplot_layers = NULL) {
##   vary_params <- unique(vary_params)
##   plt_df <- eval_results[[eval_name]] %>%
##     dplyr::filter(!is.na(!!rlang::sym(vary_params)))
##   plt <- plt_df %>%
##     tidyr::unnest(cols = tidyselect::all_of(sprintf("raw_%s", eval_id))) %>%
##     vdocs::plot_boxplot(
##       x_str = vary_params, y_str = sprintf("raw_%s", eval_id)
##     ) +
##     ggplot2::labs(y = "P-value")
##   if (vary_params == "sd") {
##     plt <- plt + ggplot2::labs(x = bquote("SD of Noise" ~ (sigma)))
##   } else if (vary_params == "beta12") {
##     plt <- plt + ggplot2::labs(x = bquote("Interaction Effect" ~ (beta[12])))
##   }
##   n_dgps <- length(unique(plt_df$.dgp_name))
##   if (n_dgps > 1) {
##     plt <- plt + ggplot2::facet_wrap(~ .dgp_name)
##   }
##   if (!is.null(add_ggplot_layers)) {
##     for (ggplot_layer in add_ggplot_layers) {
##       plt <- plt + ggplot_layer
##     }
##   }
##   return(plt)
## }
## <bytecode: 0x7feace7e3600>
```

**Input Parameters**

```
## $eval_name
## [1] "P-values Summary"
## 
## $eval_id
## [1] "pval"
## 
## $add_ggplot_layers
## $add_ggplot_layers[[1]]
## <ScaleContinuousPosition>
##  Range:  
##  Limits:    0 --    1
## 
## $add_ggplot_layers[[2]]
## mapping: yintercept = ~yintercept 
## geom_hline: na.rm = FALSE
## stat_identity: na.rm = FALSE
## position_identity 
## 
## $add_ggplot_layers[[3]]
## <ggproto object: Class CoordCartesian, Coord, gg>
##     aspect: function
##     backtransform_range: function
##     clip: on
##     default: FALSE
##     distance: function
##     expand: TRUE
##     is_free: function
##     is_linear: function
##     labels: function
##     limits: list
##     modify_scales: function
##     range: function
##     render_axis_h: function
##     render_axis_v: function
##     render_bg: function
##     render_fg: function
##     setup_data: function
##     setup_layout: function
##     setup_panel_guides: function
##     setup_panel_params: function
##     setup_params: function
##     train_panel_guides: function
##     transform: function
##     super:  <ggproto object: Class CoordCartesian, Coord, gg>
```

###### CCDC141-IGF1R, Healthy Simulation - CCDC141-IGF1R, Healthy - Varying beta12

**Function**

```
## function(fit_results = NULL, eval_results, vary_params = NULL,
##                          eval_name, eval_id, add_ggplot_layers = NULL) {
##   vary_params <- unique(vary_params)
##   plt_df <- eval_results[[eval_name]] %>%
##     dplyr::filter(!is.na(!!rlang::sym(vary_params)))
##   plt <- plt_df %>%
##     tidyr::unnest(cols = tidyselect::all_of(sprintf("raw_%s", eval_id))) %>%
##     vdocs::plot_boxplot(
##       x_str = vary_params, y_str = sprintf("raw_%s", eval_id)
##     ) +
##     ggplot2::labs(y = "P-value")
##   if (vary_params == "sd") {
##     plt <- plt + ggplot2::labs(x = bquote("SD of Noise" ~ (sigma)))
##   } else if (vary_params == "beta12") {
##     plt <- plt + ggplot2::labs(x = bquote("Interaction Effect" ~ (beta[12])))
##   }
##   n_dgps <- length(unique(plt_df$.dgp_name))
##   if (n_dgps > 1) {
##     plt <- plt + ggplot2::facet_wrap(~ .dgp_name)
##   }
##   if (!is.null(add_ggplot_layers)) {
##     for (ggplot_layer in add_ggplot_layers) {
##       plt <- plt + ggplot_layer
##     }
##   }
##   return(plt)
## }
## <bytecode: 0x7feab254bca0>
```

**Input Parameters**

```
## $eval_name
## [1] "P-values Summary"
## 
## $eval_id
## [1] "pval"
## 
## $add_ggplot_layers
## $add_ggplot_layers[[1]]
## <ScaleContinuousPosition>
##  Range:  
##  Limits:    0 --    1
## 
## $add_ggplot_layers[[2]]
## mapping: yintercept = ~yintercept 
## geom_hline: na.rm = FALSE
## stat_identity: na.rm = FALSE
## position_identity 
## 
## $add_ggplot_layers[[3]]
## <ggproto object: Class CoordCartesian, Coord, gg>
##     aspect: function
##     backtransform_range: function
##     clip: on
##     default: FALSE
##     distance: function
##     expand: TRUE
##     is_free: function
##     is_linear: function
##     labels: function
##     limits: list
##     modify_scales: function
##     range: function
##     render_axis_h: function
##     render_axis_v: function
##     render_bg: function
##     render_fg: function
##     setup_data: function
##     setup_layout: function
##     setup_panel_guides: function
##     setup_panel_params: function
##     setup_params: function
##     train_panel_guides: function
##     transform: function
##     super:  <ggproto object: Class CoordCartesian, Coord, gg>
```

###### CCDC141-IGF1R, Healthy Simulation - CCDC141-IGF1R, Healthy - Varying sd

**Function**

```
## function(fit_results = NULL, eval_results, vary_params = NULL,
##                          eval_name, eval_id, add_ggplot_layers = NULL) {
##   vary_params <- unique(vary_params)
##   plt_df <- eval_results[[eval_name]] %>%
##     dplyr::filter(!is.na(!!rlang::sym(vary_params)))
##   plt <- plt_df %>%
##     tidyr::unnest(cols = tidyselect::all_of(sprintf("raw_%s", eval_id))) %>%
##     vdocs::plot_boxplot(
##       x_str = vary_params, y_str = sprintf("raw_%s", eval_id)
##     ) +
##     ggplot2::labs(y = "P-value")
##   if (vary_params == "sd") {
##     plt <- plt + ggplot2::labs(x = bquote("SD of Noise" ~ (sigma)))
##   } else if (vary_params == "beta12") {
##     plt <- plt + ggplot2::labs(x = bquote("Interaction Effect" ~ (beta[12])))
##   }
##   n_dgps <- length(unique(plt_df$.dgp_name))
##   if (n_dgps > 1) {
##     plt <- plt + ggplot2::facet_wrap(~ .dgp_name)
##   }
##   if (!is.null(add_ggplot_layers)) {
##     for (ggplot_layer in add_ggplot_layers) {
##       plt <- plt + ggplot_layer
##     }
##   }
##   return(plt)
## }
## <bytecode: 0x7feab4433fb8>
```

**Input Parameters**

```
## $eval_name
## [1] "P-values Summary"
## 
## $eval_id
## [1] "pval"
## 
## $add_ggplot_layers
## $add_ggplot_layers[[1]]
## <ScaleContinuousPosition>
##  Range:  
##  Limits:    0 --    1
## 
## $add_ggplot_layers[[2]]
## mapping: yintercept = ~yintercept 
## geom_hline: na.rm = FALSE
## stat_identity: na.rm = FALSE
## position_identity 
## 
## $add_ggplot_layers[[3]]
## <ggproto object: Class CoordCartesian, Coord, gg>
##     aspect: function
##     backtransform_range: function
##     clip: on
##     default: FALSE
##     distance: function
##     expand: TRUE
##     is_free: function
##     is_linear: function
##     labels: function
##     limits: list
##     modify_scales: function
##     range: function
##     render_axis_h: function
##     render_axis_v: function
##     render_bg: function
##     render_fg: function
##     setup_data: function
##     setup_layout: function
##     setup_panel_guides: function
##     setup_panel_params: function
##     setup_params: function
##     train_panel_guides: function
##     transform: function
##     super:  <ggproto object: Class CoordCartesian, Coord, gg>
```

###### CCDC141-TTN, Diseased Simulation - CCDC141-TTN, Diseased - Varying beta12

**Function**

```
## function(fit_results = NULL, eval_results, vary_params = NULL,
##                          eval_name, eval_id, add_ggplot_layers = NULL) {
##   vary_params <- unique(vary_params)
##   plt_df <- eval_results[[eval_name]] %>%
##     dplyr::filter(!is.na(!!rlang::sym(vary_params)))
##   plt <- plt_df %>%
##     tidyr::unnest(cols = tidyselect::all_of(sprintf("raw_%s", eval_id))) %>%
##     vdocs::plot_boxplot(
##       x_str = vary_params, y_str = sprintf("raw_%s", eval_id)
##     ) +
##     ggplot2::labs(y = "P-value")
##   if (vary_params == "sd") {
##     plt <- plt + ggplot2::labs(x = bquote("SD of Noise" ~ (sigma)))
##   } else if (vary_params == "beta12") {
##     plt <- plt + ggplot2::labs(x = bquote("Interaction Effect" ~ (beta[12])))
##   }
##   n_dgps <- length(unique(plt_df$.dgp_name))
##   if (n_dgps > 1) {
##     plt <- plt + ggplot2::facet_wrap(~ .dgp_name)
##   }
##   if (!is.null(add_ggplot_layers)) {
##     for (ggplot_layer in add_ggplot_layers) {
##       plt <- plt + ggplot_layer
##     }
##   }
##   return(plt)
## }
## <bytecode: 0x7feab517f108>
```

**Input Parameters**

```
## $eval_name
## [1] "P-values Summary"
## 
## $eval_id
## [1] "pval"
## 
## $add_ggplot_layers
## $add_ggplot_layers[[1]]
## <ScaleContinuousPosition>
##  Range:  
##  Limits:    0 --    1
## 
## $add_ggplot_layers[[2]]
## mapping: yintercept = ~yintercept 
## geom_hline: na.rm = FALSE
## stat_identity: na.rm = FALSE
## position_identity 
## 
## $add_ggplot_layers[[3]]
## <ggproto object: Class CoordCartesian, Coord, gg>
##     aspect: function
##     backtransform_range: function
##     clip: on
##     default: FALSE
##     distance: function
##     expand: TRUE
##     is_free: function
##     is_linear: function
##     labels: function
##     limits: list
##     modify_scales: function
##     range: function
##     render_axis_h: function
##     render_axis_v: function
##     render_bg: function
##     render_fg: function
##     setup_data: function
##     setup_layout: function
##     setup_panel_guides: function
##     setup_panel_params: function
##     setup_params: function
##     train_panel_guides: function
##     transform: function
##     super:  <ggproto object: Class CoordCartesian, Coord, gg>
```

###### CCDC141-TTN, Diseased Simulation - CCDC141-TTN, Diseased - Varying sd

**Function**

```
## function(fit_results = NULL, eval_results, vary_params = NULL,
##                          eval_name, eval_id, add_ggplot_layers = NULL) {
##   vary_params <- unique(vary_params)
##   plt_df <- eval_results[[eval_name]] %>%
##     dplyr::filter(!is.na(!!rlang::sym(vary_params)))
##   plt <- plt_df %>%
##     tidyr::unnest(cols = tidyselect::all_of(sprintf("raw_%s", eval_id))) %>%
##     vdocs::plot_boxplot(
##       x_str = vary_params, y_str = sprintf("raw_%s", eval_id)
##     ) +
##     ggplot2::labs(y = "P-value")
##   if (vary_params == "sd") {
##     plt <- plt + ggplot2::labs(x = bquote("SD of Noise" ~ (sigma)))
##   } else if (vary_params == "beta12") {
##     plt <- plt + ggplot2::labs(x = bquote("Interaction Effect" ~ (beta[12])))
##   }
##   n_dgps <- length(unique(plt_df$.dgp_name))
##   if (n_dgps > 1) {
##     plt <- plt + ggplot2::facet_wrap(~ .dgp_name)
##   }
##   if (!is.null(add_ggplot_layers)) {
##     for (ggplot_layer in add_ggplot_layers) {
##       plt <- plt + ggplot_layer
##     }
##   }
##   return(plt)
## }
## <bytecode: 0x7feab45e2240>
```

**Input Parameters**

```
## $eval_name
## [1] "P-values Summary"
## 
## $eval_id
## [1] "pval"
## 
## $add_ggplot_layers
## $add_ggplot_layers[[1]]
## <ScaleContinuousPosition>
##  Range:  
##  Limits:    0 --    1
## 
## $add_ggplot_layers[[2]]
## mapping: yintercept = ~yintercept 
## geom_hline: na.rm = FALSE
## stat_identity: na.rm = FALSE
## position_identity 
## 
## $add_ggplot_layers[[3]]
## <ggproto object: Class CoordCartesian, Coord, gg>
##     aspect: function
##     backtransform_range: function
##     clip: on
##     default: FALSE
##     distance: function
##     expand: TRUE
##     is_free: function
##     is_linear: function
##     labels: function
##     limits: list
##     modify_scales: function
##     range: function
##     render_axis_h: function
##     render_axis_v: function
##     render_bg: function
##     render_fg: function
##     setup_data: function
##     setup_layout: function
##     setup_panel_guides: function
##     setup_panel_params: function
##     setup_params: function
##     train_panel_guides: function
##     transform: function
##     super:  <ggproto object: Class CoordCartesian, Coord, gg>
```

###### CCDC141-TTN, Healthy Simulation - CCDC141-TTN, Healthy - Varying beta12

**Function**

```
## function(fit_results = NULL, eval_results, vary_params = NULL,
##                          eval_name, eval_id, add_ggplot_layers = NULL) {
##   vary_params <- unique(vary_params)
##   plt_df <- eval_results[[eval_name]] %>%
##     dplyr::filter(!is.na(!!rlang::sym(vary_params)))
##   plt <- plt_df %>%
##     tidyr::unnest(cols = tidyselect::all_of(sprintf("raw_%s", eval_id))) %>%
##     vdocs::plot_boxplot(
##       x_str = vary_params, y_str = sprintf("raw_%s", eval_id)
##     ) +
##     ggplot2::labs(y = "P-value")
##   if (vary_params == "sd") {
##     plt <- plt + ggplot2::labs(x = bquote("SD of Noise" ~ (sigma)))
##   } else if (vary_params == "beta12") {
##     plt <- plt + ggplot2::labs(x = bquote("Interaction Effect" ~ (beta[12])))
##   }
##   n_dgps <- length(unique(plt_df$.dgp_name))
##   if (n_dgps > 1) {
##     plt <- plt + ggplot2::facet_wrap(~ .dgp_name)
##   }
##   if (!is.null(add_ggplot_layers)) {
##     for (ggplot_layer in add_ggplot_layers) {
##       plt <- plt + ggplot_layer
##     }
##   }
##   return(plt)
## }
## <bytecode: 0x7feab5405090>
```

**Input Parameters**

```
## $eval_name
## [1] "P-values Summary"
## 
## $eval_id
## [1] "pval"
## 
## $add_ggplot_layers
## $add_ggplot_layers[[1]]
## <ScaleContinuousPosition>
##  Range:  
##  Limits:    0 --    1
## 
## $add_ggplot_layers[[2]]
## mapping: yintercept = ~yintercept 
## geom_hline: na.rm = FALSE
## stat_identity: na.rm = FALSE
## position_identity 
## 
## $add_ggplot_layers[[3]]
## <ggproto object: Class CoordCartesian, Coord, gg>
##     aspect: function
##     backtransform_range: function
##     clip: on
##     default: FALSE
##     distance: function
##     expand: TRUE
##     is_free: function
##     is_linear: function
##     labels: function
##     limits: list
##     modify_scales: function
##     range: function
##     render_axis_h: function
##     render_axis_v: function
##     render_bg: function
##     render_fg: function
##     setup_data: function
##     setup_layout: function
##     setup_panel_guides: function
##     setup_panel_params: function
##     setup_params: function
##     train_panel_guides: function
##     transform: function
##     super:  <ggproto object: Class CoordCartesian, Coord, gg>
```

###### CCDC141-TTN, Healthy Simulation - CCDC141-TTN, Healthy - Varying sd

**Function**

```
## function(fit_results = NULL, eval_results, vary_params = NULL,
##                          eval_name, eval_id, add_ggplot_layers = NULL) {
##   vary_params <- unique(vary_params)
##   plt_df <- eval_results[[eval_name]] %>%
##     dplyr::filter(!is.na(!!rlang::sym(vary_params)))
##   plt <- plt_df %>%
##     tidyr::unnest(cols = tidyselect::all_of(sprintf("raw_%s", eval_id))) %>%
##     vdocs::plot_boxplot(
##       x_str = vary_params, y_str = sprintf("raw_%s", eval_id)
##     ) +
##     ggplot2::labs(y = "P-value")
##   if (vary_params == "sd") {
##     plt <- plt + ggplot2::labs(x = bquote("SD of Noise" ~ (sigma)))
##   } else if (vary_params == "beta12") {
##     plt <- plt + ggplot2::labs(x = bquote("Interaction Effect" ~ (beta[12])))
##   }
##   n_dgps <- length(unique(plt_df$.dgp_name))
##   if (n_dgps > 1) {
##     plt <- plt + ggplot2::facet_wrap(~ .dgp_name)
##   }
##   if (!is.null(add_ggplot_layers)) {
##     for (ggplot_layer in add_ggplot_layers) {
##       plt <- plt + ggplot_layer
##     }
##   }
##   return(plt)
## }
## <bytecode: 0x7feab278b738>
```

**Input Parameters**

```
## $eval_name
## [1] "P-values Summary"
## 
## $eval_id
## [1] "pval"
## 
## $add_ggplot_layers
## $add_ggplot_layers[[1]]
## <ScaleContinuousPosition>
##  Range:  
##  Limits:    0 --    1
## 
## $add_ggplot_layers[[2]]
## mapping: yintercept = ~yintercept 
## geom_hline: na.rm = FALSE
## stat_identity: na.rm = FALSE
## position_identity 
## 
## $add_ggplot_layers[[3]]
## <ggproto object: Class CoordCartesian, Coord, gg>
##     aspect: function
##     backtransform_range: function
##     clip: on
##     default: FALSE
##     distance: function
##     expand: TRUE
##     is_free: function
##     is_linear: function
##     labels: function
##     limits: list
##     modify_scales: function
##     range: function
##     render_axis_h: function
##     render_axis_v: function
##     render_bg: function
##     render_fg: function
##     setup_data: function
##     setup_layout: function
##     setup_panel_guides: function
##     setup_panel_params: function
##     setup_params: function
##     train_panel_guides: function
##     transform: function
##     super:  <ggproto object: Class CoordCartesian, Coord, gg>
```

###### Efficiency = 1 Simulation - Efficiency = 1 - Varying beta12

**Function**

```
## function(fit_results = NULL, eval_results, vary_params = NULL,
##                          eval_name, eval_id, add_ggplot_layers = NULL) {
##   vary_params <- unique(vary_params)
##   plt_df <- eval_results[[eval_name]] %>%
##     dplyr::filter(!is.na(!!rlang::sym(vary_params)))
##   plt <- plt_df %>%
##     tidyr::unnest(cols = tidyselect::all_of(sprintf("raw_%s", eval_id))) %>%
##     vdocs::plot_boxplot(
##       x_str = vary_params, y_str = sprintf("raw_%s", eval_id)
##     ) +
##     ggplot2::labs(y = "P-value")
##   if (vary_params == "sd") {
##     plt <- plt + ggplot2::labs(x = bquote("SD of Noise" ~ (sigma)))
##   } else if (vary_params == "beta12") {
##     plt <- plt + ggplot2::labs(x = bquote("Interaction Effect" ~ (beta[12])))
##   }
##   n_dgps <- length(unique(plt_df$.dgp_name))
##   if (n_dgps > 1) {
##     plt <- plt + ggplot2::facet_wrap(~ .dgp_name)
##   }
##   if (!is.null(add_ggplot_layers)) {
##     for (ggplot_layer in add_ggplot_layers) {
##       plt <- plt + ggplot_layer
##     }
##   }
##   return(plt)
## }
## <bytecode: 0x7feab5a03530>
```

**Input Parameters**

```
## $eval_name
## [1] "P-values Summary"
## 
## $eval_id
## [1] "pval"
## 
## $add_ggplot_layers
## $add_ggplot_layers[[1]]
## <ScaleContinuousPosition>
##  Range:  
##  Limits:    0 --    1
## 
## $add_ggplot_layers[[2]]
## mapping: yintercept = ~yintercept 
## geom_hline: na.rm = FALSE
## stat_identity: na.rm = FALSE
## position_identity 
## 
## $add_ggplot_layers[[3]]
## <ggproto object: Class CoordCartesian, Coord, gg>
##     aspect: function
##     backtransform_range: function
##     clip: on
##     default: FALSE
##     distance: function
##     expand: TRUE
##     is_free: function
##     is_linear: function
##     labels: function
##     limits: list
##     modify_scales: function
##     range: function
##     render_axis_h: function
##     render_axis_v: function
##     render_bg: function
##     render_fg: function
##     setup_data: function
##     setup_layout: function
##     setup_panel_guides: function
##     setup_panel_params: function
##     setup_params: function
##     train_panel_guides: function
##     transform: function
##     super:  <ggproto object: Class CoordCartesian, Coord, gg>
```

###### Efficiency = 1 Simulation - Efficiency = 1 - Varying sd

**Function**

```
## function(fit_results = NULL, eval_results, vary_params = NULL,
##                          eval_name, eval_id, add_ggplot_layers = NULL) {
##   vary_params <- unique(vary_params)
##   plt_df <- eval_results[[eval_name]] %>%
##     dplyr::filter(!is.na(!!rlang::sym(vary_params)))
##   plt <- plt_df %>%
##     tidyr::unnest(cols = tidyselect::all_of(sprintf("raw_%s", eval_id))) %>%
##     vdocs::plot_boxplot(
##       x_str = vary_params, y_str = sprintf("raw_%s", eval_id)
##     ) +
##     ggplot2::labs(y = "P-value")
##   if (vary_params == "sd") {
##     plt <- plt + ggplot2::labs(x = bquote("SD of Noise" ~ (sigma)))
##   } else if (vary_params == "beta12") {
##     plt <- plt + ggplot2::labs(x = bquote("Interaction Effect" ~ (beta[12])))
##   }
##   n_dgps <- length(unique(plt_df$.dgp_name))
##   if (n_dgps > 1) {
##     plt <- plt + ggplot2::facet_wrap(~ .dgp_name)
##   }
##   if (!is.null(add_ggplot_layers)) {
##     for (ggplot_layer in add_ggplot_layers) {
##       plt <- plt + ggplot_layer
##     }
##   }
##   return(plt)
## }
## <bytecode: 0x7feab5a41ce0>
```

**Input Parameters**

```
## $eval_name
## [1] "P-values Summary"
## 
## $eval_id
## [1] "pval"
## 
## $add_ggplot_layers
## $add_ggplot_layers[[1]]
## <ScaleContinuousPosition>
##  Range:  
##  Limits:    0 --    1
## 
## $add_ggplot_layers[[2]]
## mapping: yintercept = ~yintercept 
## geom_hline: na.rm = FALSE
## stat_identity: na.rm = FALSE
## position_identity 
## 
## $add_ggplot_layers[[3]]
## <ggproto object: Class CoordCartesian, Coord, gg>
##     aspect: function
##     backtransform_range: function
##     clip: on
##     default: FALSE
##     distance: function
##     expand: TRUE
##     is_free: function
##     is_linear: function
##     labels: function
##     limits: list
##     modify_scales: function
##     range: function
##     render_axis_h: function
##     render_axis_v: function
##     render_bg: function
##     render_fg: function
##     setup_data: function
##     setup_layout: function
##     setup_panel_guides: function
##     setup_panel_params: function
##     setup_params: function
##     train_panel_guides: function
##     transform: function
##     super:  <ggproto object: Class CoordCartesian, Coord, gg>
```

### CCDC141-IGF1R, Diseased Simulation

#### CCDC141-IGF1R, Diseased

##### Varying beta12

###### P-values Plot

**Parameter Values**

```
## $dgp
## $dgp$`CCDC141-IGF1R, Diseased`
## $dgp$`CCDC141-IGF1R, Diseased`$beta12
## [1]  0.0 -0.1 -0.2 -0.5 -1.0
## 
## 
## 
## $method
## list()
```

##### Varying sd

###### P-values Plot

**Parameter Values**

```
## $dgp
## $dgp$`CCDC141-IGF1R, Diseased`
## $dgp$`CCDC141-IGF1R, Diseased`$sd
## [1] 0.1 0.2 0.5 1.0 1.5 2.0
## 
## 
## 
## $method
## list()
```

### CCDC141-IGF1R, Healthy Simulation

#### CCDC141-IGF1R, Healthy

##### Varying beta12

###### P-values Plot

**Parameter Values**

```
## $dgp
## $dgp$`CCDC141-IGF1R, Healthy`
## $dgp$`CCDC141-IGF1R, Healthy`$beta12
## [1]  0.0 -0.1 -0.2 -0.5 -1.0
## 
## 
## 
## $method
## list()
```

##### Varying sd

###### P-values Plot

**Parameter Values**

```
## $dgp
## $dgp$`CCDC141-IGF1R, Healthy`
## $dgp$`CCDC141-IGF1R, Healthy`$sd
## [1] 0.1 0.2 0.5 1.0 1.5 2.0
## 
## 
## 
## $method
## list()
```

### CCDC141-TTN, Diseased Simulation

#### CCDC141-TTN, Diseased

##### Varying beta12

###### P-values Plot

**Parameter Values**

```
## $dgp
## $dgp$`CCDC141-TTN, Diseased`
## $dgp$`CCDC141-TTN, Diseased`$beta12
## [1]  0.0 -0.1 -0.2 -0.5 -1.0
## 
## 
## 
## $method
## list()
```

##### Varying sd

###### P-values Plot

**Parameter Values**

```
## $dgp
## $dgp$`CCDC141-TTN, Diseased`
## $dgp$`CCDC141-TTN, Diseased`$sd
## [1] 0.1 0.2 0.5 1.0 1.5 2.0
## 
## 
## 
## $method
## list()
```

### CCDC141-TTN, Healthy Simulation

#### CCDC141-TTN, Healthy

##### Varying beta12

###### P-values Plot

**Parameter Values**

```
## $dgp
## $dgp$`CCDC141-TTN, Healthy`
## $dgp$`CCDC141-TTN, Healthy`$beta12
## [1]  0.0 -0.1 -0.2 -0.5 -1.0
## 
## 
## 
## $method
## list()
```

##### Varying sd

###### P-values Plot

**Parameter Values**

```
## $dgp
## $dgp$`CCDC141-TTN, Healthy`
## $dgp$`CCDC141-TTN, Healthy`$sd
## [1] 0.1 0.2 0.5 1.0 1.5 2.0
## 
## 
## 
## $method
## list()
```

### Efficiency = 1 Simulation

#### Efficiency = 1

##### Varying beta12

###### P-values Plot

**Parameter Values**

```
## $dgp
## $dgp$`Efficiency = 1`
## $dgp$`Efficiency = 1`$beta12
## [1]  0.0 -0.1 -0.2 -0.5 -1.0
## 
## 
## 
## $method
## list()
```

##### Varying sd

###### P-values Plot

**Parameter Values**

```
## $dgp
## $dgp$`Efficiency = 1`
## $dgp$`Efficiency = 1`$sd
## [1] 0.1 0.2 0.5 1.0 1.5 2.0
## 
## 
## 
## $method
## list()
```

Code 

- Show All Code
- Hide All Code
- Download Rmd
